# Histopathological growth patterns of liver metastasis: updated consensus guidelines for pattern scoring, perspectives, and recent mechanistic insights

**DOI:** 10.1101/2022.04.07.22273504

**Authors:** Emily Latacz, Diederik Höppener, Ali Bohlok, Sophia Leduc, Sébastien Tabariès, Carlos Fernández Moro, Claire Lugassy, Hanna Nyström, Béla Bozóky, Giuseppe Floris, Natalie Geyer, Pnina Brodt, Laura Llado, Laura Van Mileghem, Maxim De Schepper, Ali W. Majeed, Anthoula Lazaris, Piet Dirix, Qianni Zhang, Stéphanie K. Petrillo, Sophie Vankerckhove, Ines Joye, Yannick Meyer, Alexander Gregorieff, Nuria Ruiz Roig, Fernando Vidal-Vanaclocha, Larsimont Denis, Rui Caetano Oliveira, Peter Metrakos, Dirk J. Grünhagen, Iris D. Nagtegaal, David G. Mollevi, William R Jarnagin, Michael I D’Angelica, Andrew R. Reynolds, Michail Doukas, Christine Desmedt, Luc Dirix, Vincent Donckier, Peter M. Siegel, Raymond Barnhill, Marco Gerling, Cornelis Verhoef, Peter B. Vermeulen

## Abstract

The first consensus guidelines for scoring the histopathological growth patterns (HGPs) of liver metastases were established in 2017. Since then, numerous studies have applied these guidelines, have further substantiated the potential clinical value of the HGPs in patients with liver metastases from various tumour types and are starting to shed light on the biology of the distinct HGPs. In the present guidelines, we give an overview of these studies, discuss novel strategies for predicting the HGPs of liver metastases, such as deep learning algorithms for whole slide histopathology images and medical imaging, and highlight liver metastasis animal models that exhibit features of the different HGPs. Based on a pooled analysis of large cohorts of patients with liver-metastatic colorectal cancer, we propose a new cut-off to categorize patients according to the HGPs. An up-to-date standard method for HGP assessment within liver metastases is also presented with the aim of incorporating HGPs into the decision-making processes surrounding the treatment of patients with liver metastatic cancer. Finally, we propose hypotheses on the cellular and molecular mechanisms that drive the biology of the different HGPs, opening some exciting pre-clinical and clinical research perspectives.

## Introduction

The histopathological growth patterns (HGPs) of liver metastases are a morphological reflection of the distinct ways in which cancer cells interact with the surrounding liver. These HGPs can be identified by light microscopy on tissue sections that include the metastasis-liver interface. In 2017, the first set of guidelines for scoring the growth patterns was published^1^. Since that time, numerous additional studies have utilized these consensus guidelines to score the HGPs of liver metastases. These studies, listed in Table 1, have further substantiated the clinical value of HGPs in hepatic metastases from colorectal cancer and extended this concept to other tumour types, such as breast carcinoma, melanoma, and pancreatic cancer. Moreover, these publications have significantly increased our understanding of HGP biology by describing the molecular and cellular differences between growth patterns by, for example, looking at growth pattern-specific immune responses^2–6^. In addition, attempts have been made to develop technologies for predicting HGPs using medical imaging and machine-learning algorithms^7–10^. Novel animal models for liver metastasis exhibiting features of the different HGPs are a particularly valuable development^11–17^. These models will allow us to: 1) perform functional validation of HGP-specific signalling pathways described in the clinical samples of liver metastases, 2) identify non-invasive surrogate markers for the different HGPs and 3) test the efficacy of new therapeutic strategies based on the HGPs.

**Table 1.**
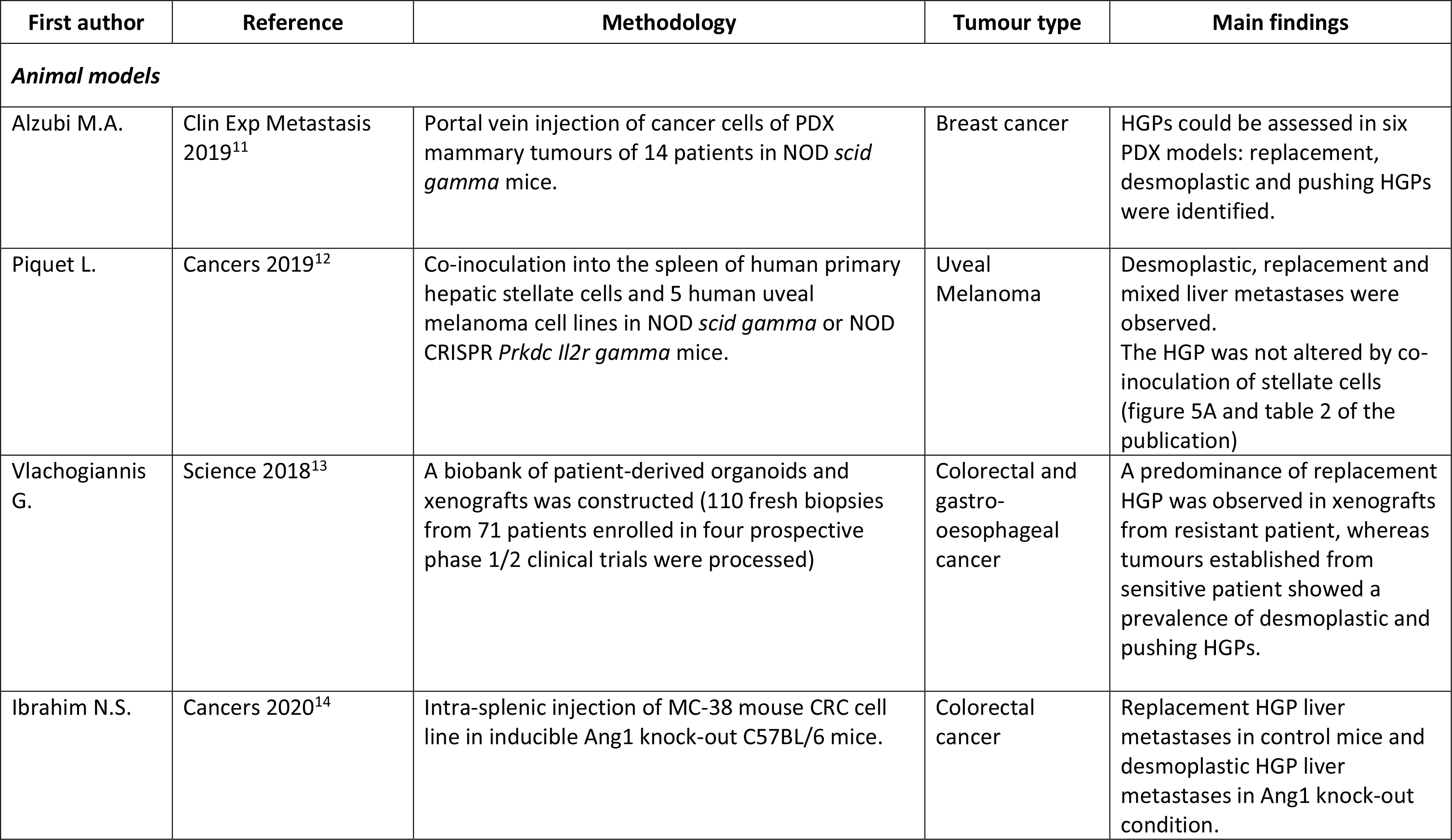
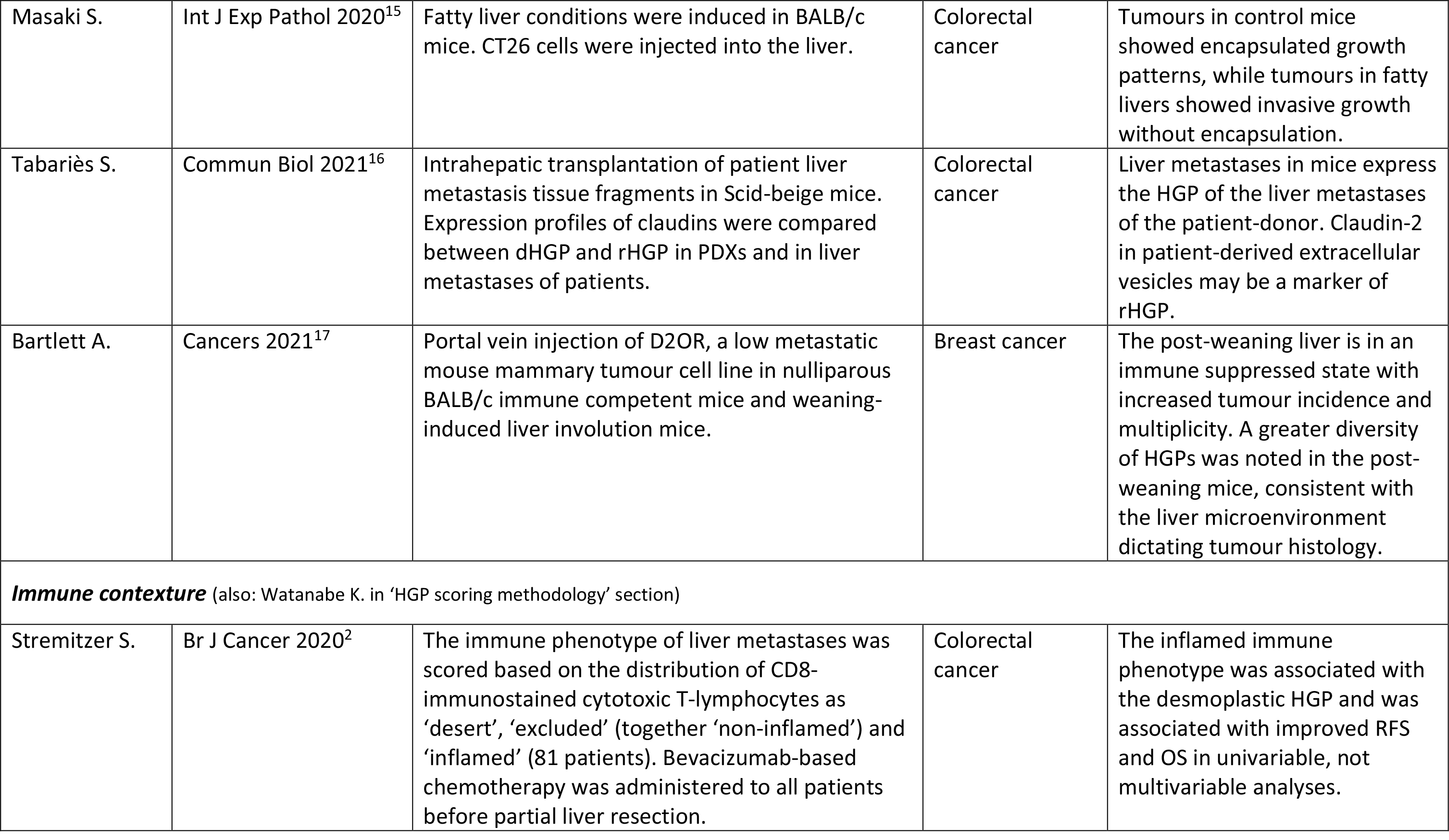
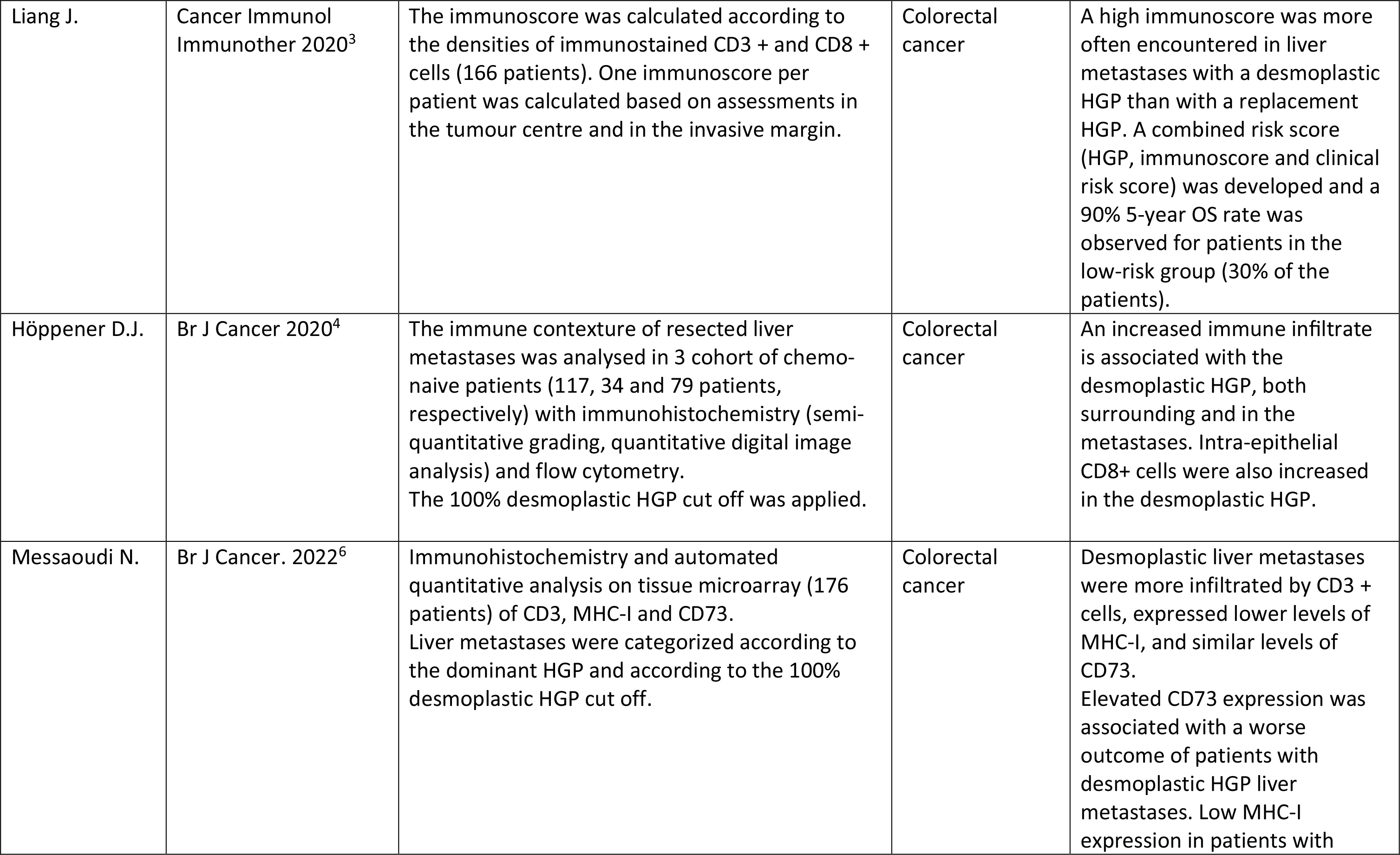
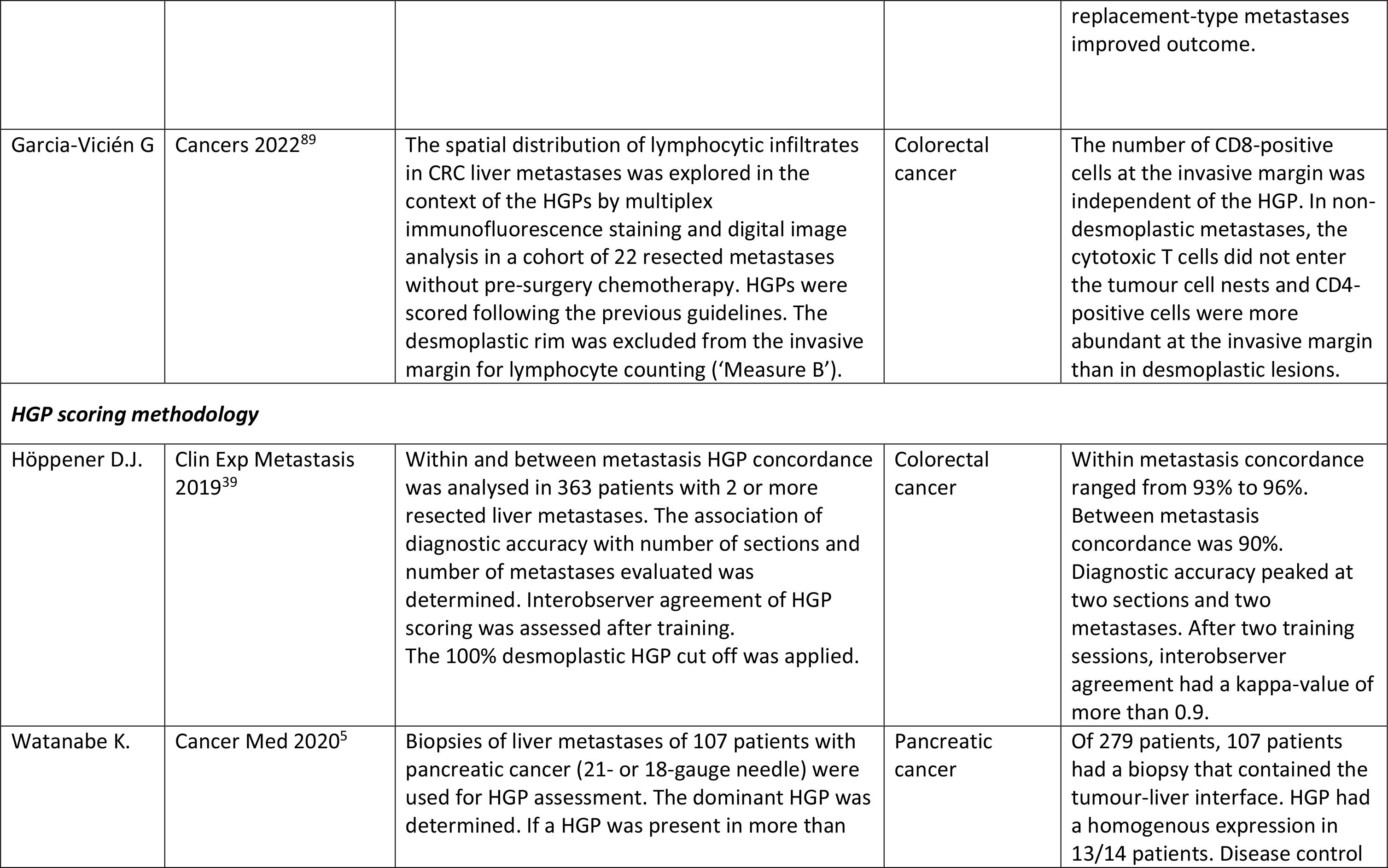
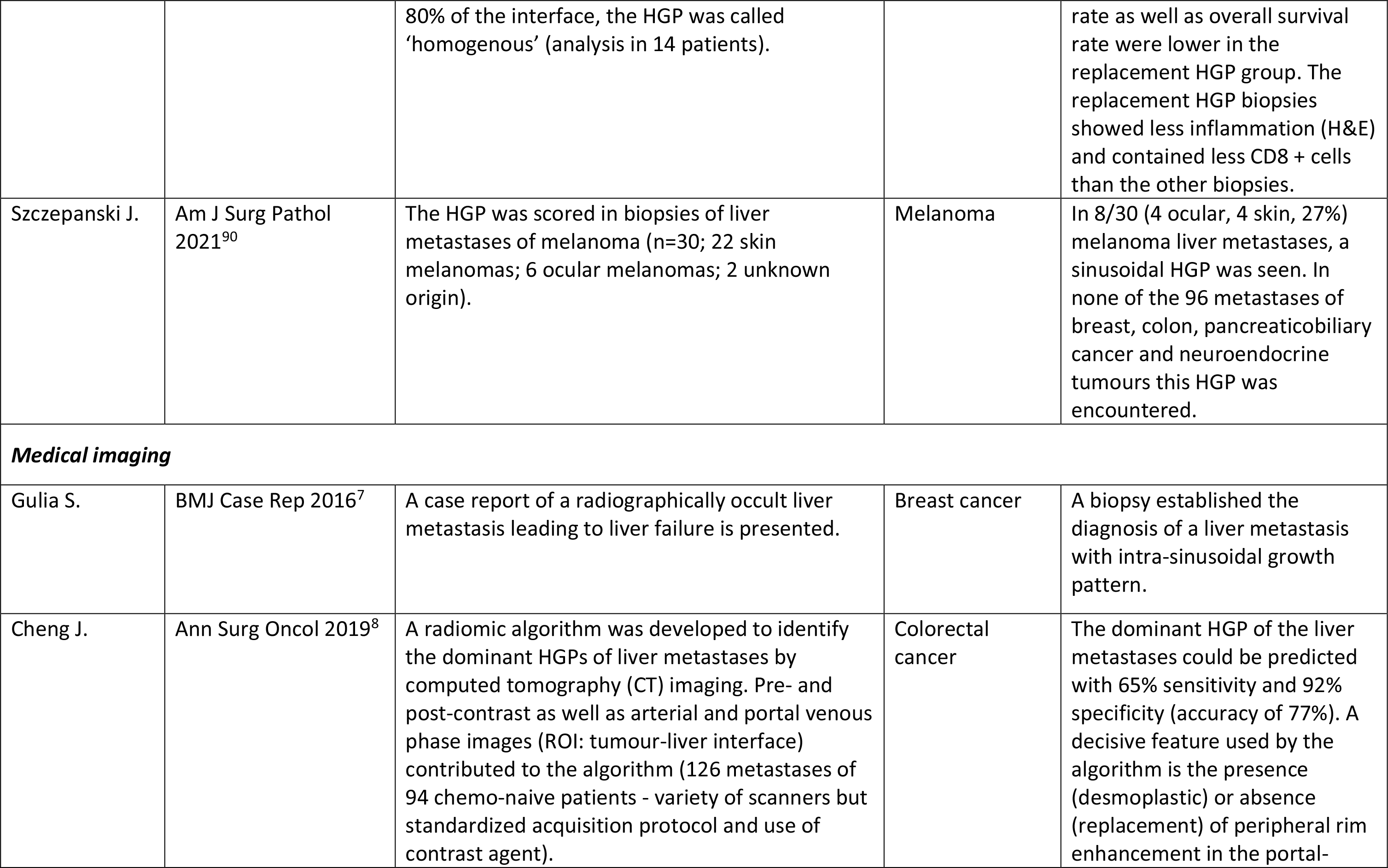
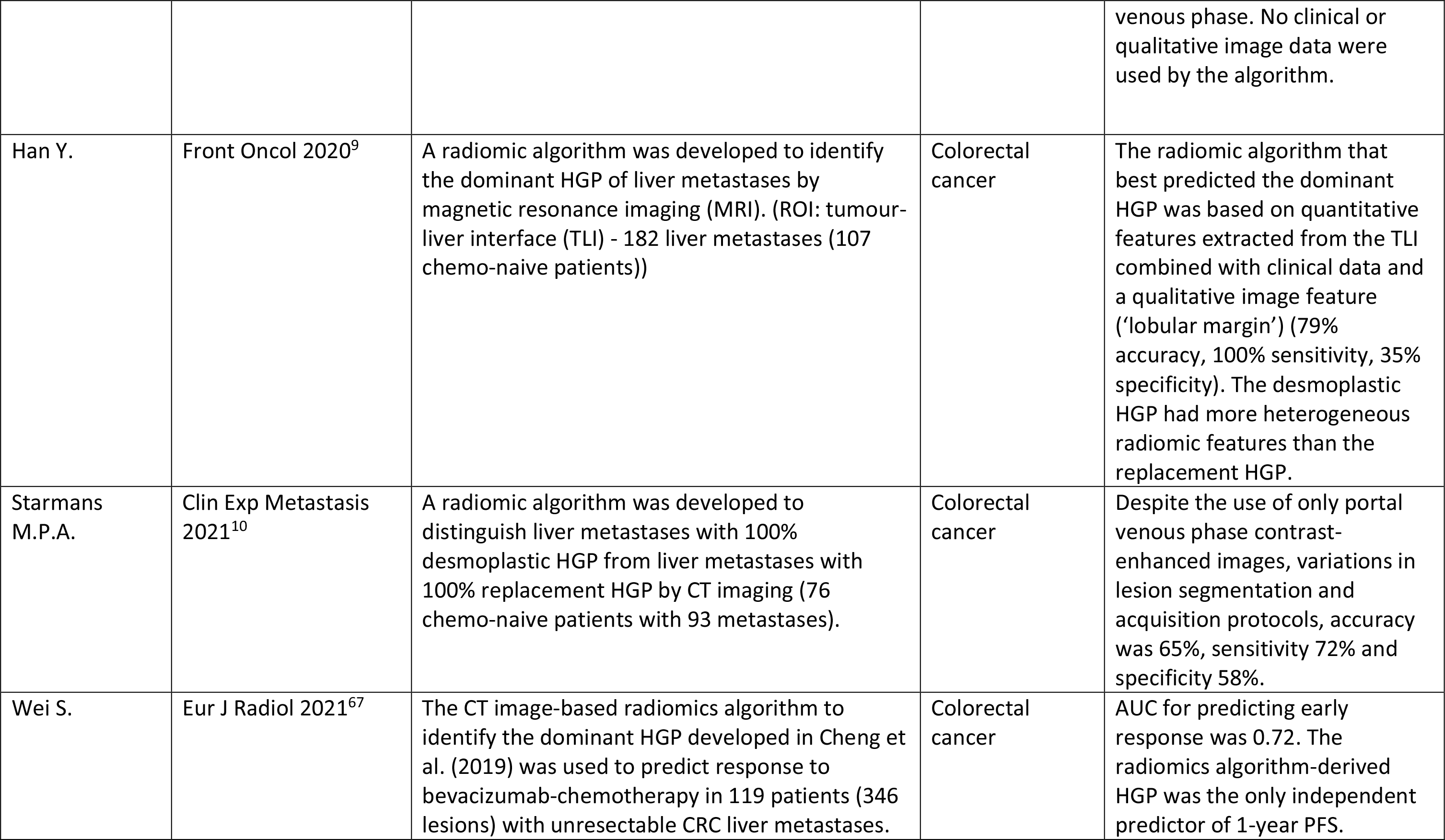
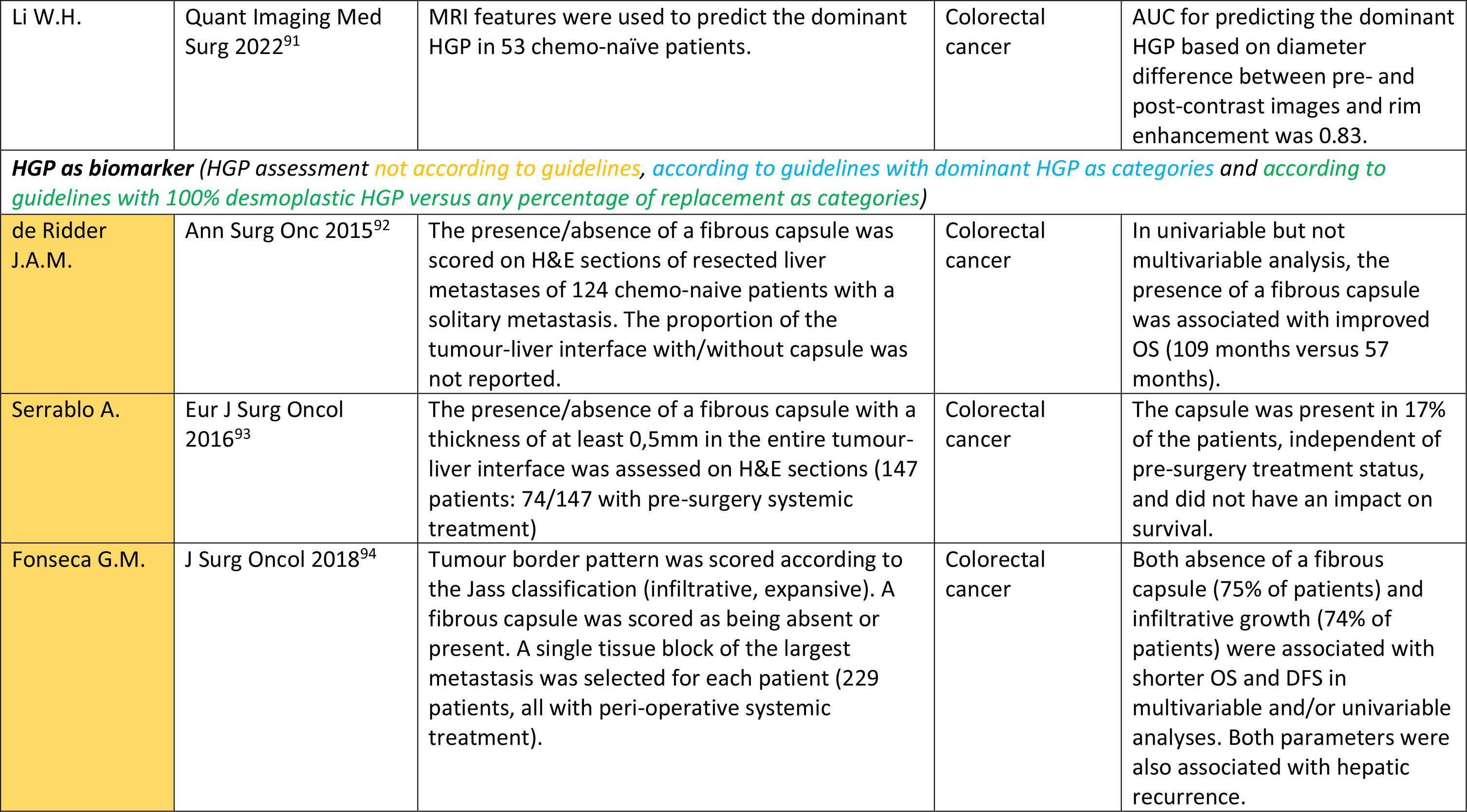
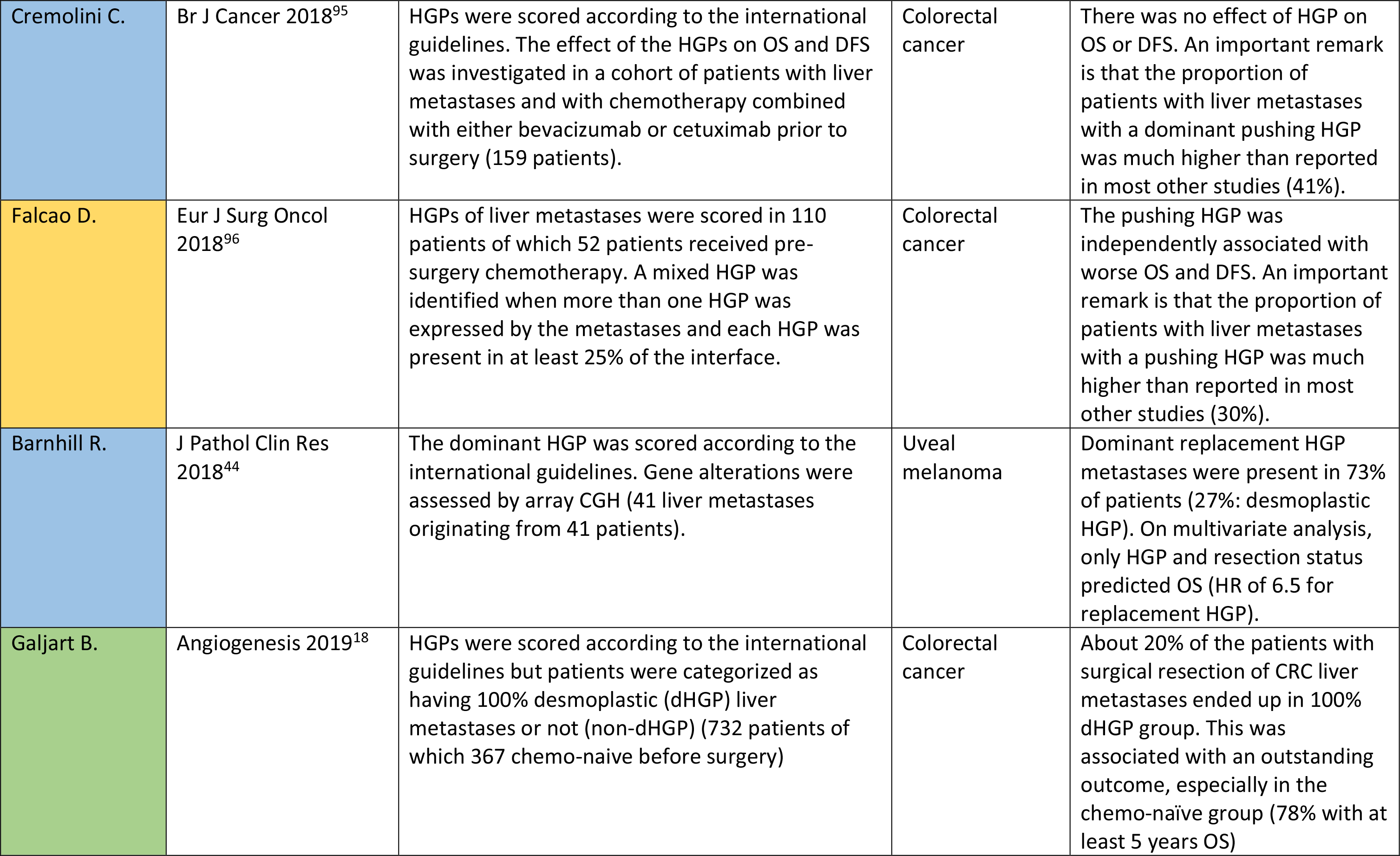
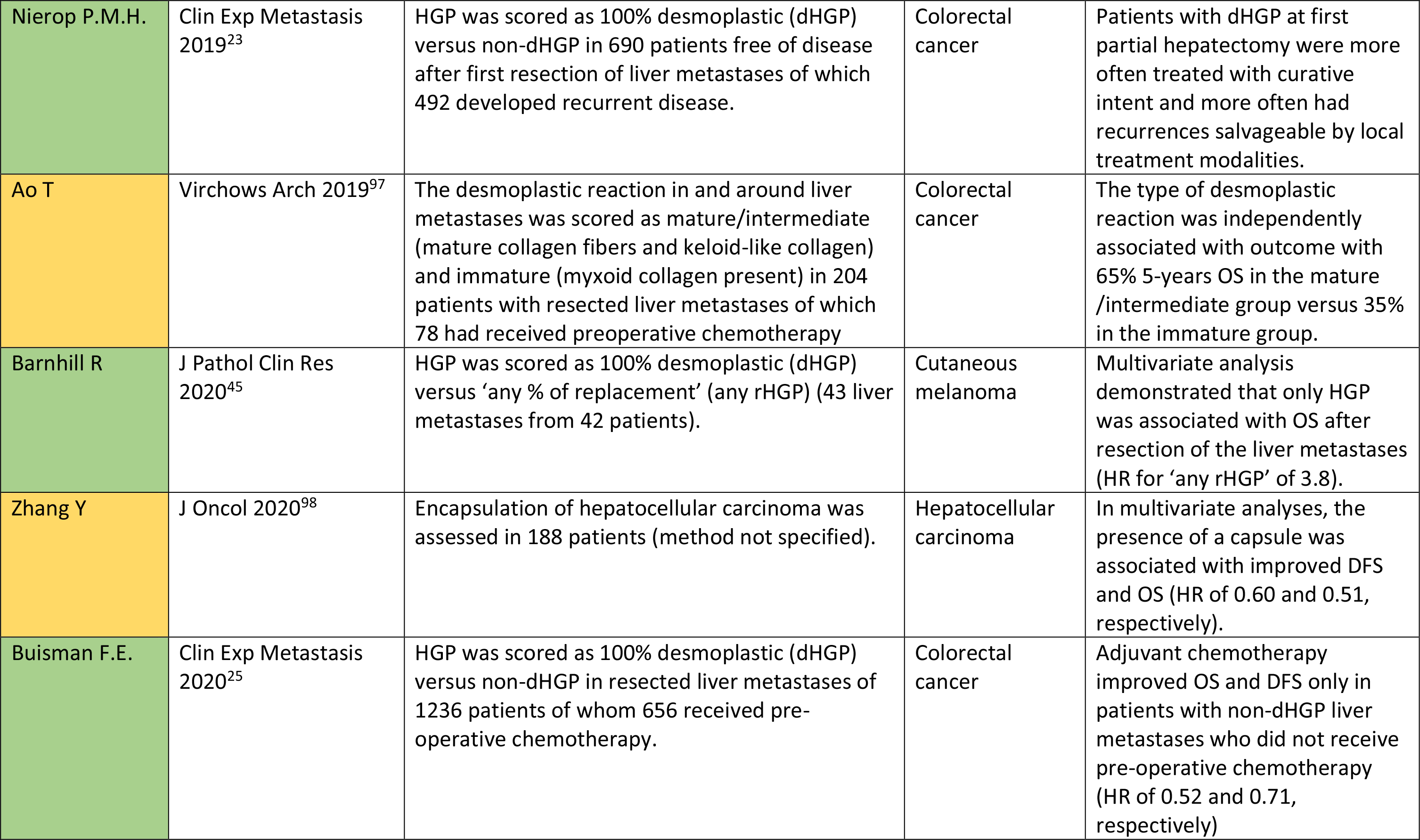
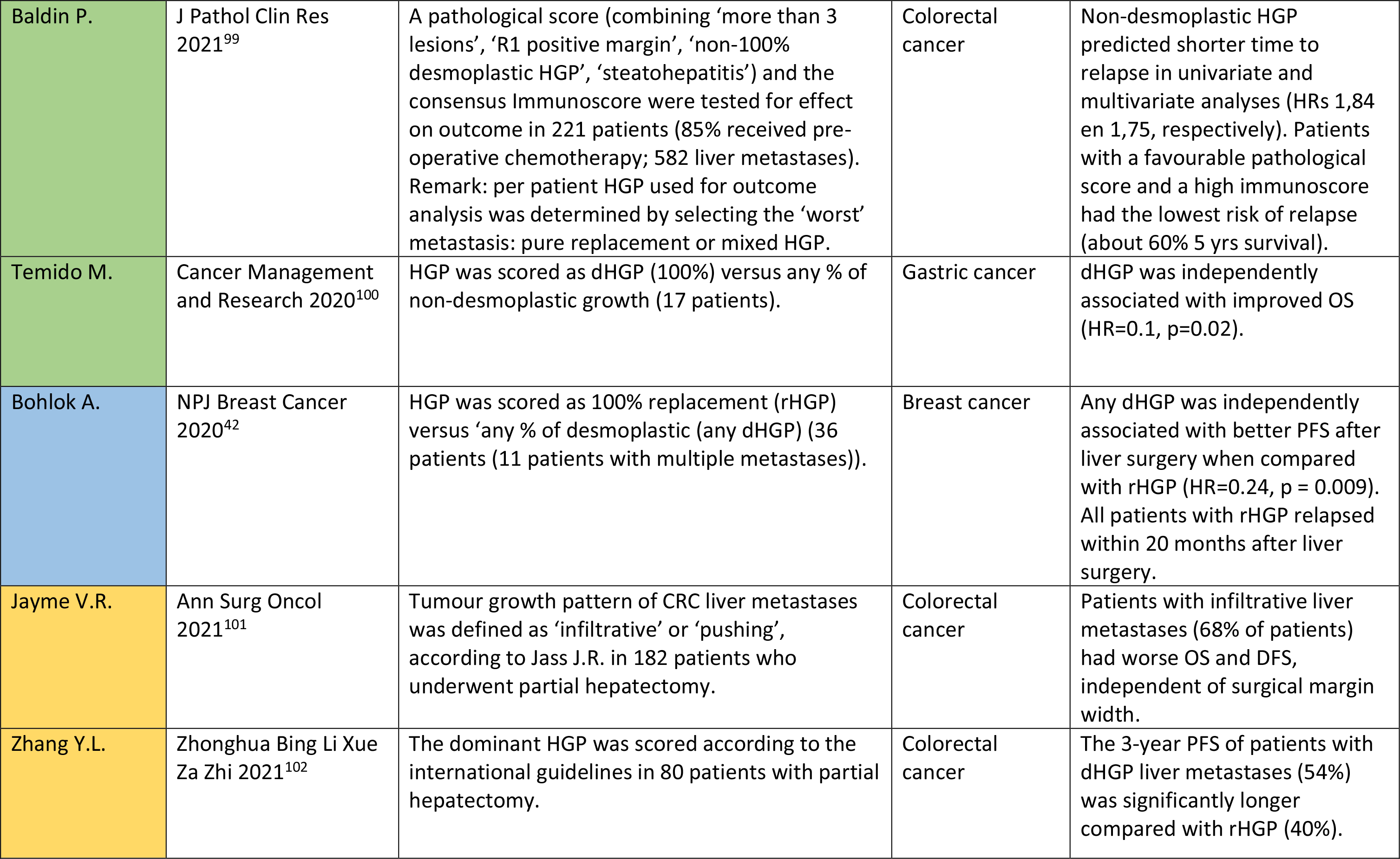
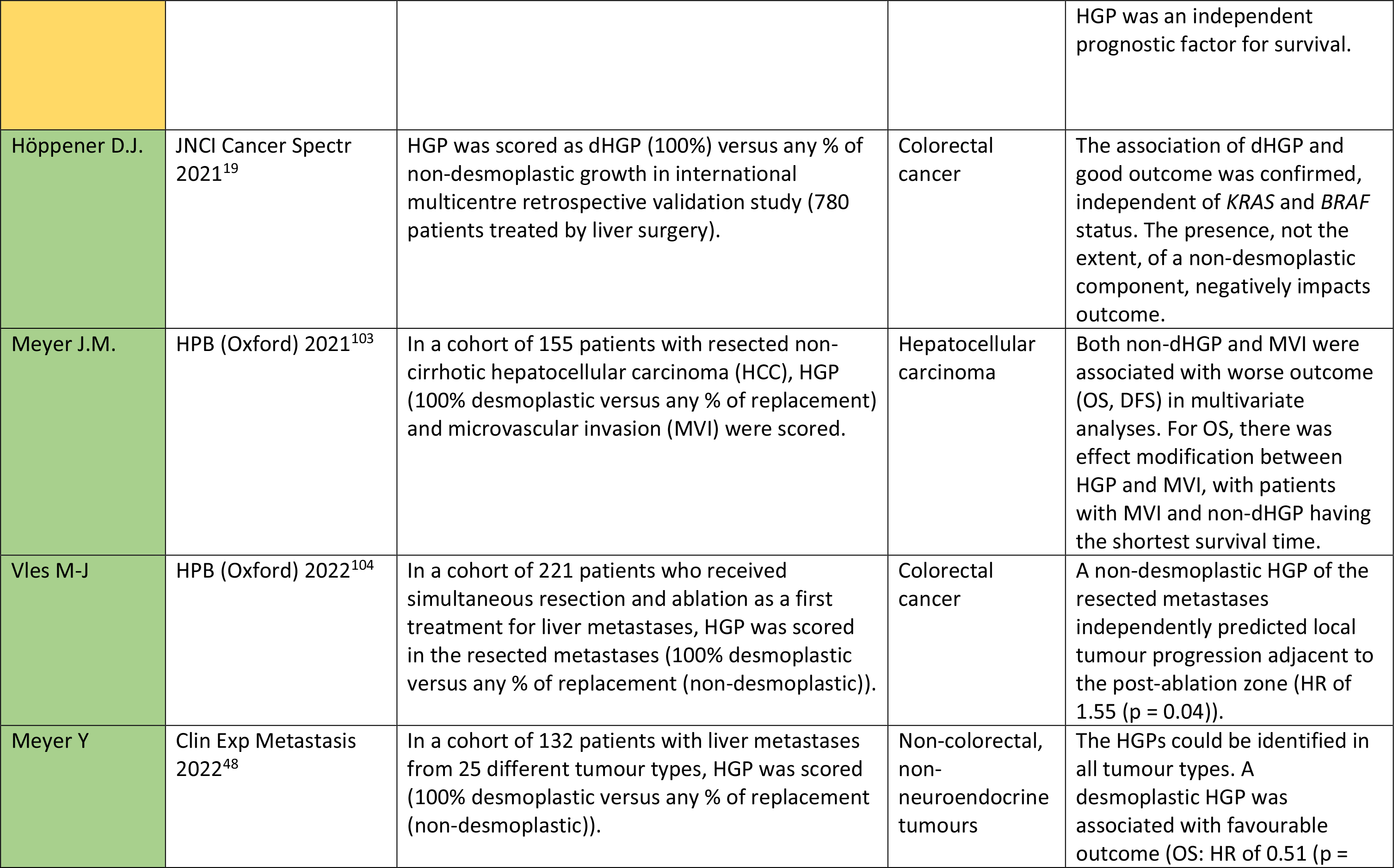
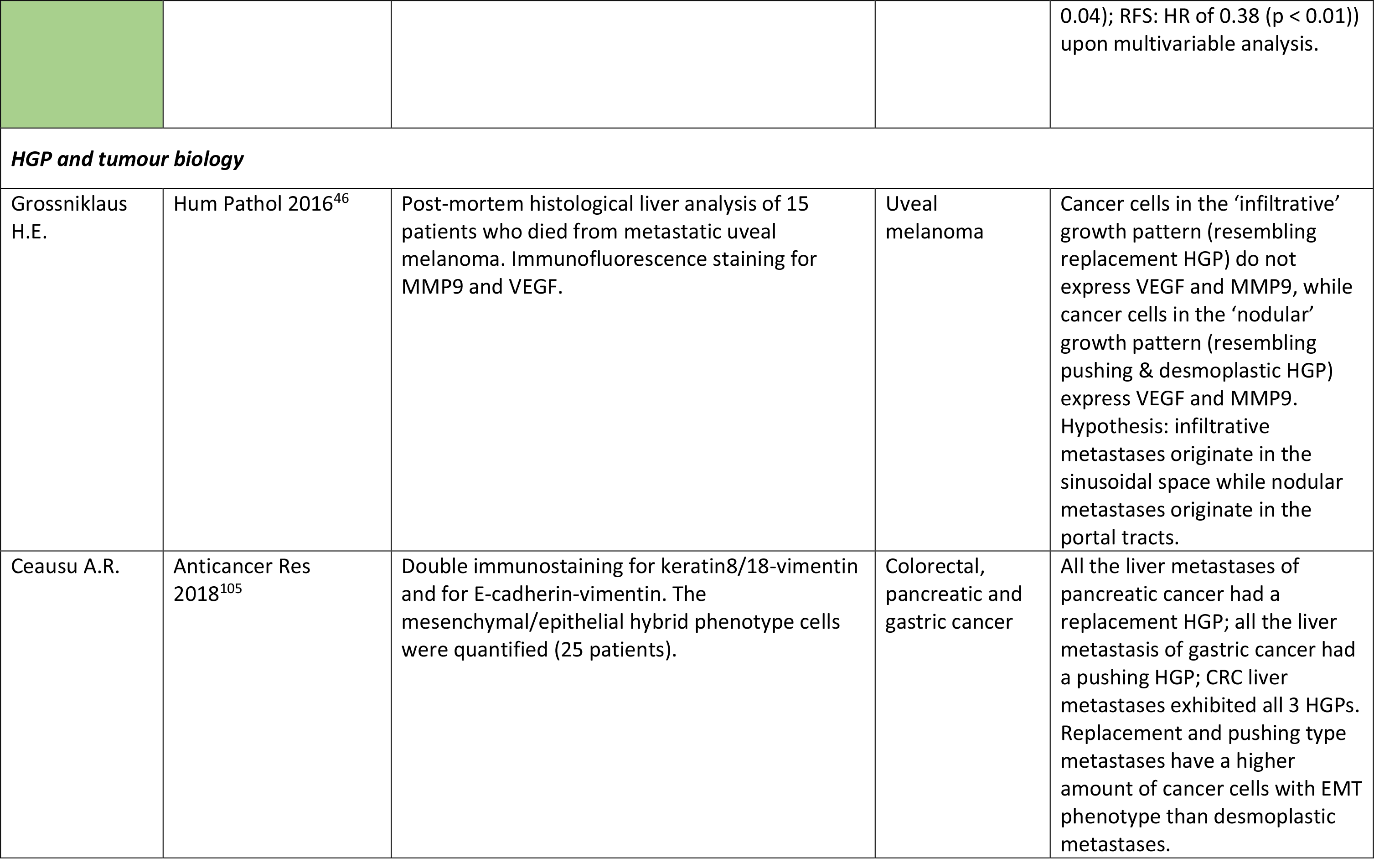
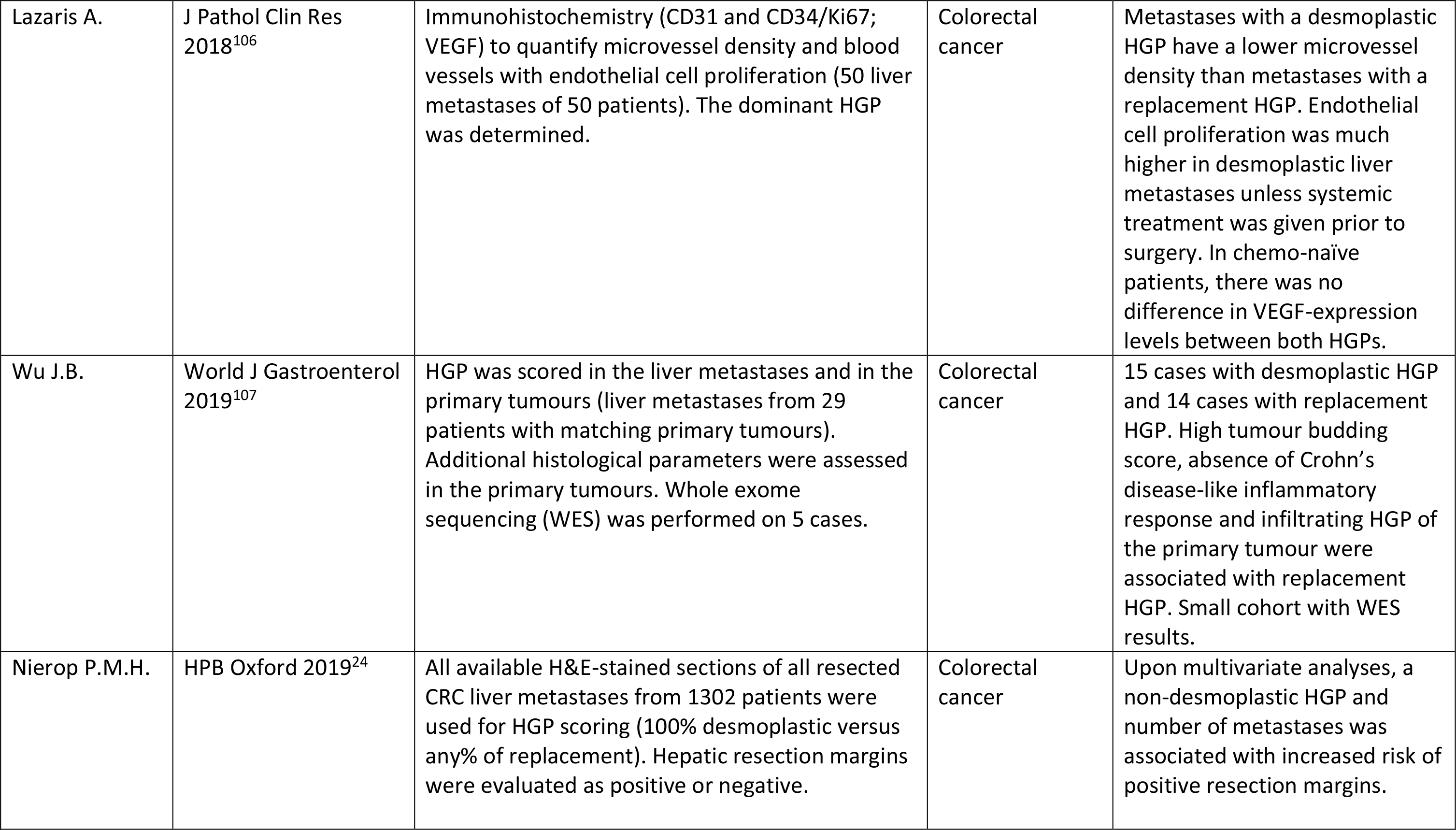
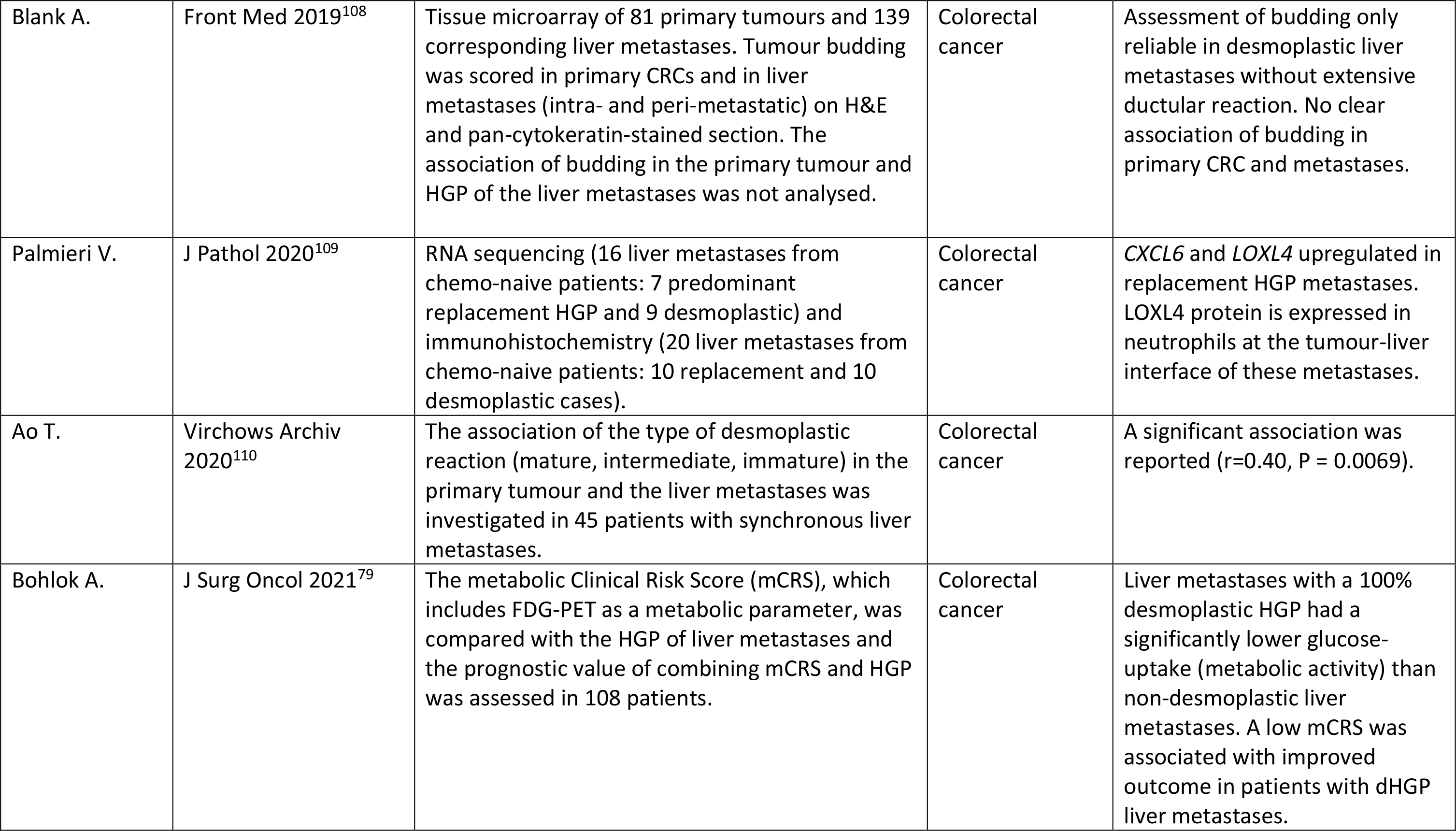
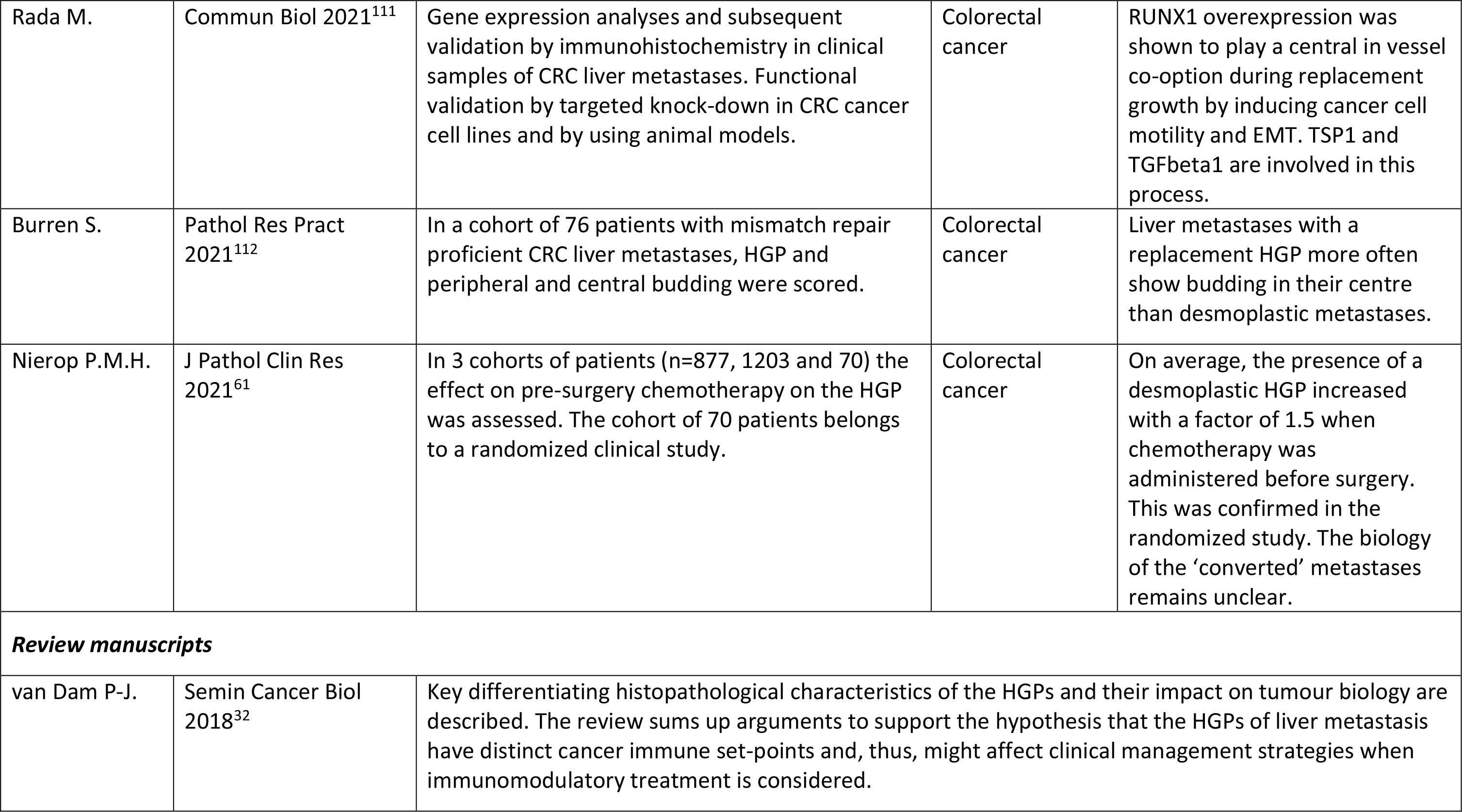
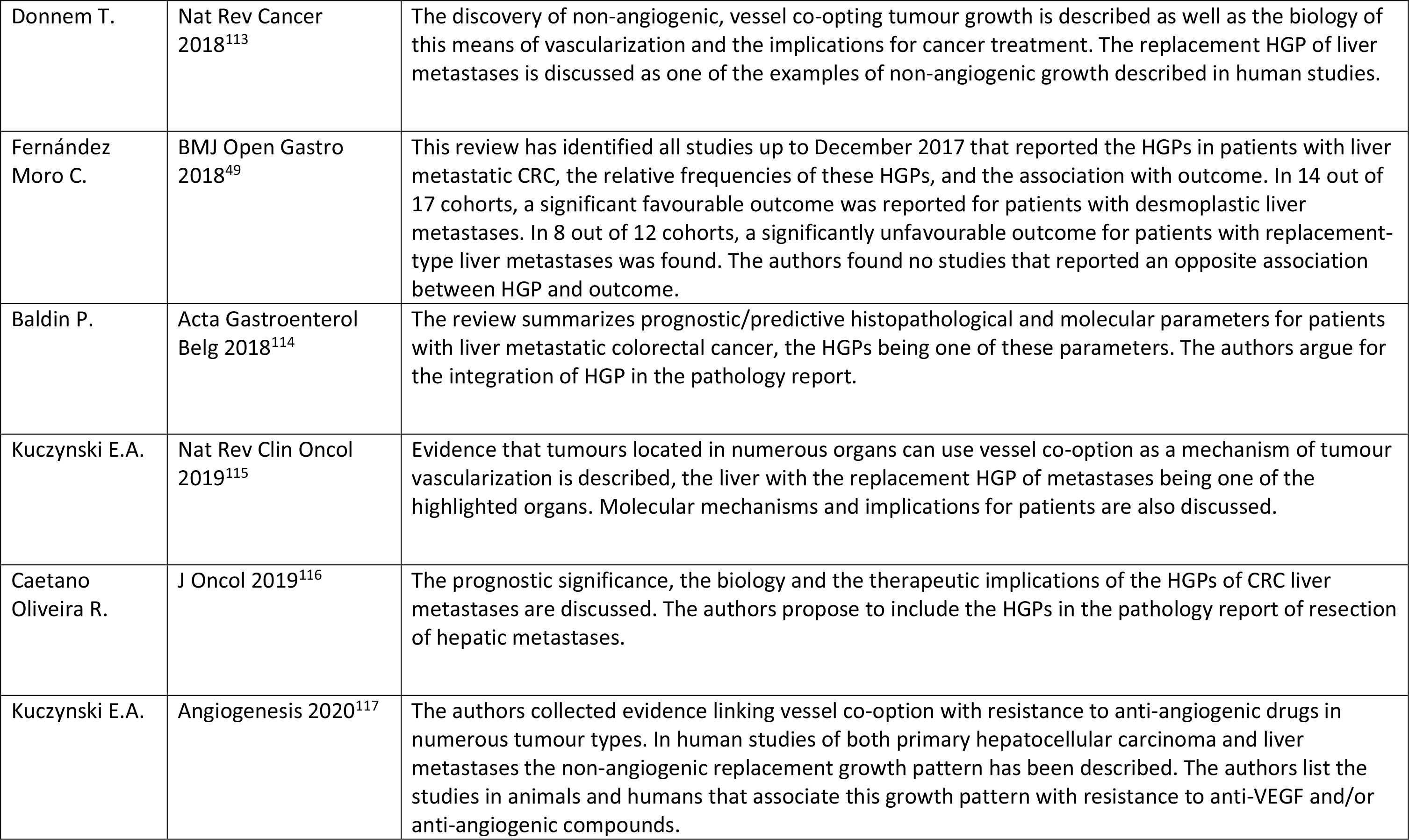
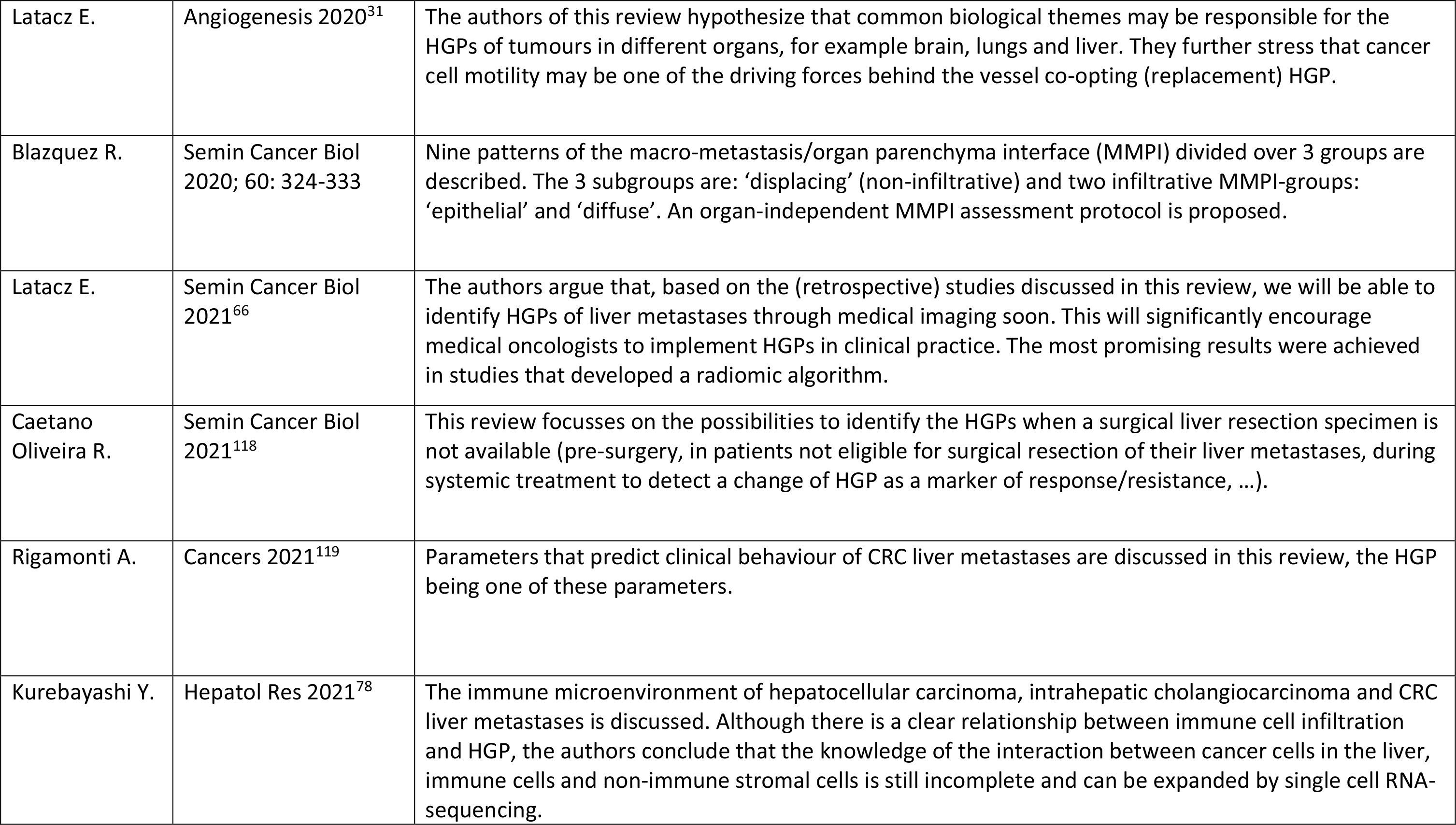
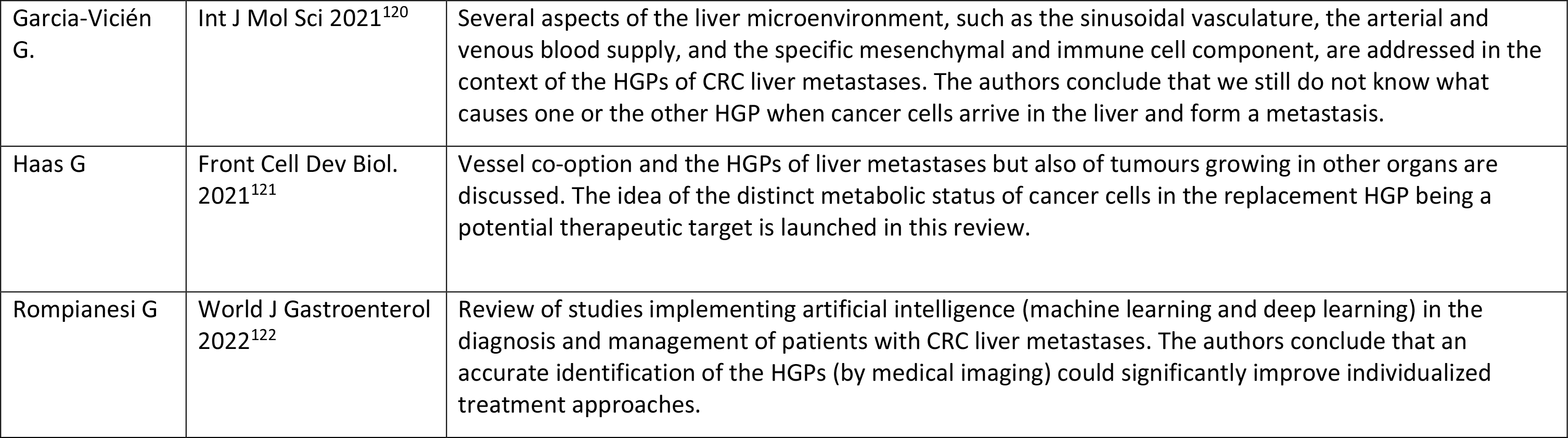
Overview of studies on the histopathological growth patterns of liver metastases, in addition to the studies listed in table 1. of the first guidelines paper (Van Dam et al 2017). Abbreviations: CGH = comparative genomic hybridization; CRC = colorectal cancer; CRISPR = clustered regularly interspaced short palindromic repeats; CT = computed tomography; DFS = disease-free survival; dHGP = desmoplastic histopathological growth pattern; EMT = epithelial-to-mesenchymal transition; H&E = haematoxylin-and-eosin stained; HGP = histopathological growth pattern; HR = hazard’s ratio; MMP = matrix metalloprotease; MRI = magnetic resonance imaging; mCRS = metabolic clinical risk score; NOD scid = nonobese diabetic severe combined immunodeficiency; MMPI = macro-metastasis/organ parenchyma interface; OS = overall survival; PFS = progression-free survival; PDX = patient-derived xenograft; RFS = relapse free survival; rHGP = replacement histopathological growth pattern; ROI = region of interest; TLI = tumour-liver interface; VEGF = vascular endothelial growth factor; WES = whole exome sequencing.

| First author | Reference | Methodology | Tumour type | Main findings |
| --- | --- | --- | --- | --- |
| ***Animal models*** |  |  |  |  |
| Alzubi M.A. | Clin Exp Metastasis 2019<sup>11</sup> | Portal vein injection of cancer cells of PDX mammary tumours of 14 patients in NOD *scid gamma* mice. | Breast cancer | HGPs could be assessed in six PDX models: replacement, desmoplastic and pushing HGPs were identified. |
| Piquet L. | Cancers 2019<sup>12</sup> | Co-inoculation into the spleen of human primary hepatic stellate cells and 5 human uveal melanoma cell lines in NOD *scid gamma* or NOD CRISPR *Prkdc Il2r gamma* mice. | Uveal Melanoma | Desmoplastic, replacement and mixed liver metastases were observed.<br>The HGP was not altered by co-inoculation of stellate cells (figure 5A and table 2 of the publication) |
| Vlachogiannis G. | Science 2018<sup>13</sup> | A biobank of patient-derived organoids and xenografts was constructed (110 fresh biopsies from 71 patients enrolled in four prospective phase 1/2 clinical trials were processed) | Colorectal and gastro-oesophageal cancer | A predominance of replacement HGP was observed in xenografts from resistant patient, whereas tumours established from sensitive patient showed a prevalence of desmoplastic and pushing HGPs. |
| Ibrahim N.S. | Cancers 2020<sup>14</sup> | Intra-splenic injection of MC-38 mouse CRC cell line in inducible Ang1 knock-out C57BL/6 mice. | Colorectal cancer | Replacement HGP liver metastases in control mice and desmoplastic HGP liver metastases in Ang1 knock-out condition. |
| Masaki S. | Int J Exp Pathol 2020<sup>15</sup> | Fatty liver conditions were induced in BALB/c mice. CT26 cells were injected into the liver. | Colorectal cancer | Tumours in control mice showed encapsulated growth patterns, while tumours in fatty livers showed invasive growth without encapsulation. |
| Tabariès S. | Commun Biol 2021<sup>16</sup> | Intrahepatic transplantation of patient liver metastasis tissue fragments in Scid-beige mice. Expression profiles of claudins were compared between dHGP and rHGP in PDXs and in liver metastases of patients. | Colorectal cancer | Liver metastases in mice express the HGP of the liver metastases of the patient-donor. Claudin-2 in patient-derived extracellular vesicles may be a marker of rHGP. |
| Bartlett A. | Cancers 2021<sup>17</sup> | Portal vein injection of D2OR, a low metastatic mouse mammary tumour cell line in nulliparous BALB/c immune competent mice and weaning-induced liver involution mice. | Breast cancer | The post-weaning liver is in an immune suppressed state with increased tumour incidence and multiplicity. A greater diversity of HGPs was noted in the post-weaning mice, consistent with the liver microenvironment dictating tumour histology. |
| **Immune contexture** (also: Watanabe K. in 'HGP scoring methodology' section) |  |  |  |  |
| Stremitzer S. | Br J Cancer 2020<sup>2</sup> | The immune phenotype of liver metastases was scored based on the distribution of CD8-immunostained cytotoxic T-lymphocytes as 'desert', 'excluded' (together 'non-inflamed') and 'inflamed' (81 patients). Bevacizumab-based chemotherapy was administered to all patients before partial liver resection. | Colorectal cancer | The inflamed immune phenotype was associated with the desmoplastic HGP and was associated with improved RFS and OS in univariable, not multivariable analyses. |
| Liang J. | Cancer Immunol Immunother 2020<sup>3</sup> | The immunoscore was calculated according to the densities of immunostained CD3 + and CD8 + cells (166 patients). One immunoscore per patient was calculated based on assessments in the tumour centre and in the invasive margin. | Colorectal cancer | A high immunoscore was more often encountered in liver metastases with a desmoplastic HGP than with a replacement HGP. A combined risk score (HGP, immunoscore and clinical risk score) was developed and a 90% 5-year OS rate was observed for patients in the low-risk group (30% of the patients). |
| Höppener D.J. | Br J Cancer 2020<sup>4</sup> | The immune contexture of resected liver metastases was analysed in 3 cohort of chemo-naive patients (117, 34 and 79 patients, respectively) with immunohistochemistry (semi-quantitative grading, quantitative digital image analysis) and flow cytometry. The 100% desmoplastic HGP cut off was applied. | Colorectal cancer | An increased immune infiltrate is associated with the desmoplastic HGP, both surrounding and in the metastases. Intra-epithelial CD8+ cells were also increased in the desmoplastic HGP. |
| Messaoudi N. | Br J Cancer. 2022<sup>6</sup> | Immunohistochemistry and automated quantitative analysis on tissue microarray (176 patients) of CD3, MHC-I and CD73. Liver metastases were categorized according to the dominant HGP and according to the 100% desmoplastic HGP cut off. | Colorectal cancer | Desmoplastic liver metastases were more infiltrated by CD3 + cells, expressed lower levels of MHC-I, and similar levels of CD73. Elevated CD73 expression was associated with a worse outcome of patients with desmoplastic HGP liver metastases. Low MHC-I expression in patients with replacement-type metastases improved outcome. |
| Garcia-Vicién G | Cancers 2022<sup>89</sup> | The spatial distribution of lymphocytic infiltrates in CRC liver metastases was explored in the context of the HGPs by multiplex immunofluorescence staining and digital image analysis in a cohort of 22 resected metastases without pre-surgery chemotherapy. HGPs were scored following the previous guidelines. The desmoplastic rim was excluded from the invasive margin for lymphocyte counting ('Measure B'). | Colorectal cancer | The number of CD8-positive cells at the invasive margin was independent of the HGP. In non-desmoplastic metastases, the cytotoxic T cells did not enter the tumour cell nests and CD4-positive cells were more abundant at the invasive margin than in desmoplastic lesions. |
| **HGP scoring methodology** |  |  |  |  |
| Höppener D.J. | Clin Exp Metastasis 2019<sup>39</sup> | Within and between metastasis HGP concordance was analysed in 363 patients with 2 or more resected liver metastases. The association of diagnostic accuracy with number of sections and number of metastases evaluated was determined. Interobserver agreement of HGP scoring was assessed after training. The 100% desmoplastic HGP cut off was applied. | Colorectal cancer | Within metastasis concordance ranged from 93% to 96%. Between metastasis concordance was 90%. Diagnostic accuracy peaked at two sections and two metastases. After two training sessions, interobserver agreement had a kappa-value of more than 0.9. |
| Watanabe K. | Cancer Med 2020<sup>5</sup> | Biopsies of liver metastases of 107 patients with pancreatic cancer (21- or 18-gauge needle) were used for HGP assessment. The dominant HGP was determined. If a HGP was present in more than | Pancreatic cancer | Of 279 patients, 107 patients had a biopsy that contained the tumour-liver interface. HGP had a homogenous expression in 13/14 patients. Disease control |
|  |  | 80% of the interface, the HGP was called 'homogenous' (analysis in 14 patients). |  | rate as well as overall survival rate were lower in the replacement HGP group. The replacement HGP biopsies showed less inflammation (H&E) and contained less CD8 + cells than the other biopsies. |
| Szczepanski J. | Am J Surg Pathol 2021<sup>90</sup> | The HGP was scored in biopsies of liver metastases of melanoma (n=30; 22 skin melanomas; 6 ocular melanomas; 2 unknown origin). | Melanoma | In 8/30 (4 ocular, 4 skin, 27%) melanoma liver metastases, a sinusoidal HGP was seen. In none of the 96 metastases of breast, colon, pancreaticobiliary cancer and neuroendocrine tumours this HGP was encountered. |
| **Medical imaging** |  |  |  |  |
| Gulia S. | BMJ Case Rep 2016<sup>7</sup> | A case report of a radiographically occult liver metastasis leading to liver failure is presented. | Breast cancer | A biopsy established the diagnosis of a liver metastasis with intra-sinusoidal growth pattern. |
| Cheng J. | Ann Surg Oncol 2019<sup>8</sup> | A radiomic algorithm was developed to identify the dominant HGPs of liver metastases by computed tomography (CT) imaging. Pre- and post-contrast as well as arterial and portal venous phase images (ROI: tumour-liver interface) contributed to the algorithm (126 metastases of 94 chemo-naïve patients - variety of scanners but standardized acquisition protocol and use of contrast agent). | Colorectal cancer | The dominant HGP of the liver metastases could be predicted with 65% sensitivity and 92% specificity (accuracy of 77%). A decisive feature used by the algorithm is the presence (desmoplastic) or absence (replacement) of peripheral rim enhancement in the portal- venous phase. No clinical or qualitative image data were used by the algorithm. |
| Han Y. | Front Oncol 2020<sup>9</sup> | A radiomic algorithm was developed to identify the dominant HGP of liver metastases by magnetic resonance imaging (MRI). (ROI: tumour-liver interface (TLI) - 182 liver metastases (107 chemo-naive patients)) | Colorectal cancer | The radiomic algorithm that best predicted the dominant HGP was based on quantitative features extracted from the TLI combined with clinical data and a qualitative image feature ('lobular margin') (79% accuracy, 100% sensitivity, 35% specificity). The desmoplastic HGP had more heterogeneous radiomic features than the replacement HGP. |
| Starmans M.P.A. | Clin Exp Metastasis 2021<sup>10</sup> | A radiomic algorithm was developed to distinguish liver metastases with 100% desmoplastic HGP from liver metastases with 100% replacement HGP by CT imaging (76 chemo-naive patients with 93 metastases). | Colorectal cancer | Despite the use of only portal venous phase contrast-enhanced images, variations in lesion segmentation and acquisition protocols, accuracy was 65%, sensitivity 72% and specificity 58%. |
| Wei S. | Eur J Radiol 2021<sup>67</sup> | The CT image-based radiomics algorithm to identify the dominant HGP developed in Cheng et al. (2019) was used to predict response to bevacizumab-chemotherapy in 119 patients (346 lesions) with unresectable CRC liver metastases. | Colorectal cancer | AUC for predicting early response was 0.72. The radiomics algorithm-derived HGP was the only independent predictor of 1-year PFS. |
| Li W.H. | Quant Imaging Med Surg 2022<sup>91</sup> | MRI features were used to predict the dominant HGP in 53 chemo-naïve patients. | Colorectal cancer | AUC for predicting the dominant HGP based on diameter difference between pre- and post-contrast images and rim enhancement was 0.83. |
| **HGP as biomarker** (HGP assessment *not according to guidelines*, *according to guidelines with dominant HGP as categories* and *according to guidelines with 100% desmoplastic HGP versus any percentage of replacement as categories*) |  |  |  |  |
| de Ridder J.A.M. | Ann Surg Onc 2015<sup>92</sup> | The presence/absence of a fibrous capsule was scored on H&E sections of resected liver metastases of 124 chemo-naïve patients with a solitary metastasis. The proportion of the tumour-liver interface with/without capsule was not reported. | Colorectal cancer | In univariable but not multivariable analysis, the presence of a fibrous capsule was associated with improved OS (109 months versus 57 months). |
| Serrablo A. | Eur J Surg Oncol 2016<sup>93</sup> | The presence/absence of a fibrous capsule with a thickness of at least 0,5mm in the entire tumour-liver interface was assessed on H&E sections (147 patients: 74/147 with pre-surgery systemic treatment) | Colorectal cancer | The capsule was present in 17% of the patients, independent of pre-surgery treatment status, and did not have an impact on survival. |
| Fonseca G.M. | J Surg Oncol 2018<sup>94</sup> | Tumour border pattern was scored according to the Jass classification (infiltrative, expansive). A fibrous capsule was scored as being absent or present. A single tissue block of the largest metastasis was selected for each patient (229 patients, all with peri-operative systemic treatment). | Colorectal cancer | Both absence of a fibrous capsule (75% of patients) and infiltrative growth (74% of patients) were associated with shorter OS and DFS in multivariable and/or univariable analyses. Both parameters were also associated with hepatic recurrence. |
| Cremolini C. | Br J Cancer 2018<sup>95</sup> | HGPs were scored according to the international guidelines. The effect of the HGPs on OS and DFS was investigated in a cohort of patients with liver metastases and with chemotherapy combined with either bevacizumab or cetuximab prior to surgery (159 patients). | Colorectal cancer | There was no effect of HGP on OS or DFS. An important remark is that the proportion of patients with liver metastases with a dominant pushing HGP was much higher than reported in most other studies (41%). |
| Falcao D. | Eur J Surg Oncol 2018<sup>96</sup> | HGPs of liver metastases were scored in 110 patients of which 52 patients received pre-surgery chemotherapy. A mixed HGP was identified when more than one HGP was expressed by the metastases and each HGP was present in at least 25% of the interface. | Colorectal cancer | The pushing HGP was independently associated with worse OS and DFS. An important remark is that the proportion of patients with liver metastases with a pushing HGP was much higher than reported in most other studies (30%). |
| Barnhill R. | J Pathol Clin Res 2018<sup>44</sup> | The dominant HGP was scored according to the international guidelines. Gene alterations were assessed by array CGH (41 liver metastases originating from 41 patients). | Uveal melanoma | Dominant replacement HGP metastases were present in 73% of patients (27%: desmoplastic HGP). On multivariate analysis, only HGP and resection status predicted OS (HR of 6.5 for replacement HGP). |
| Galjart B. | Angiogenesis 2019<sup>18</sup> | HGPs were scored according to the international guidelines but patients were categorized as having 100% desmoplastic (dHGP) liver metastases or not (non-dHGP) (732 patients of which 367 chemo-naïve before surgery) | Colorectal cancer | About 20% of the patients with surgical resection of CRC liver metastases ended up in 100% dHGP group. This was associated with an outstanding outcome, especially in the chemo-naïve group (78% with at least 5 years OS) |
| Nierop P.M.H. | Clin Exp Metastasis 2019<sup>23</sup> | HGP was scored as 100% desmoplastic (dHGP) versus non-dHGP in 690 patients free of disease after first resection of liver metastases of which 492 developed recurrent disease. | Colorectal cancer | Patients with dHGP at first partial hepatectomy were more often treated with curative intent and more often had recurrences salvageable by local treatment modalities. |
| Ao T | Virchows Arch 2019<sup>97</sup> | The desmoplastic reaction in and around liver metastases was scored as mature/intermediate (mature collagen fibers and keloid-like collagen) and immature (myxoid collagen present) in 204 patients with resected liver metastases of which 78 had received preoperative chemotherapy | Colorectal cancer | The type of desmoplastic reaction was independently associated with outcome with 65% 5-years OS in the mature /intermediate group versus 35% in the immature group. |
| Barnhill R | J Pathol Clin Res 2020<sup>45</sup> | HGP was scored as 100% desmoplastic (dHGP) versus 'any % of replacement' (any rHGP) (43 liver metastases from 42 patients). | Cutaneous melanoma | Multivariate analysis demonstrated that only HGP was associated with OS after resection of the liver metastases (HR for 'any rHGP' of 3.8). |
| Zhang Y | J Oncol 2020<sup>98</sup> | Encapsulation of hepatocellular carcinoma was assessed in 188 patients (method not specified). | Hepatocellular carcinoma | In multivariate analyses, the presence of a capsule was associated with improved DFS and OS (HR of 0.60 and 0.51, respectively). |
| Buisman F.E. | Clin Exp Metastasis 2020<sup>25</sup> | HGP was scored as 100% desmoplastic (dHGP) versus non-dHGP in resected liver metastases of 1236 patients of whom 656 received pre-operative chemotherapy. | Colorectal cancer | Adjuvant chemotherapy improved OS and DFS only in patients with non-dHGP liver metastases who did not receive pre-operative chemotherapy (HR of 0.52 and 0.71, respectively) |
| Baldin P. | J Pathol Clin Res 2021<sup>99</sup> | A pathological score (combining 'more than 3 lesions', 'R1 positive margin', 'non-100% desmoplastic HGP', 'steatohepatitis') and the consensus Immunoscore were tested for effect on outcome in 221 patients (85% received pre-operative chemotherapy; 582 liver metastases). Remark: per patient HGP used for outcome analysis was determined by selecting the 'worst' metastasis: pure replacement or mixed HGP. | Colorectal cancer | Non-desmoplastic HGP predicted shorter time to relapse in univariate and multivariate analyses (HRs 1,84 en 1,75, respectively). Patients with a favourable pathological score and a high immunoscore had the lowest risk of relapse (about 60% 5 yrs survival). |
| Temido M. | Cancer Management and Research 2020<sup>100</sup> | HGP was scored as dHGP (100%) versus any % of non-desmoplastic growth (17 patients). | Gastric cancer | dHGP was independently associated with improved OS (HR=0.1, p=0.02). |
| Bohlok A. | NPJ Breast Cancer 2020<sup>42</sup> | HGP was scored as 100% replacement (rHGP) versus 'any % of desmoplastic (any dHGP) (36 patients (11 patients with multiple metastases)). | Breast cancer | Any dHGP was independently associated with better PFS after liver surgery when compared with rHGP (HR=0.24, p = 0.009). All patients with rHGP relapsed within 20 months after liver surgery. |
| Jayme V.R. | Ann Surg Oncol 2021<sup>101</sup> | Tumour growth pattern of CRC liver metastases was defined as 'infiltrative' or 'pushing', according to Jass J.R. in 182 patients who underwent partial hepatectomy. | Colorectal cancer | Patients with infiltrative liver metastases (68% of patients) had worse OS and DFS, independent of surgical margin width. |
| Zhang Y.L. | Zhonghua Bing Li Xue Za Zhi 2021<sup>102</sup> | The dominant HGP was scored according to the international guidelines in 80 patients with partial hepatectomy. | Colorectal cancer | The 3-year PFS of patients with dHGP liver metastases (54%) was significantly longer compared with rHGP (40%). |
|  |  |  |  | HGP was an independent prognostic factor for survival. |
| Höppener D.J. | JNCI Cancer Spectr 2021<sup>19</sup> | HGP was scored as dHGP (100%) versus any % of non-desmoplastic growth in international multicentre retrospective validation study (780 patients treated by liver surgery). | Colorectal cancer | The association of dHGP and good outcome was confirmed, independent of *KRAS* and *BRAF* status. The presence, not the extent, of a non-desmoplastic component, negatively impacts outcome. |
| Meyer J.M. | HPB (Oxford) 2021<sup>103</sup> | In a cohort of 155 patients with resected non-cirrhotic hepatocellular carcinoma (HCC), HGP (100% desmoplastic versus any % of replacement) and microvascular invasion (MVI) were scored. | Hepatocellular carcinoma | Both non-dHGP and MVI were associated with worse outcome (OS, DFS) in multivariate analyses. For OS, there was effect modification between HGP and MVI, with patients with MVI and non-dHGP having the shortest survival time. |
| Vles M-J | HPB (Oxford) 2022<sup>104</sup> | In a cohort of 221 patients who received simultaneous resection and ablation as a first treatment for liver metastases, HGP was scored in the resected metastases (100% desmoplastic versus any % of replacement (non-desmoplastic)). | Colorectal cancer | A non-desmoplastic HGP of the resected metastases independently predicted local tumour progression adjacent to the post-ablation zone (HR of 1.55 (p = 0.04)). |
| Meyer Y | Clin Exp Metastasis 2022<sup>48</sup> | In a cohort of 132 patients with liver metastases from 25 different tumour types, HGP was scored (100% desmoplastic versus any % of replacement (non-desmoplastic)). | Non-colorectal, non-neuroendocrine tumours | The HGPs could be identified in all tumour types. A desmoplastic HGP was associated with favourable outcome (OS: HR of 0.51 (p = |
|  |  |  |  | 0.04); RFS: HR of 0.38 (p < 0.01)) upon multivariable analysis. |
| **HGP and tumour biology** |  |  |  |  |
| Grossniklaus H.E. | Hum Pathol 2016<sup>46</sup> | Post-mortem histological liver analysis of 15 patients who died from metastatic uveal melanoma. Immunofluorescence staining for MMP9 and VEGF. | Uveal melanoma | Cancer cells in the 'infiltrative' growth pattern (resembling replacement HGP) do not express VEGF and MMP9, while cancer cells in the 'nodular' growth pattern (resembling pushing & desmoplastic HGP) express VEGF and MMP9. Hypothesis: infiltrative metastases originate in the sinusoidal space while nodular metastases originate in the portal tracts. |
| Ceausu A.R. | Anticancer Res 2018<sup>105</sup> | Double immunostaining for keratin8/18-vimentin and for E-cadherin-vimentin. The mesenchymal/epithelial hybrid phenotype cells were quantified (25 patients). | Colorectal, pancreatic and gastric cancer | All the liver metastases of pancreatic cancer had a replacement HGP; all the liver metastasis of gastric cancer had a pushing HGP; CRC liver metastases exhibited all 3 HGPs. Replacement and pushing type metastases have a higher amount of cancer cells with EMT phenotype than desmoplastic metastases. |
| Lazaris A. | J Pathol Clin Res 2018<sup>106</sup> | Immunohistochemistry (CD31 and CD34/Ki67; VEGF) to quantify microvessel density and blood vessels with endothelial cell proliferation (50 liver metastases of 50 patients). The dominant HGP was determined. | Colorectal cancer | Metastases with a desmoplastic HGP have a lower microvessel density than metastases with a replacement HGP. Endothelial cell proliferation was much higher in desmoplastic liver metastases unless systemic treatment was given prior to surgery. In chemo-naïve patients, there was no difference in VEGF-expression levels between both HGPs. |
| Wu J.B. | World J Gastroenterol 2019<sup>107</sup> | HGP was scored in the liver metastases and in the primary tumours (liver metastases from 29 patients with matching primary tumours). Additional histological parameters were assessed in the primary tumours. Whole exome sequencing (WES) was performed on 5 cases. | Colorectal cancer | 15 cases with desmoplastic HGP and 14 cases with replacement HGP. High tumour budding score, absence of Crohn's disease-like inflammatory response and infiltrating HGP of the primary tumour were associated with replacement HGP. Small cohort with WES results. |
| Nierop P.M.H. | HPB Oxford 2019<sup>24</sup> | All available H&E-stained sections of all resected CRC liver metastases from 1302 patients were used for HGP scoring (100% desmoplastic versus any% of replacement). Hepatic resection margins were evaluated as positive or negative. | Colorectal cancer | Upon multivariate analyses, a non-desmoplastic HGP and number of metastases was associated with increased risk of positive resection margins. |
| Blank A. | Front Med 2019<sup>108</sup> | Tissue microarray of 81 primary tumours and 139 corresponding liver metastases. Tumour budding was scored in primary CRCs and in liver metastases (intra- and peri-metastatic) on H&E and pan-cytokeratin-stained section. The association of budding in the primary tumour and HGP of the liver metastases was not analysed. | Colorectal cancer | Assessment of budding only reliable in desmoplastic liver metastases without extensive ductular reaction. No clear association of budding in primary CRC and metastases. |
| Palmieri V. | J Pathol 2020<sup>109</sup> | RNA sequencing (16 liver metastases from chemo-naïve patients: 7 predominant replacement HGP and 9 desmoplastic) and immunohistochemistry (20 liver metastases from chemo-naïve patients: 10 replacement and 10 desmoplastic cases). | Colorectal cancer | *CXCL6* and *LOXL4* upregulated in replacement HGP metastases. *LOXL4* protein is expressed in neutrophils at the tumour-liver interface of these metastases. |
| Ao T. | Virchows Archiv 2020<sup>110</sup> | The association of the type of desmoplastic reaction (mature, intermediate, immature) in the primary tumour and the liver metastases was investigated in 45 patients with synchronous liver metastases. | Colorectal cancer | A significant association was reported ($r=0.40$, $P = 0.0069$). |
| Bohlok A. | J Surg Oncol 2021<sup>79</sup> | The metabolic Clinical Risk Score (mCRS), which includes FDG-PET as a metabolic parameter, was compared with the HGP of liver metastases and the prognostic value of combining mCRS and HGP was assessed in 108 patients. | Colorectal cancer | Liver metastases with a 100% desmoplastic HGP had a significantly lower glucose-uptake (metabolic activity) than non-desmoplastic liver metastases. A low mCRS was associated with improved outcome in patients with dHGP liver metastases. |
| Rada M. | Commun Biol 2021<sup>111</sup> | Gene expression analyses and subsequent validation by immunohistochemistry in clinical samples of CRC liver metastases. Functional validation by targeted knock-down in CRC cancer cell lines and by using animal models. | Colorectal cancer | RUNX1 overexpression was shown to play a central in vessel co-option during replacement growth by inducing cancer cell motility and EMT. TSP1 and TGFbeta1 are involved in this process. |
| Burren S. | Pathol Res Pract 2021<sup>112</sup> | In a cohort of 76 patients with mismatch repair proficient CRC liver metastases, HGP and peripheral and central budding were scored. | Colorectal cancer | Liver metastases with a replacement HGP more often show budding in their centre than desmoplastic metastases. |
| Nierop P.M.H. | J Pathol Clin Res 2021<sup>61</sup> | In 3 cohorts of patients (n=877, 1203 and 70) the effect on pre-surgery chemotherapy on the HGP was assessed. The cohort of 70 patients belongs to a randomized clinical study. | Colorectal cancer | On average, the presence of a desmoplastic HGP increased with a factor of 1.5 when chemotherapy was administered before surgery. This was confirmed in the randomized study. The biology of the 'converted' metastases remains unclear. |
| ***Review manuscripts*** |  |  |  |  |
| van Dam P-J. | Semin Cancer Biol 2018<sup>32</sup> | Key differentiating histopathological characteristics of the HGPs and their impact on tumour biology are described. The review sums up arguments to support the hypothesis that the HGPs of liver metastasis have distinct cancer immune set-points and, thus, might affect clinical management strategies when immunomodulatory treatment is considered. |  |  |

|  |  |  |
| --- | --- | --- |
| Donnem T. | Nat Rev Cancer 2018<sup>113</sup> | The discovery of non-angiogenic, vessel co-opting tumour growth is described as well as the biology of this means of vascularization and the implications for cancer treatment. The replacement HGP of liver metastases is discussed as one of the examples of non-angiogenic growth described in human studies. |
| Fernández Moro C. | BMJ Open Gastro 2018<sup>49</sup> | This review has identified all studies up to December 2017 that reported the HGPs in patients with liver metastatic CRC, the relative frequencies of these HGPs, and the association with outcome. In 14 out of 17 cohorts, a significant favourable outcome was reported for patients with desmoplastic liver metastases. In 8 out of 12 cohorts, a significantly unfavourable outcome for patients with replacement-type liver metastases was found. The authors found no studies that reported an opposite association between HGP and outcome. |
| Baldin P. | Acta Gastroenterol Belg 2018<sup>114</sup> | The review summarizes prognostic/predictive histopathological and molecular parameters for patients with liver metastatic colorectal cancer, the HGPs being one of these parameters. The authors argue for the integration of HGP in the pathology report. |
| Kuczynski E.A. | Nat Rev Clin Oncol 2019<sup>115</sup> | Evidence that tumours located in numerous organs can use vessel co-option as a mechanism of tumour vascularization is described, the liver with the replacement HGP of metastases being one of the highlighted organs. Molecular mechanisms and implications for patients are also discussed. |
| Caetano Oliveira R. | J Oncol 2019<sup>116</sup> | The prognostic significance, the biology and the therapeutic implications of the HGPs of CRC liver metastases are discussed. The authors propose to include the HGPs in the pathology report of resection of hepatic metastases. |
| Kuczynski E.A. | Angiogenesis 2020<sup>117</sup> | The authors collected evidence linking vessel co-option with resistance to anti-angiogenic drugs in numerous tumour types. In human studies of both primary hepatocellular carcinoma and liver metastases the non-angiogenic replacement growth pattern has been described. The authors list the studies in animals and humans that associate this growth pattern with resistance to anti-VEGF and/or anti-angiogenic compounds. |
| Latacz E. | Angiogenesis 2020<sup>31</sup> | The authors of this review hypothesize that common biological themes may be responsible for the HGP of tumours in different organs, for example brain, lungs and liver. They further stress that cancer cell motility may be one of the driving forces behind the vessel co-opting (replacement) HGP. |
| Blazquez R. | Semin Cancer Biol 2020; 60: 324-333 | Nine patterns of the macro-metastasis/organ parenchyma interface (MMPI) divided over 3 groups are described. The 3 subgroups are: 'displacing' (non-infiltrative) and two infiltrative MMPI-groups: 'epithelial' and 'diffuse'. An organ-independent MMPI assessment protocol is proposed. |
| Latacz E. | Semin Cancer Biol 2021<sup>66</sup> | The authors argue that, based on the (retrospective) studies discussed in this review, we will be able to identify HGPs of liver metastases through medical imaging soon. This will significantly encourage medical oncologists to implement HGPs in clinical practice. The most promising results were achieved in studies that developed a radiomic algorithm. |
| Caetano Oliveira R. | Semin Cancer Biol 2021<sup>118</sup> | This review focusses on the possibilities to identify the HGPs when a surgical liver resection specimen is not available (pre-surgery, in patients not eligible for surgical resection of their liver metastases, during systemic treatment to detect a change of HGP as a marker of response/resistance, ...). |
| Rigamonti A. | Cancers 2021<sup>119</sup> | Parameters that predict clinical behaviour of CRC liver metastases are discussed in this review, the HGP being one of these parameters. |
| Kurebayashi Y. | Hepatol Res 2021<sup>78</sup> | The immune microenvironment of hepatocellular carcinoma, intrahepatic cholangiocarcinoma and CRC liver metastases is discussed. Although there is a clear relationship between immune cell infiltration and HGP, the authors conclude that the knowledge of the interaction between cancer cells in the liver, immune cells and non-immune stromal cells is still incomplete and can be expanded by single cell RNA-sequencing. |
| Garcia-Vicién G. | Int J Mol Sci 2021<sup>120</sup> | Several aspects of the liver microenvironment, such as the sinusoidal vasculature, the arterial and venous blood supply, and the specific mesenchymal and immune cell component, are addressed in the context of the HGPs of CRC liver metastases. The authors conclude that we still do not know what causes one or the other HGP when cancer cells arrive in the liver and form a metastasis. |
| Haas G | Front Cell Dev Biol. 2021<sup>121</sup> | Vessel co-option and the HGPs of liver metastases but also of tumours growing in other organs are discussed. The idea of the distinct metabolic status of cancer cells in the replacement HGP being a potential therapeutic target is launched in this review. |
| Rompianesi G | World J Gastroenterol 2022<sup>122</sup> | Review of studies implementing artificial intelligence (machine learning and deep learning) in the diagnosis and management of patients with CRC liver metastases. The authors conclude that an accurate identification of the HGPs (by medical imaging) could significantly improve individualized treatment approaches. |

Clinical and experimental studies have provided ample new information that warrants an updated, second version of the international guidelines for scoring the HGPs in the context of liver metastasis. The main goal of the guidelines is to incorporate these histological features into the clinical decision-making processes surrounding the treatment of patients with liver metastatic cancer. We therefore provide a detailed histopathological description of the growth patterns of liver metastases and propose an updated standard method for HGP assessment within liver metastases, including immunohistochemical staining as an aid to scoring HGPs. One of the important features of the new guidelines is a modified and clinically applicable cut-off for considering a colorectal cancer (CRC) liver metastasis (CRLM) as desmoplastic or non-desmoplastic. This change in cut-off is supported by retrospective studies with large cohorts of patients with liver metastatic CRC^18, 19^. In the new guidelines, we present a pooled analysis of previously published cohorts to demonstrate the improved prognostic value of this new cut-off recommendation. In addition, we propose hypotheses that could explain the transition from one HGP to another, based on comprehensive immunohistochemical analyses of both the tumour-liver interface and the centre of the metastases. We also speculate on molecular mechanisms that may underlie the biological differences of the growth patterns. Finally, we discuss exciting new research perspectives for the HGPs, including digital image processing techniques and deep learning methods for automated HGP scoring using digitized haematoxylin-and-eosin-stained (H&E-stained) tissue sections^20–22^.

## Methods

### Literature search

We performed a literature search for studies published since January 2015 that focused on the HGPs of liver metastases using the PubMed^R^ resource of the U.S. National Library of Medicine. The search terms were designed to find studies on the evaluation of the interface between liver metastases and the surrounding liver tissue, independent of the primary tumour type and the host species. Additional studies were found by manual cross-referencing. Ultimately, manuscripts were selected by three reviewers (EL, DJH and PV). Only manuscripts that were not already presented in Table 1 of the first consensus guidelines publication^1^ are discussed in the current overview table (Table 1).

### Evaluation of the HGP cut-off algorithms

To compare the prognostic value of different HGP cut-off algorithms, survival analyses were performed. The HGP and survival data used for these analyses have been previously published as separate cohorts and were pooled for the current analysis^1, 18, 23–25^. All available H&E-stained sections of all resected liver metastases for every patient included in this assessment were analysed according to the 2017 consensus guidelines^1^. The final HGP score per patient is the average of all metastases, independent of the size of the metastases or number of analysed tissue sections per metastasis. Data on overall and disease-free survival (OS, DFS, defined as the time between first liver metastasis resection and death or cancer recurrence, respectively) and HGP were available for 1931 patients: 903 patients underwent surgical resection (1998 - 2019) in the Erasmus MC Cancer Institute (Rotterdam, the Netherlands), 716 patients in the Memorial Sloan Kettering Cancer Center (New York, NY, USA), and 312 patients in the Radboud University Medical Centre (Nijmegen, the Netherlands). All patients treated with curative intent, who did not receive hepatic arterial infusion pump chemotherapy, and for whom H&E-stained sections were available, were included. Approval by the institutional ethical review boards was obtained in each individual centre separately.

### Immunohistochemistry

For immunohistochemistry with antibodies (clone; manufacturer’s code) directed at CK7 (RN7; NCL-L-CK7-560), CK18 (DC-10; NCL-CK18), CK19 (b170; NCL-CK19), CK20 (PW31; NCL-L- CK20-561), Caldesmon (H-CD; Dako-M3557), CD34 (QBEnd/10; Dako-M7165), CD146 (UMAB154; Origene-UM800051), NGFR (polyclonal; Atlas-HPA004765) and alpha-SMA (1a4; DAKO-M0851), formalin-fixed paraffin-embedded (FFPE) tissue representing the respective areas were cut to 4 µm thickness. All immunohistochemical stains were done on a Leica (Germany) BOND-MAX automated stainer as part of clinical routine at Karolinska University Hospital, Huddinge, Sweden. Pretreatment was done using Bond Epitope Retrieval Solution 2 EDTA (Leica) for 20 minutes. Immunohistochemistry for antibodies directed at melan-A (A103; Dako-M7196) was done on a Leica BOND-RX automated stainer at Institut Curie, Paris, France. Pretreatment was done using Bond Epitope Retrieval Solution 2 EDTA (Leica) for 20 minutes.

### Statistics

For the comparison of different cut-off algorithms, OS and DFS were estimated using the Kaplan-Meier method and reported as 5-year (%), 10-year (%) and median (months) survival including a corresponding 95% confidence interval (CI). Adjusted hazard ratios (HR) for OS and DFS are based on multivariable Cox proportional hazards regression models. All statistical analyses were performed with the R Project for Statistical Computing (version 4.0.2; https://www.r-project.org/).

## Results Guidelines

### Histopathological description of the growth patterns of liver metastases

Liver metastases can interact differently with the liver parenchyma as they colonise the liver, which is manifest histologically as one of several distinct growth patterns. These patterns can generally be identified by light microscopy in H&E-stained sections of FFPE tissue at the interface between the cancer cells and the liver parenchyma^26–30^. The key histopathological characteristics of the HGPs have been described in Table 2 of the first international consensus guidelines^1^ and remain valid in that form. An updated overview of the histology of the different HGPs is presented in Table 2 and in Figures 1A-K of the current scoring guidelines. The desmoplastic and the replacement HGPs are the most common patterns, based on recent studies that have used the 2017 consensus guidelines (Table 1). For example, either the desmoplastic or the replacement HGP was evident in 97.5% of the tumour-liver interface of all CRC liver metastases of 732 patients^18^, almost equally distributed between both HGPs. In the desmoplastic HGP, the cancer cells are separated from the surrounding liver parenchyma by a fibrotic rim. Often a dense infiltrate of immune cells is present at the transition between the liver parenchyma and the fibrous rim. Desmoplastic liver metastases frequently show glandular differentiation (when derived from an adenocarcinoma) and are vascularized by a process of angiogenesis^31^ (Figures 1A-C).

**Figure 1.**
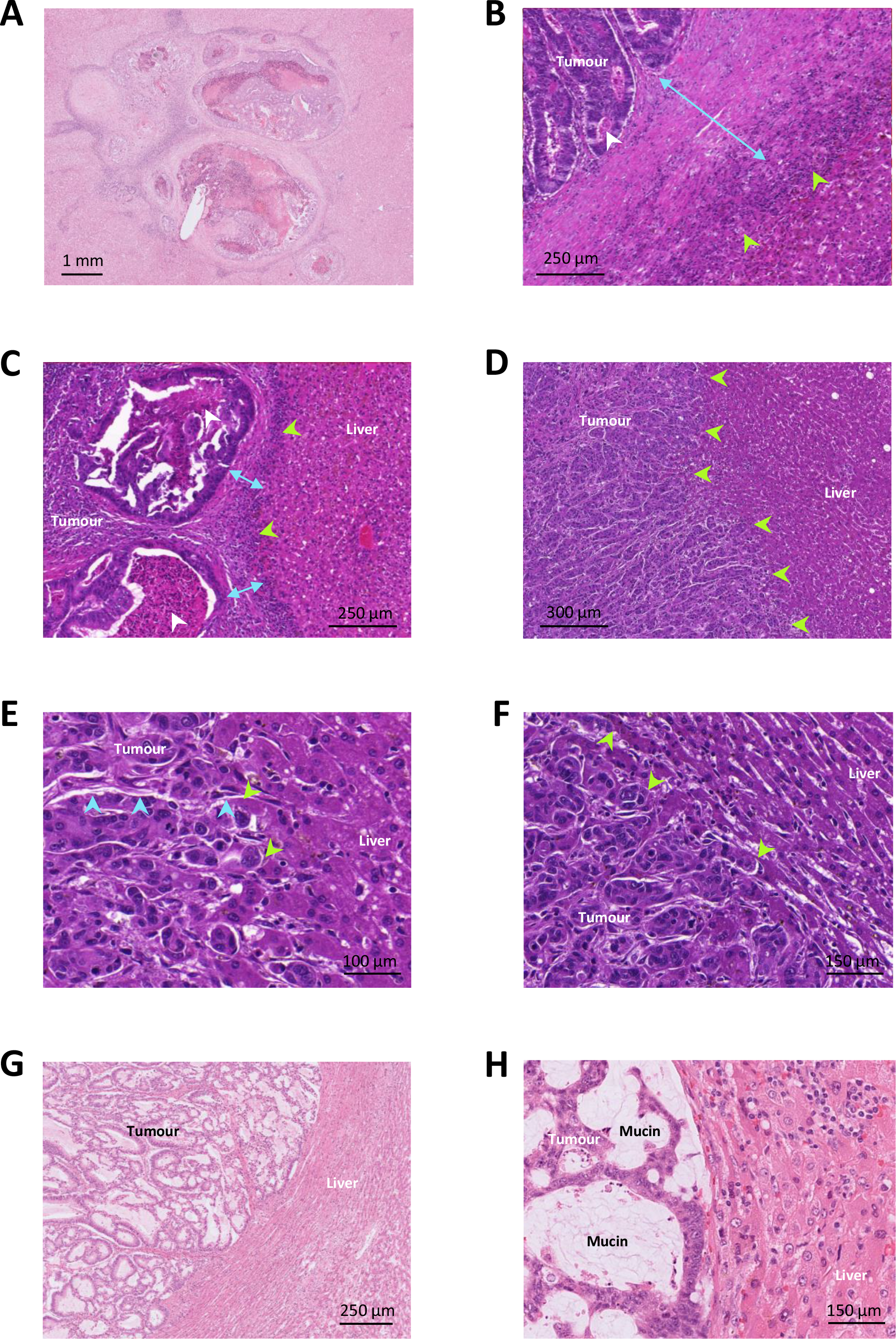
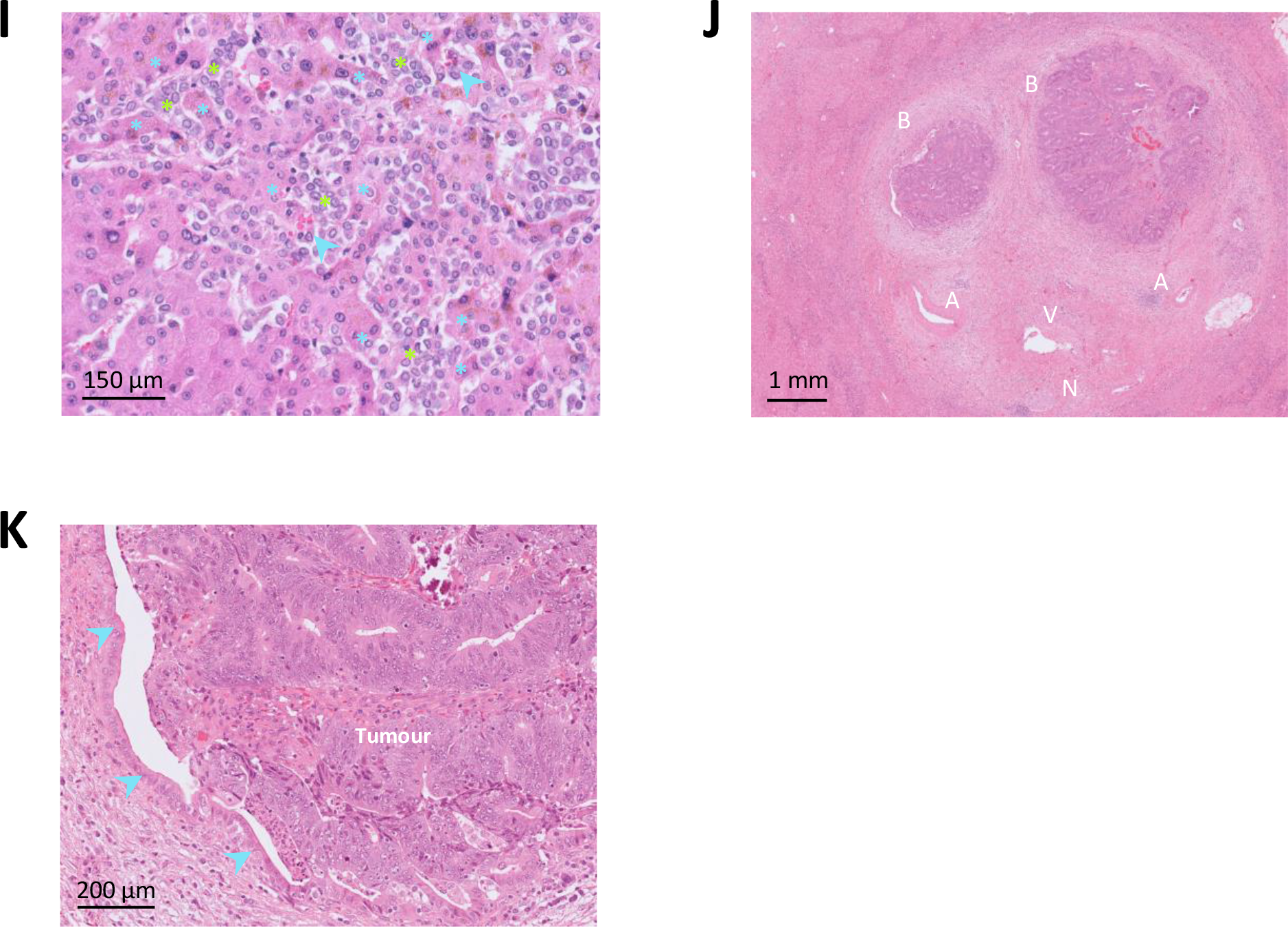
The histopathological growth patterns of liver metastases (H&E images). (A) Low magnification image of a CRC liver metastasis with a desmoplastic HGP. (B) & (C) Higher magnification of the tumour-liver interface of CRC liver metastases with a desmoplastic HGP. The blue double headed arrow indicates the desmoplastic rim that separates the carcinoma from the liver parenchyma. The green arrowheads indicate the immune cell infiltrate which is typically located at the transition between the desmoplastic rim and the liver parenchyma. The tumours show glandular differentiation and cell detritus in the lumina of these glandular structures, reminiscent of the histology of a primary CRC (white arrowheads). (D) Low magnification image of a CRC liver metastasis with a replacement HGP. The green arrowheads indicate the tumour-liver interface. There is no glandular differentiation: cancer cells from solid nests and trabeculae. (E) & (F) Higher magnification of the tumour-liver interface of CRC liver metastases with a replacement HGP. The green arrowheads indicate contact between cancer cells and hepatocytes. In (E), cancer cells form cell plates that are in continuity with the liver cell plates. A co-opted sinusoidal blood vessel is marked by the blue arrowheads. In (F), the liver cell plates are pushed aside but cancer cells are still in contact with hepatocytes while invading into these liver cell plates (green arrowheads). (G) Low magnification image of a CRC liver metastasis with a pushing HGP. (H) On higher magnification, a sharp tumour-liver interface is noticed without desmoplastic rim and without cancer cells invading into the liver parenchyma. Often metastases with a pushing HGP produce mucin, as shown in this example. (I) Lobular breast carcinoma liver metastasis with a sinusoidal HGP (autopsy case). Cancer cells are located within the lumen of sinusoidal blood vessels (green asterisks), in between liver cell plates (blue asterisks). Red blood cells are intermingled with the cancer cells (blue arrowheads). (J) Low magnification image of intrabiliary tumour growth (CRC) in a portal tract. The structures constituting a portal tract are present: artery branches (A), vein branch (V), nerve bundle (N), and branches of the bile duct (B), in this case filled with cancer cells. (K) Higher magnification of the left bile duct branch of image J. The normal bile duct epithelium (blue arrowheads) is still present but is replaced by cancer tissue that fills the lumen of the bile duct branch.

**Table 2.** Key histopathological characteristics of the growth patterns of liver metastases.

|  | **Desmoplastic** | **Replacement** | **Pushing** | **Sinusoidal** | **Portal**<br>**(including intrabiliary)** |
| --- | --- | --- | --- | --- | --- |
| General architecture | A desmoplastic rim separates metastatic tissue from liver tissue. | Cancer cell are arranged in plates in continuity with the hepatocyte plates. | Metastatic tissue pushes the liver tissue aside (without recognizable desmoplastic rim). | Cancer cells grow in the sinusoidal vessel lumina or in the Disse space, adjacent to the hepatocyte plates. | Metastatic tissue grows within portal tracts and septa and/or within the lumen of biliary branches |
| Liver architecture mimicry | - | + | - | + | n.a. |
| Liver stroma preserved | - | + | - | + | + |
| Contact of cancer cells with liver epithelial cells | Not with hepatocytes<br>Occasional contact with cholangiocytes of ductular reaction | + (hepatocytes) | - | - | With cholangiocytes if intrabiliary growth |
| Desmoplastic reaction around the metastasis | + | - | - | - | n.a. |
| Compression of liver cell plates | + | -/+ | + | - | n.a. |
| Contour | sharp | irregular | sharp | irregular | n.a. |
| Inflammatory cell infiltrate | ++ | +/- | +/- | +/- | n.a. |
| Proliferation of bile ducts (ductular reaction) | +/- | - | - | - | -/+ |
| Glandular differentiation (if adenocarcinoma) | + | - | + | - | + |
|  | Figures 1A-C | Figures 1D-F | Figures 1G & H | Figure 1I | Figures 1J & K |

In replacement-type liver metastases, cancer cells are in contact with the hepatocytes, they replace the hepatocytes, and, in the process, they co-opt the sinusoidal blood vessels of the liver. As a result, the tissue architecture of the metastases with this HGP mimics the tissue architecture of the liver, such that the metastatic cancer cells arrangement recapitulates ‘hepatic cell plates’ in between co-opted hepatic sinusoidal blood vessels. Typically, and based on observations done in carcinoma liver metastases, only a few immune cells are present at the tumour-liver interface and in the tumour centre^32^, although this is not a scoring criterion. Adenocarcinoma metastases with a replacement growth pattern do not usually show glandular differentiation at the tumour-liver interface (Figures 1D-F). Angiotropic extravascular migration has been observed in replacement-type liver metastases of melanoma^33^ (see section dedicated to angiotropic extravascular migration): single or small clusters of melanoma cells may extend along sinusoidal channels into the surrounding liver parenchyma with distances of several millimeters.

The pushing growth pattern is an uncommon pattern. For example, the pushing HGP was present in only 2.5% of the tumour-liver interface of all CRC liver metastases of 732 patients^18^. This growth pattern is characterized by cancer cells that appear to push away the liver parenchyma without an intervening fibrous rim. Cancer cells do not invade the hepatocyte plates, they do not replace the hepatocytes, and they do not co-opt the sinusoidal blood vessels. The surrounding liver is composed of hepatocytes that are arranged parallel to the tumour-liver interface and appear slender because they are atrophic or compressed by the growing metastases (Figures 1G and 1H).

Liver metastases with a sinusoidal HGP are characterized by cancer cells that fill the sinusoidal vascular spaces (Figure 1I). The sinusoidal HGP appears limited to patients with aggressive disease and is more frequently encountered in autopsy specimens, which could imply that it is a feature of end-stage disease ^7, 34–37^. Liver metastases can also spread along the portal tracts. Cancer cells can invade the fibrous stroma of these tracts, fill the lumen of portal vein branches or the lymphatic vessels, or grow along nerves (neurotropism) and blood vessels (angiotropism). In addition, cancer cells can proliferate inside biliary ducts of the portal tracts by replacing the normal epithelial lining of these ducts (Figures 1J and 1K).

Tumour type-dependent differences in the growth patterns have been described. For example, when comparing the replacement HGP in breast cancer metastases and CRLM, the histological characteristics of replacement growth were often present from the tumour-liver interface and up to the centre of the metastases in the breast cancer cases, while they were limited to the interface in all CRLM^38^. Also, the presence of single cancer cells in the liver parenchyma at a distance from the tumour-liver interface in replacement-type liver metastases (so called angiotropic extravascular migration) appears to be more obvious in melanoma liver metastases than in liver metastases of CRC or other carcinomas (unpublished observations).

### Update of the cut-off value to categorize patients with colorectal cancer according to the histopathological growth pattern of the liver metastases

Given that a single liver metastasis can be composed of regions with different growth patterns, this histological parameter is assessed by estimating the relative fraction of the total length of the interface for each growth pattern present in the metastasis. In cases of multiple sections per metastasis or multiple liver metastases per patient, the mean percentage across sections and lesions, respectively, is calculated^1^. In the previous version of the scoring guidelines, a 50% cut-off was proposed to categorize patients, based on its prognostic value. This approach generated four distinct HGP classes: ‘predominant desmoplastic’, ‘predominant replacement’, ‘predominant pushing’ and a ‘mixed’ class in the absence of a predominant HGP. Multiple studies have demonstrated a favourable outcome in patients with CRC liver metastases with a predominant desmoplastic HGP (Table 1).

However, the results of a study by Galjart and colleagues from the Erasmus Medical Centre in Rotterdam^18^ provide a strong rationale for revising the cut-off value used to clinically categorize patients with CRC liver metastases according to the HGP. The study compared different cut-offs based on a large dataset of patients with CRLM. The results suggest that the prognosis of patients with resected CRC liver metastases is primarily determined by the presence of a replacement and/or a pushing growth pattern as opposed to a pure desmoplastic growth pattern (corresponding to 100% of the assessed tumour-liver interface). Favourable survival rates were demonstrated only for patients with liver metastases with complete desmoplastic growth, a condition present in 24% of all patients included in the study by Galjart et al (2019)^18^. Remarkably, non-desmoplastic growth - of any fraction - reduced the 5-year OS rate from 78% to 37% in the cohort of patients who did not receive pre-surgery systemic treatment (adjusted HR 0.39; 95% CI: 0.23-0.67) and from 53% to 40% in the cohort of patients who did receive pre-surgery systemic treatment (adjusted HR 0.92; 95% CI: 0.64- 1.30). This difference in outcome was recently confirmed in a large multicentre external validation study^19^.

We now present a comprehensive clinical evaluation of a large international multicentre cohort of 1931 patients with CRC in which we assessed the impact on outcome using the recent ‘Rotterdam cut-off’^18, 19^ compared to the ‘predominant HGP cut-off’ described in the original international consensus guidelines^1^. The clinicopathological baseline and treatment characteristics are summarized in Table 3. The median follow-up for survivors was 67 months (interquartile range: 34 – 112 months). When applying the Rotterdam cut-off, 1516 (79%) patients had non-desmoplastic liver metastases and 21% had pure desmoplastic liver metastases. Of the 1516 patients with a non-desmoplastic HGP, 201 (10%), 549 (28%), 305 (16%), and 461 (24%) patients had liver metastases with a 100%, 67.1-99%, 33.1-67%, and 0.1- 33% non-desmoplastic HGP, respectively (Table 4). When patients were classified according to the predominant HGP cut-off, 839 (43%) patients had liver metastases with a predominant replacement HGP, 19 (1%) with a predominant pushing HGP, 1031 (53%) with a predominant desmoplastic HGP, and 42 (2%) with a mixed HGP (Table 4). The following findings support the ‘Rotterdam cut-off’:

1. Patients with resected CRC liver metastases that possess an exclusively desmoplastic growth pattern have a clear survival advantage over all other patients. Median OS (months (95% CI)) for desmoplastic versus non-desmoplastic patient cohorts is 88 (77-112) versus 53 (49-58) months, respectively. Median DFS for desmoplastic versus non-desmoplastic patient cohorts is 24 (20-33) versus 11 (11-12) months, respectively (Figures 2A and 2B, Table 4). The adjusted HRs for OS and DFS (95% CI) are 0.64 (0.52- 0.78) and 0.61 (0.52-0.71), respectively (Table 4).
2. There is no difference in survival among patients belonging to the discrete non-desmoplastic classes (Figures 2C and 2D, Table 4). This probably explains why the survival advantage of the favourable patient cohort over the unfavourable patient cohort is less pronounced when the predominant HGP cut-off algorithm is used (Figures 2E and 2F, Table 4). For example, the adjusted HR for OS is 0.64 (95% CI: 0.52- 0.78) versus 0.76 (95% CI: 0.65-0.88) respectively, when comparing the Rotterdam and the ‘predominant HGP’ cut-offs (Table 4). A similar difference of 0.61 (95% CI: 0.52- 0.71) versus 0.82 (95% CI: 0.73-0.93) can be observed for DFS (Table 4).
3. The learnability and accuracy of HGP-scoring according to the new cut-off have been shown to be high^39^. Moreover, this algorithm represents a simplified method of HGP scoring when considering prognostic impact. Indeed, when a non-desmoplastic component (replacement or pushing) is detected while analysing a series of H&E-stained sections from a patient, the result is clear, and no further scoring is required. However, for scientific research purposes, and to further validate the new cut-off approach, care should be taken not to compromise the acquisition of more detailed quantitative data and assessing the HGPs in all the available H&E-stained sections of all the resected liver metastases is still preferred.

**Figure 2.**
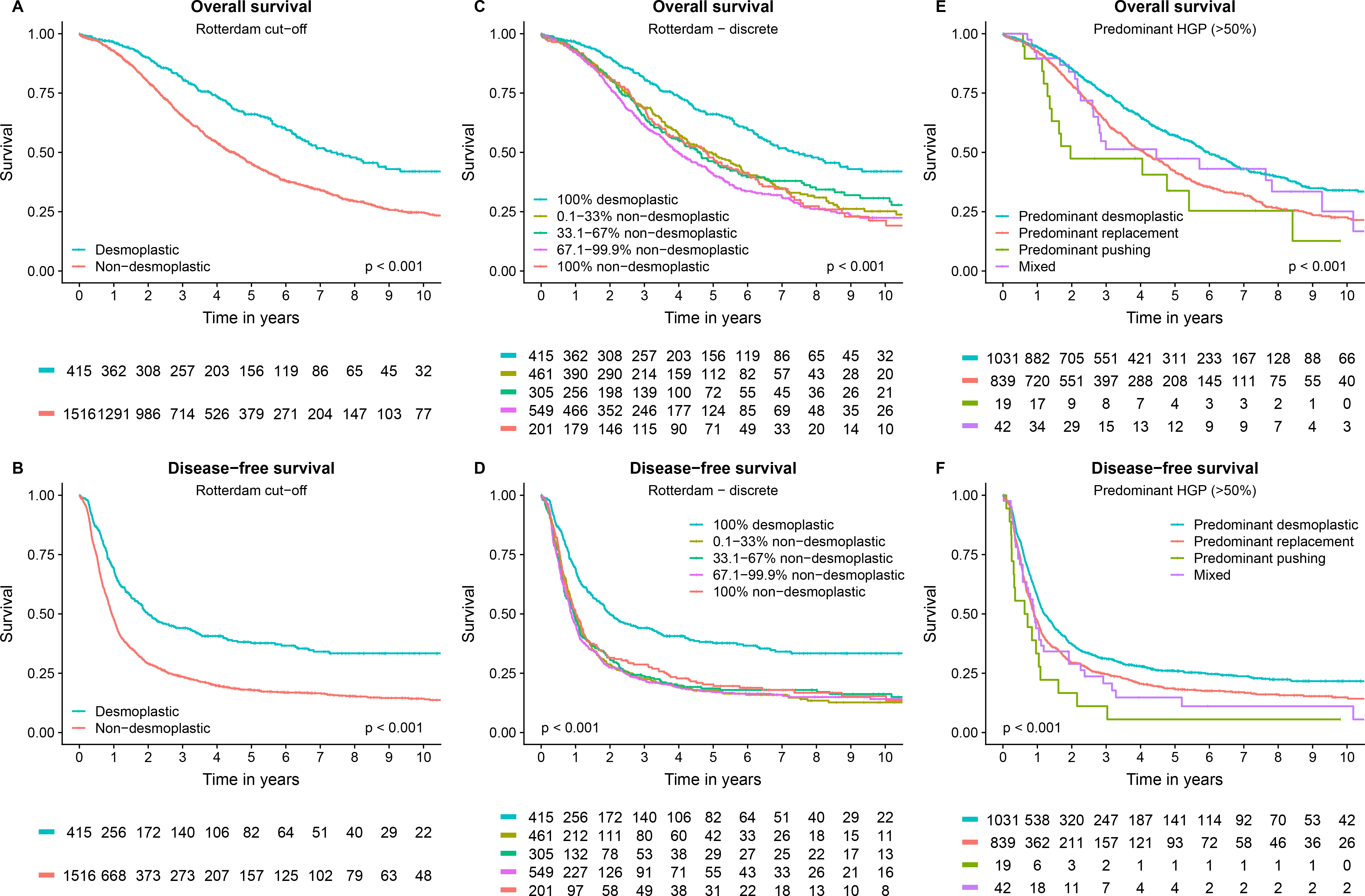
Kaplan–Meier curves depicting overall and disease-free survival of patients with colorectal liver metastases, stratified by the new cut-off for histopathological growth patterns categorisation (A to D) and by the predominant growth pattern (E and F). N = 1931 patients with resected colorectal liver metastases.

**Table 3.** Clinicopathological baseline and treatment characteristics of the cohort of 1931 patients used to evaluate the impact on outcome of the new cut-off for patient categorisation according to the histopathological growth pattern of resected colorectal liver metastases. Abbreviations: ASA = American Society of Anaesthesiologists; CEA = carcinoembryonic antigen; CRLM = colorectal liver metastasis; Erasmus MC = Erasmus Medical Center Cancer Institute; IQR = interquartile range; MSKCC = Memorial Sloan Kettering Cancer Center; Radboud UMC = Radboud University Medical Center.

|  |  | missing (%) | n = 1931 (%) |
| --- | --- | --- | --- |
| Cohort | *Erasmus MC* |  | 903 (47) |
|  | *MSKCC* |  | 716 (37) |
|  | *Radboud UMC* |  | 312 (16) |
| Age at resection CRLM - (*median [IQR]*) |  |  | 64.0 [56.0, 71.0] |
| Gender | *Male* |  | 1170 (61) |
|  | *Female* |  | 761 (39) |
| ASA classification | *ASA I-II* | 39 (2) | 1284 (68) |
|  | *ASA >II* |  | 609 (32) |
| Primary tumour location | *Left-sided* | 62 (3) | 458 (25) |
|  | *Right-sided* |  | 798 (43) |
|  | *Rectal* |  | 613 (33) |
| T-stage | *pT 0-2* | 87 (5) | 287 (16) |
|  | *pT 3-4* |  | 1557 (84) |
| N-stage | *N0* | 31 (2) | 729 (38) |
|  | *N+* |  | 1172 (62) |
| Number of CRLM - (*median [IQR]*) |  | 12 (1) | 2.0 [1.0, 3.0] |
| Diameter of largest CRLM in cm - (*median [IQR]*) |  | 35 (2) | 2.8 [1.9, 4.5] |
| Disease-free interval in months* - (*median [IQR]*) |  | 14 (1) | 1.0 [0.0, 17.0] |
| Synchronous (DFI ≤3 months) | *Synchronous* |  | 1023 (53) |
|  | *Metachronous* |  | 908 (47) |
| Preoperative CEA in µg/L - (*median [IQR]*) |  | 143 (7) | 11.0 [4.0, 33.7] |
| Perioperative systemic chemotherapy | *No chemotherapy* | 41 (2) | 773 (41) |
|  | *Neoadjuvant only* |  | 689 (36) |
|  | *Adjuvant only* |  | 232 (12) |
|  | *Perioperative* |  | 196 (10) |
| Resection margin involved | *No* | 10 (1) | 1675 (87) |
|  | *Yes* |  | 247 (13) |
| Extrahepatic disease** | *No* |  | 1731 (90) |
|  | *Yes* |  | 200 (10) |

\* *Between resection of primary tumour and detection of CRLM*
\*\* *Defined as any extrahepatic disease with the exception of the primary tumour present at the time of or prior to first CRLM surgery.*
Abbreviations in alphabetical order: ASA: American Society of Anaesthesiologists; CEA: carcinoembryonic antigen; CRLM: colorectal liver metastasis; DFI: disease-free interval; Erasmus MC: Erasmus MC Cancer Institute; IQR: interquartile range; MSKCC: Memorial Sloan Kettering Cancer Center; Radboud UMC: Radboud University Medical Center.

**Table 4.** Overall and disease-free survival estimates for different histopathological growth pattern cut-offs in 1931 patients treated with curative intent resection for colorectal liver metastases. Abbreviations: ASA = American Society of Anaesthesiologists; CEA = carcinoembryonic antigen; CRLM = colorectal liver metastasis; HGP = histopathological growth pattern; NA = not available.

| Cut-off | n (%) | Overall survival (95%CI) |  |  |  | Disease-free survival (95%CI) |  |  |  |
| --- | --- | --- | --- | --- | --- | --- | --- | --- | --- |
|  |  | 5 year - % | 10 year - % | Median - months | Adjusted HR* | 5 year - % | 10 year - % | Median - months | Adjusted HR* |
| Entire cohort | 1931 (100) | 50 (47-53) | 28 (26-32) | 60 (56-65) | - | 22 (20-24) | 18 (16-21) | 13 (12-13) | - |
| Rotterdam cut-off |  |  |  |  |  |  |  |  |  |
| Non-desmoplastic | 1516 (79) | 45 (42-48) | 25 (22-28) | 53 (49-58) | *reference* | 18 (16-20) | 14 (12-17) | 11 (11-12) | *reference* |
| Desmoplastic | 415 (21) | 66 (61-72) | 42 (35-50) | 88 (77-112) | 0.64 (0.52-0.78) | 38 (33-43) | 33 (28-39) | 24 (20-33) | 0.61 (0.52-0.71) |
| Rotterdam cut-off - discrete |  |  |  |  |  |  |  |  |  |
| 100% non-desmoplastic | 201 (10) | 48 (41-56) | 21 (14-31) | 57 (44-69) | *reference* | 20 (15-27) | 15 (10-23) | 12 (10-15) | *reference* |
| 67.1-99.9% non-desmoplastic | 549 (28) | 41 (36-46) | 22 (18-28) | 48 (44-55) | 1.13 (0.89-1.45) | 17 (14-21) | 14 (11-18) | 11 (10-12) | 1.10 (0.89-1.35) |
| 33.1-67% non-desmoplastic | 305 (16) | 46 (40-54) | 31 (24-39) | 55 (45-66) | 0.95 (0.72-1.25) | 19 (14-24) | 16 (12-22) | 11 (10-13) | 0.99 (0.79-1.25) |
| 0.1-33% non-desmoplastic | 461 (24) | 49 (44-55) | 25 (20-32) | 58 (51-70) | 1.05 (0.81-1.36) | 18 (14-22) | 13 (9-17) | 12 (11-13) | 1.05 (0.85-1.31) |
| 100% desmoplastic | 415 (21) | 66 (61-72) | 42 (35-50) | 88 (77-112) | 0.67 (0.51-0.88) | 38 (33-43) | 33 (28-39) | 24 (20-33) | 0.64 (0.51-0.80) |
| Predominant HGP cut-off (>50%) |  |  |  |  |  |  |  |  |  |
| Predominant replacement | 839 (43) | 42 (38-46) | 23 (19-27) | 49 (45-56) | *reference* | 18 (16-22) | 15 (12-18) | 11 (10-12) | *reference* |
| Predominant pushing | 19 (1) | 34 (17-66) | NA | 24 (17-NA) | 1.03 (0.55-1.95) | 6 (1-37) | NA | 8 (4-19) | 1.12 (0.64-1.95) |
| Mixed | 42 (2) | 47 (33-69) | 25 (11-55) | 53 (33-129) | 1.04 (0.65-1.68) | 15 (7-32) | 11 (4-29) | 11 (8-23) | 1.23 (0.84-1.78) |
| Predominant desmoplastic | 1031 (53) | 57 (53-61) | 34 (30-39) | 72 (67-79) | 0.76 (0.65-0.88) | 26 (23-29) | 22 (19-25) | 14 (13-17) | 0.82 (0.73-0.93) |

\* Multivariable regression model (n=1565 included in full-case analyses) corrected for age, gender, ASA class, primary tumour location, T-stage, nodal status, disease-free interval (continuous), number of CRLM (continuous), size of largest CRLM (continuous), preoperative CEA (continuous), extrahepatic disease, resection margin status, and perioperative systemic chemotherapy.
Abbreviations in alphabetical order: ASA: American Society of Anaesthesiologists; CEA: carcinoembryonic antigen; CRLM: colorectal liver metastasis; HGP: histopathological growth pattern; HR: hazard ratio; NA: not available.

The international group of authors of this second consensus guidelines for scoring HGPs of hepatic metastases therefore proposes to test this algorithm in prognostic studies with other primary tumour types as well. In studies that aim at deciphering the molecular underpinnings of the different growth patterns, a cut-off agnostic approach should probably be adopted, to not obscure lessons to be learned from inter-tumour heterogeneity of the HGPs.

### Categorization of the histopathological growth patterns of non-CRC liver metastases

Distinct HGPs have been identified in liver metastases from a broad range of primary solid tumours, mostly carcinomas. The replacement (also referred to sometimes as ‘replacing’, ‘trabecular’ or ‘infiltrative’) growth pattern, the desmoplastic growth pattern (also sometimes called ‘encapsulated’) and the pushing growth pattern (also sometimes called ‘expansive’) have been described in liver metastases from primary lung, pancreatic, stomach, gallbladder/bile duct and breast carcinoma^5, 38, 40–42^. The study of HGPs in liver metastases from these tumour types is relevant given that, for example, about 11% of patients with lung carcinomas, 36% of patients with pancreatic carcinoma and 14% of patients with stomach cancer have liver metastases at diagnosis^43^. The sinusoidal growth pattern has been encountered in autopsy specimens of patients with non-small cell lung cancer (NSCLC) and breast cancer^7, 34–37^. In addition to carcinomas, the desmoplastic, pushing, replacement and sinusoidal growth patterns have also been identified in hepatic metastases of both skin and uveal melanoma^44–46^. Additional types of HGP have also been described in uveal melanoma, however without evaluation of the interface between liver metastases and the surrounding liver tissue^46, 47^. In these studies, the different results reported may be ascribed to the sources of material studied, almost entirely derived from autopsies, and of partial biopsy samplings. The HGPs have recently also been identified in sarcoma-derived hepatic metastases, in a study describing the HGPs in a cohort of patients with non-colorectal, non-neuroendocrine liver metastases^48^.

Although the prognostic/predictive role of the HGPs has been studied mainly in patients with CRC^1, 18, 19, 49^, there are recent reports on the impact of the HGPs on outcome in patients with liver metastatic melanoma, breast carcinoma and pancreatic cancer^5, 42, 44, 45^. In a study of 42 patients with skin melanoma, the presence of any replacement HGP (1% of the tumour-liver interface or more), present in 20 patients (48%), significantly predicted worse overall survival while the 100% desmoplastic HGP correlated with improved OS, an effect that continued to be significant upon multivariate analysis (HR = 3.79, p = 0.01)^45^. In a study of 41 patients with liver metastatic uveal melanoma, the dominant HGP (>50% of tumour-liver interface) was used to categorize patients^44^. A dominant replacement HGP, present in 30 patients (73%), predicted diminished OS with a HR in multivariate analysis of 6.51 (p = 0.008). An updated analysis with extension of the patient cohort and categorisation according to the 100% desmoplastic HGP cut-off has recently been completed (Barnhill et al, manuscript in preparation).

The HGPs of breast cancer liver metastases have only been sporadically studied and have been mainly described in autopsy specimens^34, 35, 38, 41^. In this context, and when compared with CRC liver metastases, the replacement HGP and even the sinusoidal HGP are more frequently encountered in breast cancer liver metastases. Surgical removal of breast cancer hepatic metastases is still rarely practiced. However, there is a subpopulation of patients with liver metastatic breast carcinoma for whom a favourable course after resection has been documented, contradicting the common idea that breast cancer is always a systemic disease^50^ and a rationale behind ongoing clinical trials, for example BreCLIM-2 (ClinicalTrials.gov Identifier: NCT04079049). With this in mind, Bohlok et al (2020)^42^ have scored the HGPs in 36 patients who underwent surgical resection for breast cancer liver metastases. Given that only one patient presented with liver metastases with a pure desmoplastic HGP while 16 patients had liver metastases with a pure replacement HGP, a pragmatic approach was adopted to categorize patients as having liver metastases with ‘100% replacement’ versus ‘any desmoplastic’ HGP. The study confirmed the association of replacement HGP liver metastases with poor outcome as observed with other tumour types. Indeed, all patients with a pure replacement HGP relapsed within 2 years after surgery. In addition, even in this small cohort of patients, improved OS was observed for patients with ‘any desmoplastic’ HGP liver metastases as compared to the other patients upon multivariate analysis (HR = 0.20, p = 0.023)^42^. A large international study has recently been undertaken by several authors of the guidelines to further address the impact of the HGPs on outcome in patients with liver metastatic breast cancer.

More than one-third of patients with neuroendocrine tumours (NETs) present with distant disease, with the liver being the most common metastatic site. Although newer therapeutic options are becoming available, resection of NET liver metastases is still often performed^51^. Given the broad spectrum of NETs, from well-differentiated NETs to poorly differentiated neuroendocrine carcinomas, it would be interesting to study the HGPs of NET liver metastases. To the best of our knowledge, this has not been done yet.

In conclusion, the distinct HGPs can be identified independently of the primary solid tumour type and the desmoplastic HGP is invariably associated with better outcome than the replacement HGP, after surgical removal of liver metastases. This is consistent with the idea that common, tumour type-independent and liver-specific biological programs are activated in liver-metastatic cancer cells and shape growth pattern emergence in the liver^52^.

### Clinical significance of the pushing growth pattern

The prognostic/predictive value of the pushing HGP is still unclear. Before the first international guidelines were published, there were no unequivocal instructions for distinguishing the pushing HGP from the replacement HGP where tumour cells appear to push away the liver parenchyma (so called pushing-type or type-2 replacement HGP)^1^. As a result, the proportion of metastases with a pushing HGP has been overestimated in studies carried out prior to the publication of the first consensus guidelines^49^. For example, Nielsen et al. (2014)^53^ and Eefsen et al. (2015)^54^ reported that 45% of the patients with resected CRC liver metastases presented with a dominant pushing HGP. By applying the consensus guidelines of 2017, the proportion of metastases with a pushing HGP was found to be reproducibly smaller across more recent studies. In the study by Galjart et al. of 2019^18^, for example, less than 1% of patients presented with a dominant pushing HGP in their CRC liver metastases. Determining the clinical value of the pushing HGP will therefore only be possible in large multi-centre studies.

### The histopathological growth patterns and treatment response

Several observations suggest that systemic treatment can alter the HGP of liver metastases. In the study by Frentzas et al. (2016)^41^, the growth pattern of recurrent CRLM, defined as those metastases that were not detectable by imaging before systemic treatment but appeared during bevacizumab-chemotherapy, was compared with the growth pattern of metastases that were already visible before systemic treatment. The recurrent metastases more often demonstrated a replacement HGP when compared to the metastases that were already visible before systemic treatment (80% versus 50%). In support of these observations, several preclinical studies have demonstrated the switch from an angiogenic to a vessel co-opting growth pattern associated with resistance to treatment with anti-VEGF drugs in several malignancies. These include hepatocellular carcinoma^55^, lung metastases of renal cell carcinoma^56^, brain metastases of melanoma^57^ and glioblastoma^58^.

Other studies^59, 60^ found associations between systemic treatment of patients with CRLM and histological characteristics that are highly suggestive of replacement growth. The so-called ‘dangerous halo’ consists of an irregular tumour-liver interface in a CRLM that was seen selectively in patients that received chemotherapy before partial hepatectomy. Although beyond the scope of the Mentha et al. study, the histological images in their report show that the ‘dangerous halo’ consists of areas of replacement growth while the lesion without the ‘dangerous halo’ has a desmoplastic HGP (Figure 1 in Mentha et al. (2009)^59^). Taken together, the findings of Frentzas et al. (2016)^41^ and the reports on the ‘dangerous halo’^59, 60^ link the replacement HGP to chemotherapy resistance with or without anti-VEGF treatment in patients with liver metastatic colorectal cancer.

There are, however, studies suggesting that chemotherapy induces the desmoplastic growth pattern in patients with replacement-type CRLM^18, 61^. Nierop and colleagues (2021)^61^ have assessed the HGP of resected liver metastases in three cohorts of respectively 877, 1203 and 70 patients with CRC, respectively. The latter cohort was derived from a phase III clinical trial in which patients were randomized between either peri-operative chemotherapy and resection or resection only. In all three cohorts, the average presence of the desmoplastic HGP at the tumour-liver interface was significantly higher in patients with pre-operative chemotherapy compared to chemo-naïve patients (67% versus 43%, 63% versus 40%, and 61% versus 33%, respectively (p<0.005)). The fact that this shift in HGP was observed in a randomised study is consistent with a lack of selection in the association of pre-operative chemotherapy and the desmoplastic HGP. However, it remains to be determined whether chemotherapy induces a transformation of replacement-type liver metastases into lesions that form a desmoplastic rim or whether pre-existing desmoplastic lesions are more resistant to chemotherapy.

Taken together, it appears that a transition from one HGP to another could occur in patients with CRLM following systemic treatment. However, despite all the studies discussed above, a reliable assessment in individual patients of the effect of systemic treatment on the HGPs of liver metastases will only be possible when non-invasive imaging (as discussed below) or blood analyses will be available to identify the HGPs at several time points during treatment. One promising blood marker was recently proposed^16^. Circulating extracellular vesicles (EVs) derived from patients with replacement-type CRLM exhibited significantly higher protein expression of Claudin-2 relative to EVs isolated from patients with desmoplastic liver metastases. Thus, high protein levels of Claudin-2 in EVs isolated in the blood circulation of patients with liver metastatic CRC may predict the replacement HGP in CRLM.

### Standard method for assessment of the histopathological growth patterns of liver metastases

The updated consensus guidelines for tissue sampling of surgical liver resections and for scoring and reporting of the HGPs of liver metastases are presented in Table 5. The proposed sampling guidelines are not based on published experimental evidence but are rather an empirical approach^62^. Given that the invasion front of liver metastases is often heterogeneous in respect to HGPs, a balance must be struck between accurate assessment of growth patterns and practical feasibility of sampling in a pathology laboratory. In addition, the sampling procedure may be tumour-type dependent. For example, when dealing with CRLM, a two-step approach can be envisaged for clinical routine, given that the presence of any proportion of the interface with a non-desmoplastic HGP in any of the resected metastases has clear prognostic significance^18, 19^. Initial sampling or scoring may consist of a limited number of paraffin blocks and in the event that a region with a non-desmoplastic growth is identified in the H&E-stained sections, the patient will be categorized into the corresponding HGP group. In accordance with our proposed updated guidelines, additional and more extensive sampling or scoring will only be necessary if no regions with non-desmoplastic growth are encountered at initial sampling or scoring.

**Table 5.**
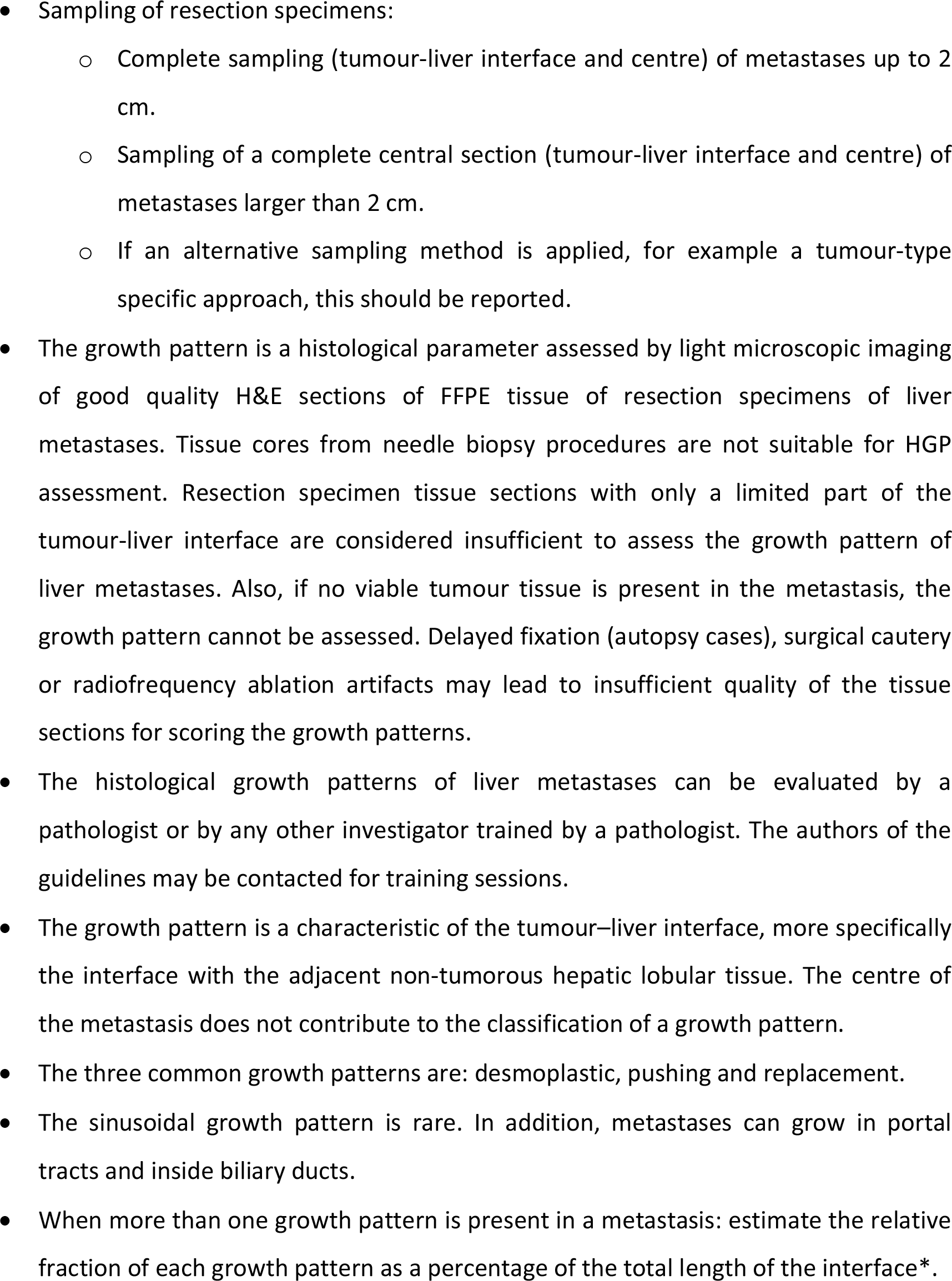
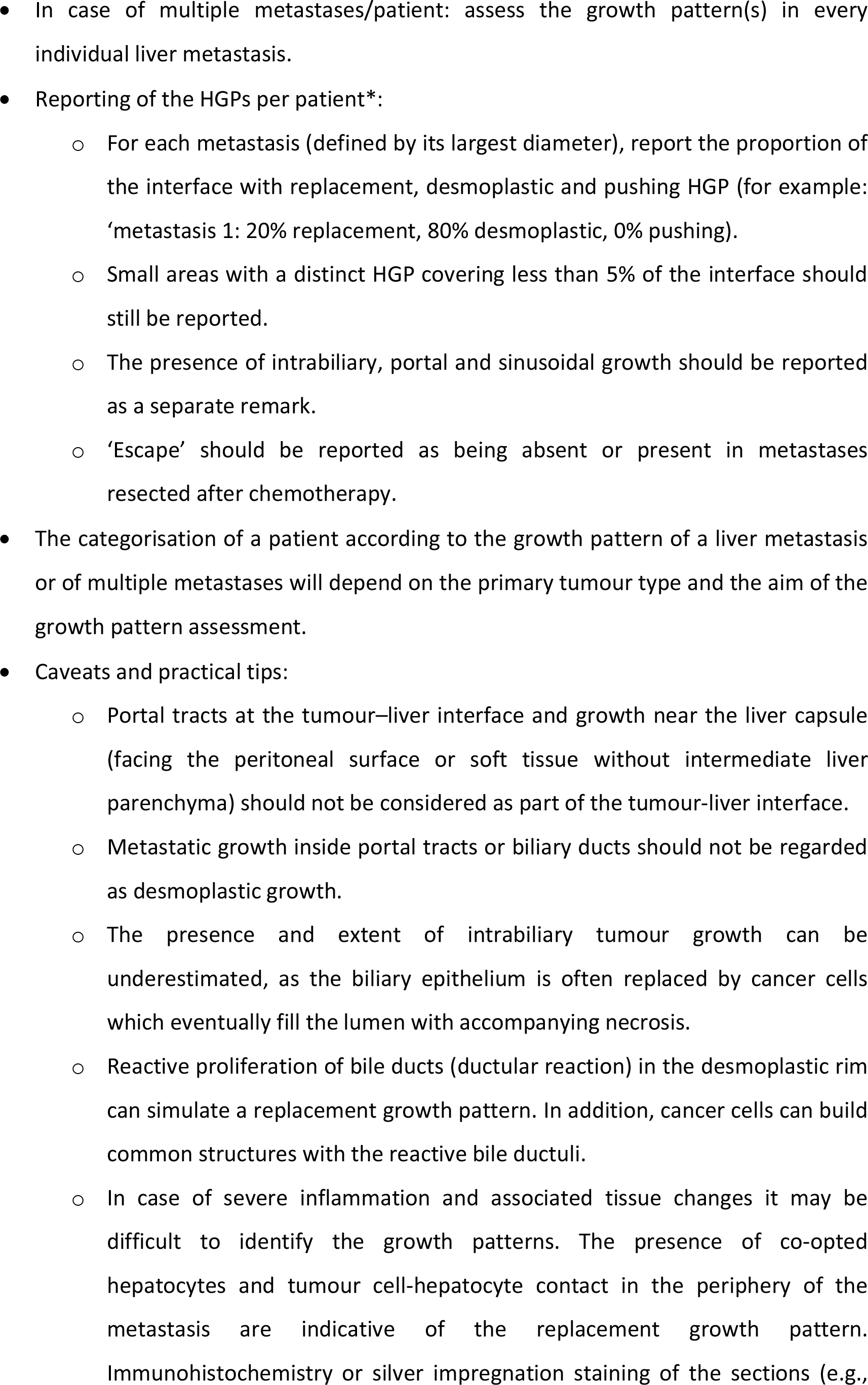
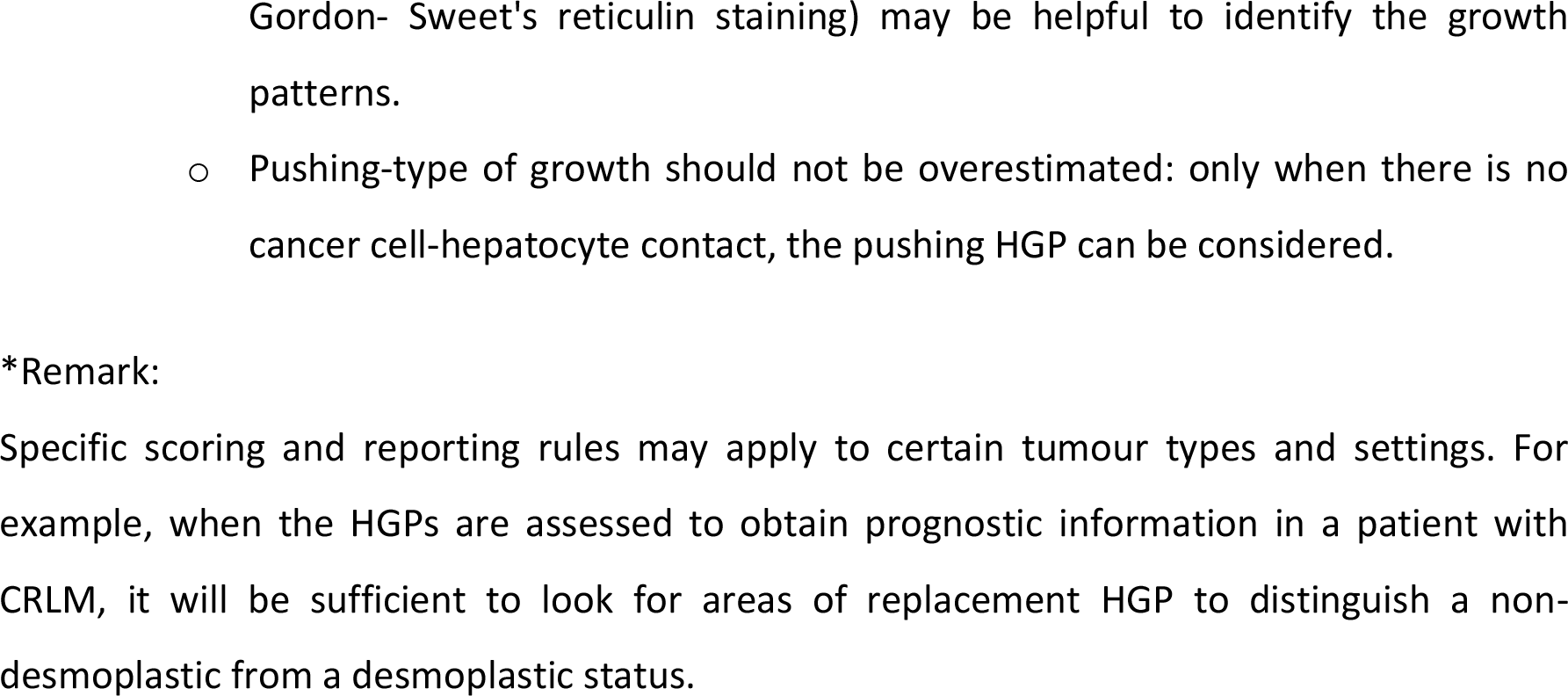
Updated standard method for histopathological growth pattern assessment of liver metastases.

In reporting the HGPs of liver metastases, several factors will need to be considered. The context of HGP assessment and the primary tumour type need to be considered because they will determine how a patient will be categorised based on the liver metastasis HGP. For example, for patients with CRLM, the HGP can provide prognostic information. For these patients, categorization can, therefore, be based on the cut-off specified in the current guidelines. For other primary tumour types, large studies that have defined a clinically relevant cut-off value are still lacking and data reporting should be as precise as possible, in order for the HGP-score to be available for future data analyses because predictive and prognostic HGP cut-off values may be different for different primary tumour types.

There are essentially two ways to report HGPs when multiple metastases are resected. One approach simply averages the scores for each HGP (desmoplastic, replacement, pushing) across every available H&E-stained section for all the resected metastases. The other approach uses an average of the scores for each HGP of all the available H&E-stained sections for each individual metastasis separately and reports a score for every metastasis that has been resected. The latter approach may be used when biological differences between metastases are expected, for example related to a difference in response to pre-surgery systemic treatment.

With the aim of identifying the presumed treatment-induced transition towards the replacement HGP in future studies, we propose the following clinicopathological definition of an ‘escape’ phenotype: ‘Liver metastases resected after pre-operative systemic treatment combining signs of pathological response in the centre of the metastases while also exhibiting at least a partly preserved desmoplastic rim and small peripheral areas of replacement-type outgrowth or a complete halo of replacement growth’. Typically, these areas of replacement growth do not show any of the characteristic signs of treatment response, as shown in examples in Figure 3. Further information on the clinical value of this phenotype and its biological underpinning will be derived from future studies on the HGPs of liver metastases. We therefore propose to score the presence or absence of ‘escape’ in liver metastases that are resected after administration of systemic pre-operative treatment.

**Figure 3.**
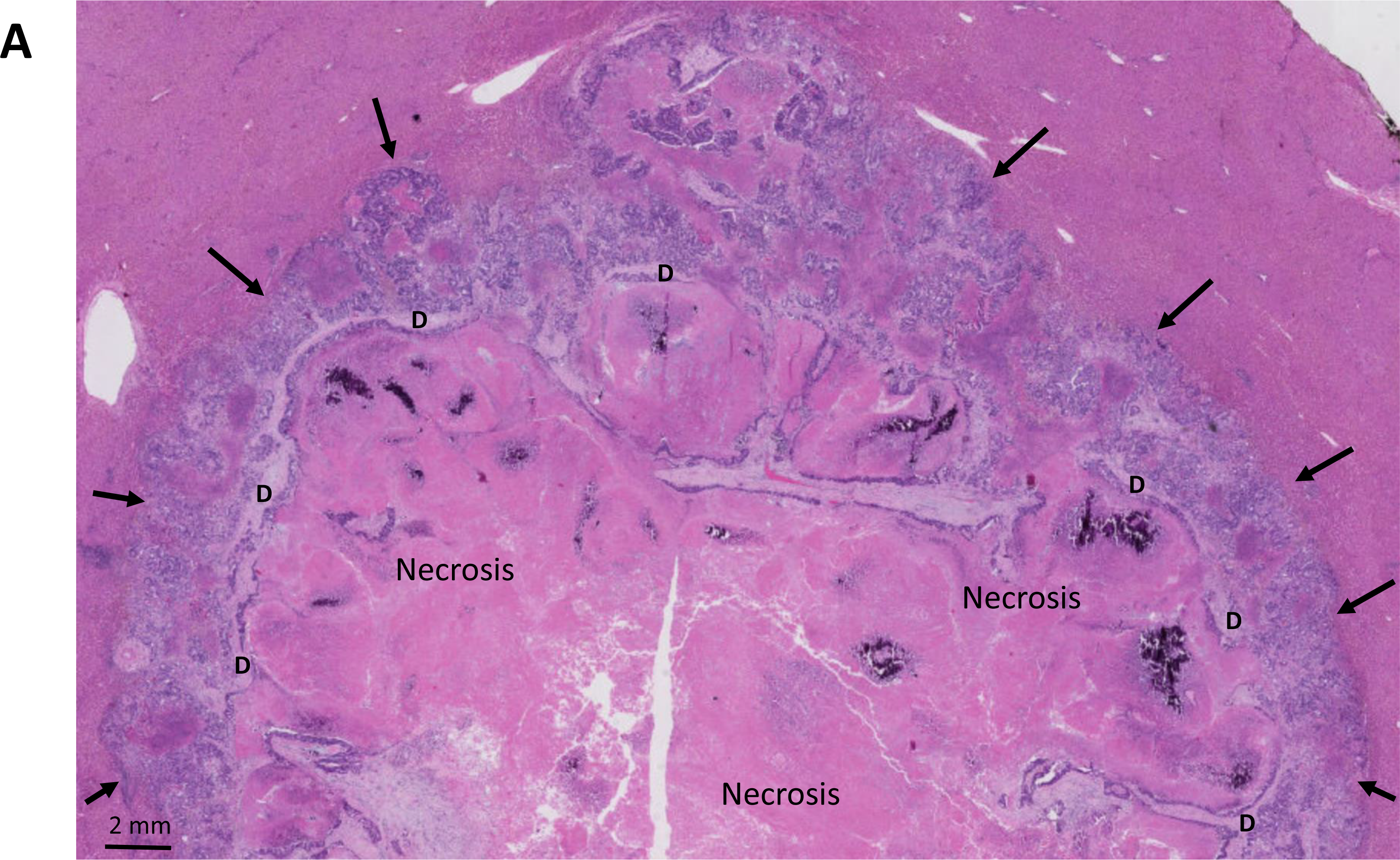
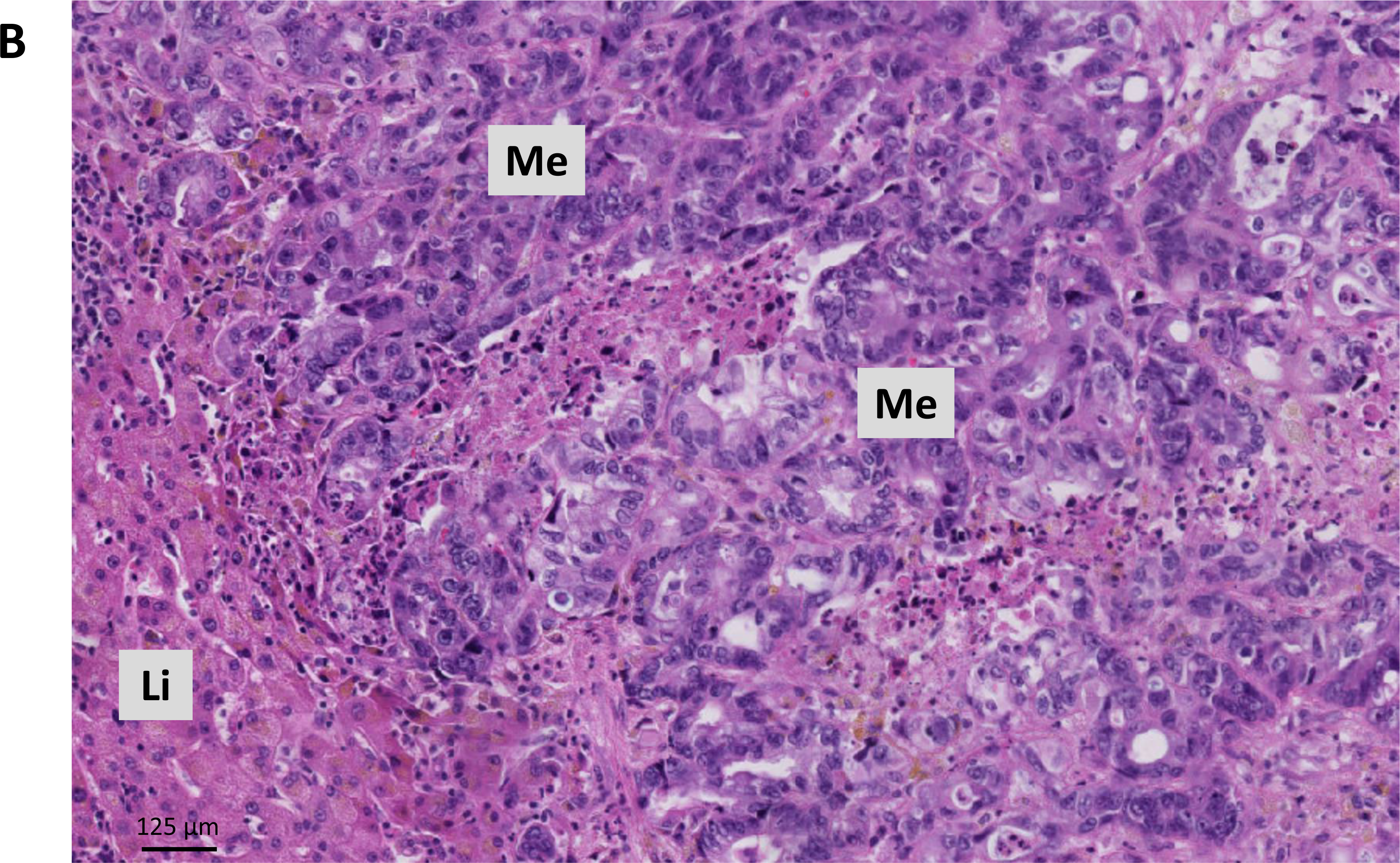
H&E image of the escape phenotype. (A) Low magnification image with large necrotic areas in the centre of the CRC liver metastasis, remnants of the desmoplastic rim (d) and vital replacement-type outgrowth at the tumour-liver interface (arrows). (B) Higher magnification of the ‘escape’ area with replacement HGP. Li, liver; Me, metastatic tumour tissue.

### Immunohistochemical staining as an aid to scoring HGPs

In some liver metastases, the histology is more complex, and this can result in a less straightforward assessment of the HGPs. The ‘caveats’ are listed in the Table 5. Although the assessment of HGPs of liver metastases is based exclusively on H&E-stained tissue sections, additional immunohistochemical analyses may provide clarity when these challenging conditions arise.

One example is the presence of an extensive immune cell infiltrate that obscures the tumour- liver interface. In this case, the presence or absence of contact between tumour cells and hepatocytes and the degree of hepatocyte co-option will determine whether the replacement HGP must be considered. A double immunostaining approach coupling a hepatocyte marker and a tumour cell marker can also be useful in such cases. For example, for liver metastases from a colorectal carcinoma, the combination of antibodies directed against caudal type homeobox 2 (CDX-2), cytokeratin (CK) 20 or CK19 (tumour cells) and Hepar-1, arginase1, or CK18 (hepatocytes) can be used (Figure 4A). This immunostaining may also help to distinguish a replacement HGP in which the liver cell plates are pushed away from the rare pushing HGP (Figure 4B).

**Figure 4.**
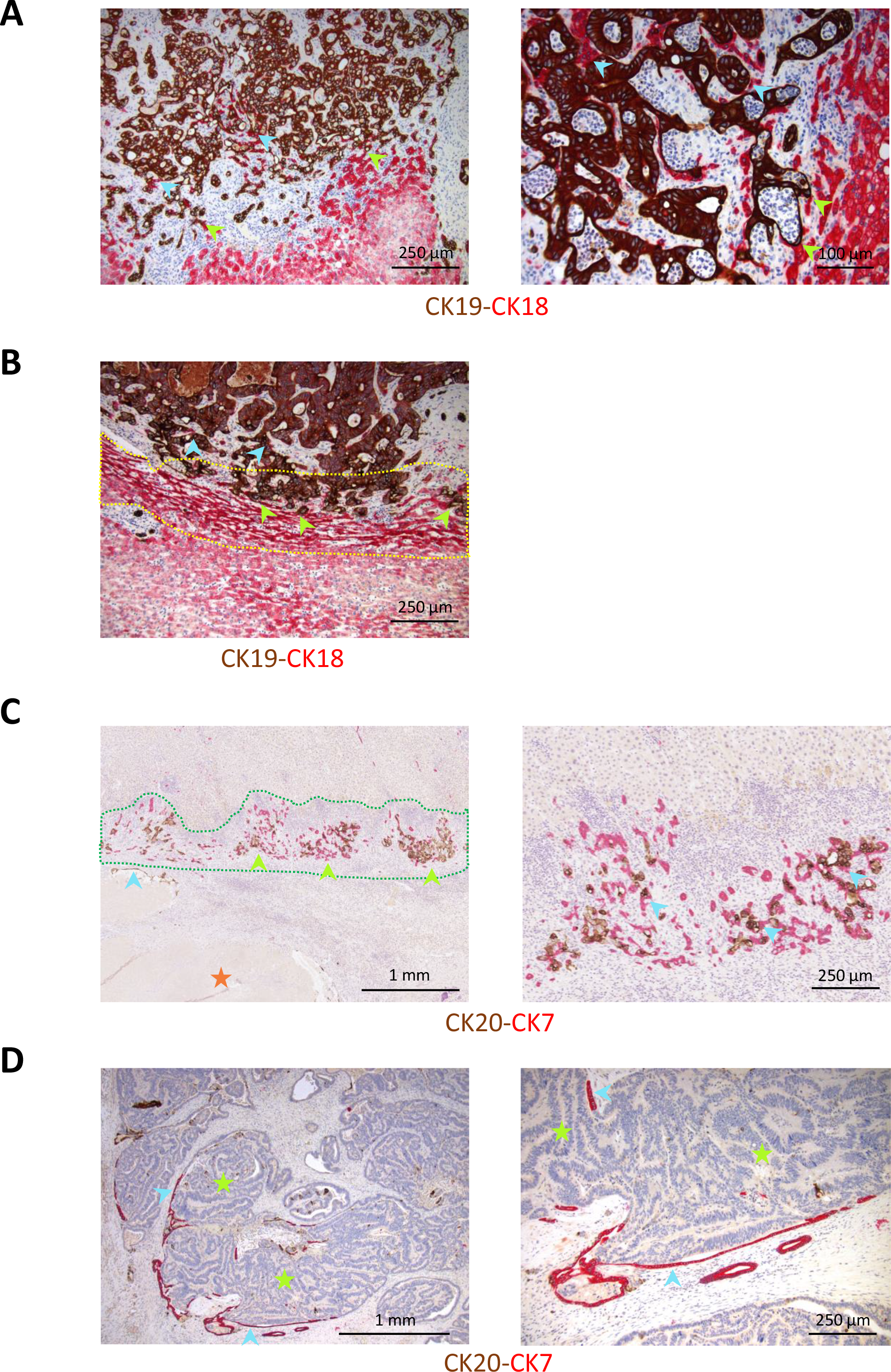
Immunohistochemical staining as an aid to HGP scoring A. Detection of the replacement HGP in the presence of an extensive immune cell infiltrate that obscures the tumour-liver interface by identification of cancer cell-hepatocyte contact (green arrowheads) at the tumour-liver interface and co-option of hepatocytes (blue arrowheads) in liver lobules undergoing replacement by cancer cells. CK19 (DAB, brown) stains colorectal cancer cells. CK18 (AP, red) stains hepatocytes. Left: low magnification; Right: high magnification. B. Detection of the pushing-type replacement (type 2) HGP in which the hepatocyte plates are slender (yellow dotted area) and arranged in parallel with the tumour-liver interface. Green arrowheads indicate cancer cell-hepatocyte contact and blue arrowheads hepatocyte co- option. CK19 (DAB, brown) stains colorectal cancer cells. CK18 (AP, red) stains hepatocytes. C. Prominent ductular reaction at the tumour-liver interface in the desmoplastic HGP. Areas of ductular reaction (green arrowheads) are present in the outer region of the fibrous rim (green dotted region). Cancer cells are (blue arrow) identified in the metastasis centre, adjacent to necrotic areas (orange star). Right. Detail of the ductular reaction at the tumour- liver interface. Cholangiocytes (CK7+) form irregular, angulated, anastomosing ductuli. Note the presence of interspersed cancer cells (CK20+, blue arrows) within the ductuli, forming common ductular structures. CK20 (DAB, brown) stains colorectal cancer cells. CK7 (AP, red) stains cholangiocytes. D. Detection of intrabiliary tumour growth. A discontinuous lining of biliary epithelial cells (blue arrows) can be identified surrounding colorectal cancer cells (sparsely positive for CK20 in this case) with focal contact between colorectal cancer cells and biliary epithelial cells (green stars). CK20 (DAB, brown) stains colorectal cancer cells. CK7 (AP, red) stains cholangiocytes. Left: low magnification; right: high magnification.

A second example where a clear-cut assessment of the HGP may be challenging is the presence of a prominent ductular reaction at the tumour-liver interface. It can indeed be difficult to distinguish cancer cells from cholangiocytes in this ductular reaction, especially when nuclear pleomorphism of the cancer cells is limited and small aggregates or glandular structures of cancer cells are formed. In addition, cancer cells and cholangiocytes can be involved in common ductular structures. A possible solution is to combine cholangiocyte (CK7, CK19 or carbohydrate antigen 19-9 (CA19-9)) and cancer cell markers (for CRLM, for example CK20 or CDX-2) (Figure 4C) as an added tool for the analysis. Double immunostaining for cancer cell and cholangiocyte markers can also be used to identify intrabiliary growth when only a few cholangiocytes remain that are difficult to detect on an H&E-stained section (Figure 4D).

## Perspectives

### Patient-derived xenograft models to study the HGPs of liver metastases

The characterization of the distinct growth patterns using protein-based and genomic approaches on surgically resected clinical specimens has begun to shed light on the underlying biological processes that might drive the formation and growth of these lesions (Table 1). However, the field currently lacks animal models that faithfully recapitulate the specific histological features of these metastases (in particular desmoplastic metastases), necessary for functional dissection of the molecular mediators that are currently only associated with one type of lesion or the other.

To better understand the underlying biology of desmoplastic and replacement liver metastases and to test therapeutic strategies tailored to these distinct lesions, it will be important to develop PDXs that faithfully recapitulate the histological features seen in patients. To this end, members of the Liver Metastasis Research Network at the Goodman Cancer Institute (McGill University) and the McGill University Health Centre have developed a patient-derived xenograft (PDX) pipeline where freshly resected CRLM, or biopsy samples, from the operating theatre are brought immediately to the laboratory and are directly implanted into the livers of SCID/beige mice^16^. The surgical specimen is divided into approximately 1mm^3^ fragments, which are then carefully inserted into an incision made in the left cardiac liver lobe of recipient mice. This approach has led to the successful establishment of more than 30 PDX models that represent both replacement and desmoplastic lesions. Importantly, a high degree of concordance (over 95%) between the HGPs of the metastases that develop in the PDX models, when compared to the metastatic lesion in the patients from which they were derived, has been achieved. In addition, organoids from these PDX models have been generated (PDXOs) and propagated in culture (Tabariès S, Gregorieff A and Siegel P, unpublished observations). When re-injected into the livers of mice, these PDXOs generate desmoplastic or replacement lesions that recapitulate the HGP of the patient sample and PDX model (Figure 5). While these models may provide useful information on the drivers underlying specific HGPs, the lack of an adaptive immune response in the recipient mice, may present a challenge to obtaining complete information on the associated immune microenvironments. Although several methods have been described to generate so-called ‘humanised mice’, a less challenging approach is represented by the patient-derived explants (PDE), ex vivo systems in which the in vivo tissue architecture and immune microenvironment of human tumours can be maintained^63^. These PDE platforms have been shown to be able to predict clinical response to inhibitors of the PD-1-PD-L1 axis in patients with various types of cancer^64^ and might thus be used to study the biology of liver metastases with distinct HGPs.

**Figure 5.**
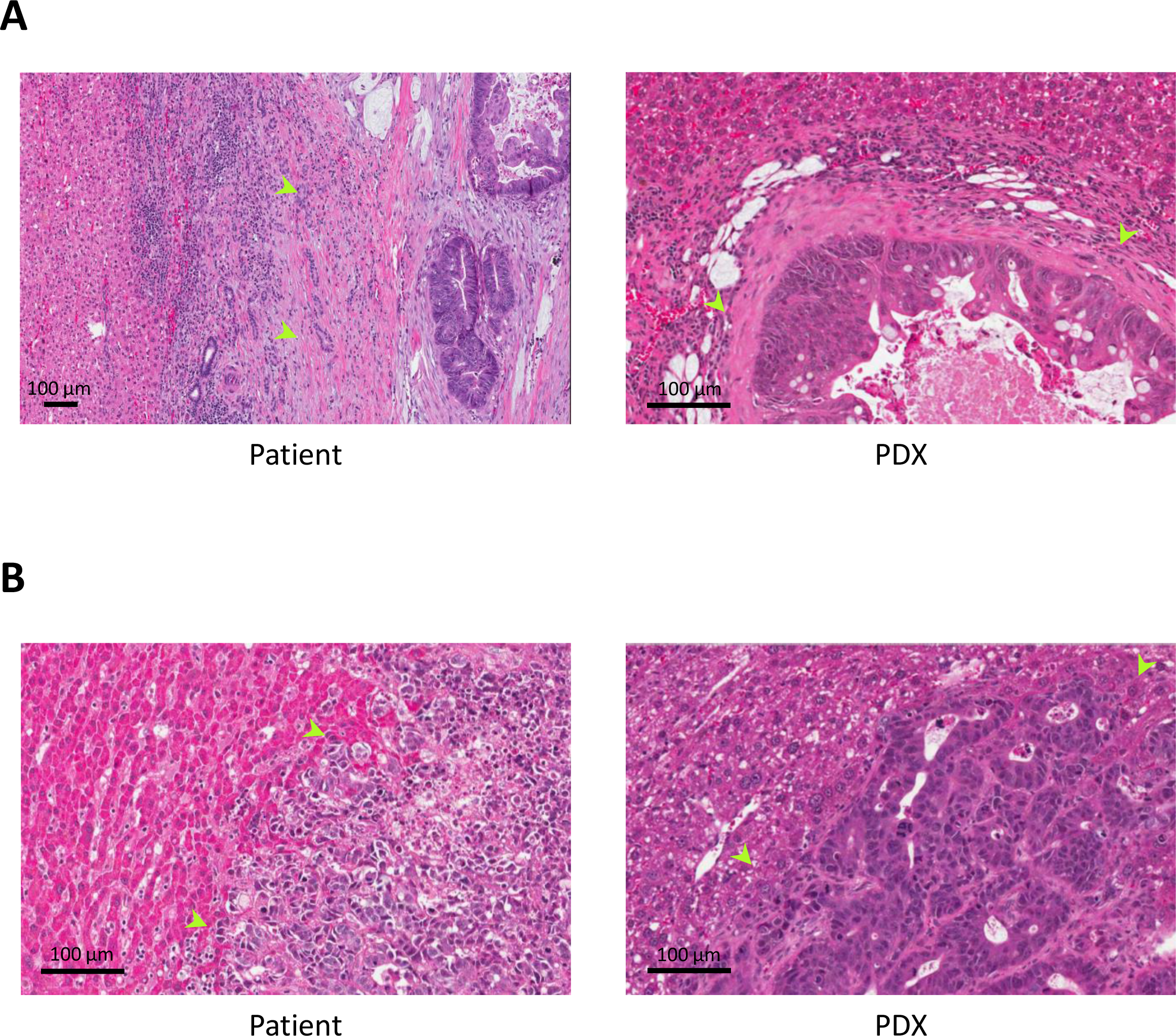
Patient-derived xenograft (PDX) mice models for CRC liver metastases with a desmoplastic and a replacement HGP (H&E images). A. Resected liver metastasis with a desmoplastic HGP (Left) and corresponding xenograft PDX- model (Right). Green arrows indicate the desmoplastic rim in the patient sample and in the liver metastasis of the mouse (PDX#35, see supplementary table 2 in Tabariès S, 2021)^16^. B. Resected liver metastasis with a replacement HGP (Left) and corresponding xenograft PDX- model (Right). Green arrows indicate some of the areas in which the cancer cells grow into the liver cell plates and contact the hepatocytes, both in the patient sample and in the liver metastasis of the mouse (PDX#30, see supplementary table 2 in Tabariès S, 2021)^16^.

### Automated scoring of HGPs of liver metastasis

An increasing number of pathology laboratories are digitising glass slides into high-resolution whole slide images (WSIs). This creates an opportunity to develop algorithms based on machine learning and artificial intelligence that can extract clinically useful information from, for example, WSIs of H&E-stained tumour sections. At least two teams have implemented this approach to score the HGPs of liver metastases in an automated way.

The algorithm developed by Qianni Zhang and her team determines the relative contribution of the replacement and of the desmoplastic HGP in a CRC liver metastasis, including the proportion of the tumour-liver interface with ‘uncertain’ HGP^20^. By combining image processing and deep learning methods, they can achieve pixel level segmentation of the tumour-liver interface. The algorithm is based on the accurate identification and segmentation of the different tissue types at this interface by using deep neural networks and by taking both cell and tissue characteristics into account. The neural network is employed to identify the tissue type using patches of a certain size. The characterization of cell types within these patches then adds sensitivity, especially at the transition of one tissue type to another. In addition, uncertain regions are classified by analysing the similarity of this region and its neighbour, a concept called ‘context-aware tissue region classification’. To train the model at the tissue level, many patches were annotated by pathologists at the Karolinska University Hospital, as belonging to liver parenchyma, fibrosis, necrosis, tumour, or inflammation. At the cell level, the model was trained by pathologists to recognize hepatocytes, cells belonging to fibrotic tissue, tumour cells and inflammatory cells. Once the algorithm succeeded in accurately classifying the tissue types of an entire WSI, rules were developed to detect the growth patterns based on the apposition of different types of tissue at the tumour-liver interface: ‘liver-fibrosis-tumour’ for the desmoplastic HGP and ‘liver-tumour’ for the replacement HGP. Extensive analytical and clinical validation is still ongoing.

The algorithm developed by Jeroen Van der Laak and his team was designed to distinguish CRLM with 100% desmoplastic HGP from liver metastases with any proportion of non-desmoplastic HGP by mimicking the visual feature extraction of an entire WSI at once, as done by pathologists^21, 22^. Due to the extensive computational power required to process the gigapixel WSIs at once, reduction of dimensionality (or compression) was necessary. This was achieved by training an encoder in a supervised way to solve several representative tasks in computational pathology. This encoder then reduced both the size and the noise level of the WSIs. In a second step, a convolutional neural network was trained using the image-level labels of ‘100% desmoplastic HGP’ and ‘any % of non-desmoplastic HGP’. When the algorithm was applied to predict the HGP of CRLM, an AUC by ROC analysis of 0.895 was obtained. The algorithm was also able to divide a cohort of 337 patients into two risk categories that predicted OS (HR: 2.35, p<0.001). It appears therefore that the HGP of liver metastases can reliably be assessed through the compression and analysis of the WSIs of H&E-stained sections and that this assessment has prognostic power.

These methods^20, 21^ demonstrate the power of automated scoring algorithms to assist the pathologist in collecting prognostic information based on parameters reflecting tumour biology. Moreover, when these computer vision algorithms can directly learn from clinical data such as survival, they will also be useful as a biomarker discovery tool^21^.

### Angiotropic extravascular migratory metastasis by pericytic mimicry

Migration of cancer cells along blood vessels at and distal to the advancing front of primary tumours and metastases has been extensively studied by the team of Lugassy and Barnhill, particularly in melanoma (for review:^33^). During this process of angiotropic extravascular migration, cancer cells are in contact with endothelial cells (‘angiotropism’) via an amorphous matrix that abundantly contains laminin and other constituents of the basement membrane, thereby replacing the pericytes (‘pericyte mimicry’). This type of extravascular migration has been proposed as an alternative to the intravascular route of metastatic spread and seems to be driven by cancer cells re-activating embryogenesis-like programs^31, 65^. In replacement-type but not in desmoplastic liver metastases of melanoma, individual cancer cells can be observed in the liver parenchyma disconnected and at a distance from the tumour-liver interface (Figure 6). As such, growth of liver metastases in a replacement pattern and extravascular migration by angiotropism and pericytic mimicry can be regarded as complementary processes representing a continuum of cancer progression with likely common underlying biological mechanisms. To accurately detect extravascular migration of individual cancer cells in liver metastases with a replacement growth pattern, immunohistochemical staining with cancer cell-specific markers is necessary. Studies that quantify the extent of this angiotropic extravascular migration in liver metastases are ongoing. It will be important to determine whether the presence of angiotropic extravascular migration in liver metastases with a replacement HGP contributes to a poorer outcome.

**Figure 6.**
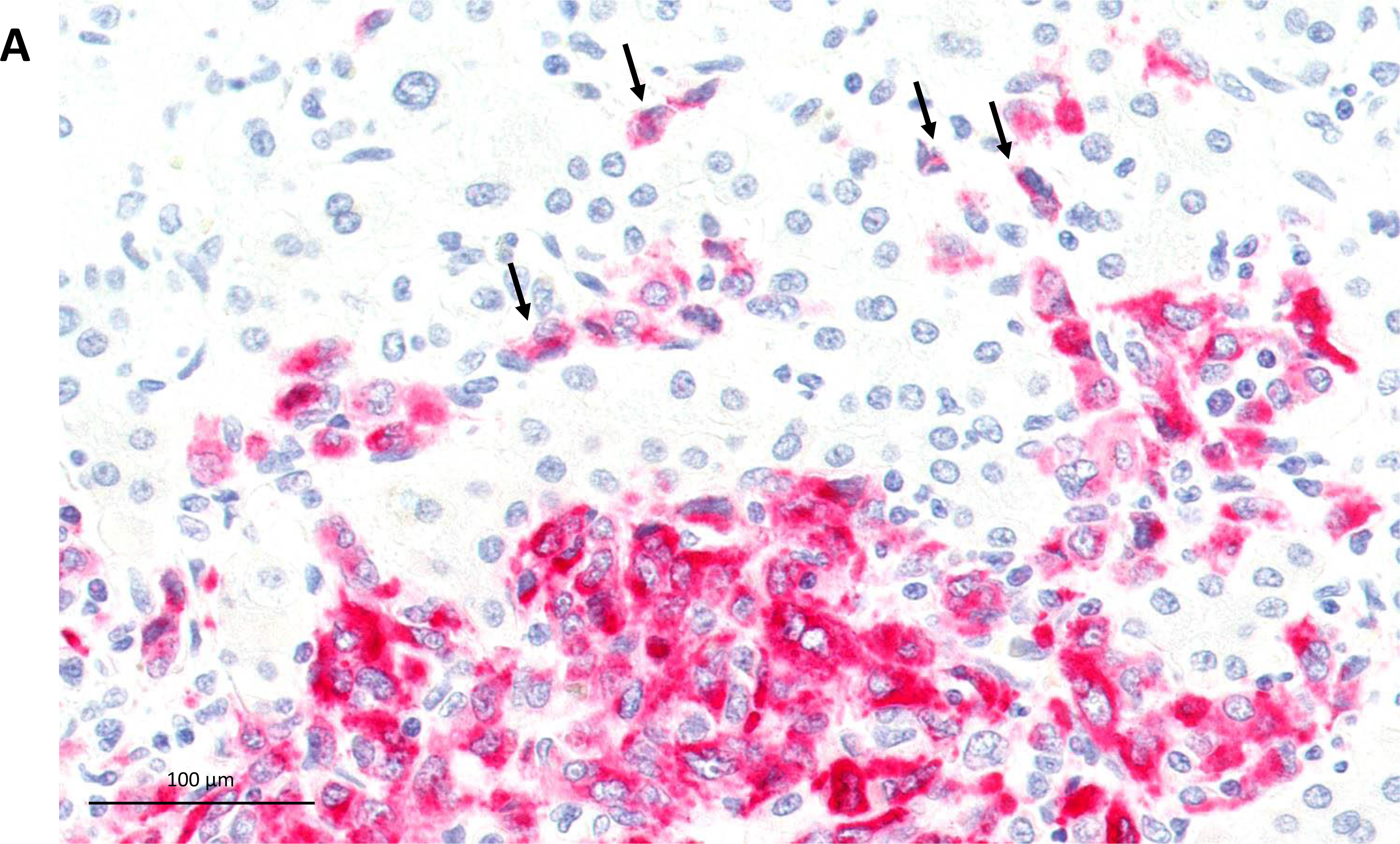
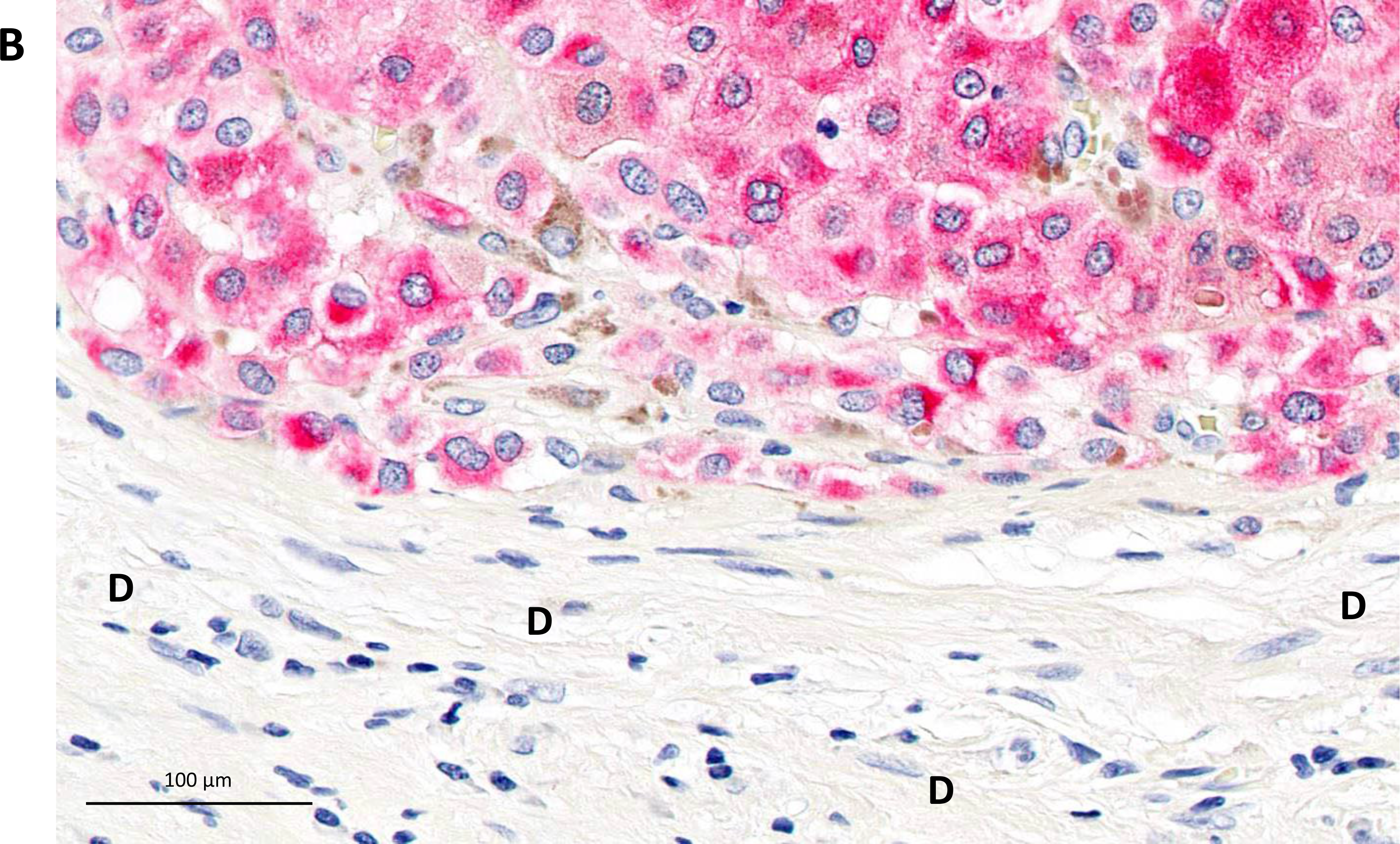
Images of melan-A immunostaining of melanoma liver metastases. (A) High magnification images of the tumour-liver interface of a melanoma liver metastasis with a replacement HGP. Small groups of melanoma cells and individual melanoma cells have migrated away from the tumour-liver interface (arrows). (B) High magnification images of the tumour-liver interface of a melanoma liver metastasis with a desmoplastic HGP. No migration of melanoma cells in the desmoplastic rim, marked by ‘D’.

### Medical imaging as a tool to identify the HGPs of liver metastases

The implementation of the HGPs in clinical practice will depend, in part, on creating the means for recognising the growth patterns without the need for surgical removal of the liver metastases and analysis by a pathologist. Medical imaging may be a promising approach to solve this challenge. Indeed, several smaller studies suggest that CT and MRI images contain information about the growth pattern (see Table 1 of previous guidelines manuscript by van Dam P et al (2017)^1^ and of the current guidelines)^66^. This is not surprising, given the major histological and biological differences between the desmoplastic and replacement growth pattern. It is, however, only during the last few years that two teams have attempted to identify growth patterns of liver metastases by medical imaging in a more systematic manner. In Erasmus MC, Rotterdam, Starmans and colleagues have extracted more than 500 radiomics features from CT-images of 76 patients with 93 CRC liver metastases with pure desmoplastic (48%) or pure replacement (52%) HGP^10^. Importantly, these features were extracted from entire metastases, not only from the lesion boundaries. A decision model based on the selection of relevant features and classification of these features by machine learning had a mean area under the curve of 0.69. Adding clinical information to the model did not improve the power to predict the HGPs. Obviously, future studies will have to include metastases with a mixed HGP. Nevertheless, this study is a valuable proof of concept for the utility of this approach.

A team at the Peking University People’s Hospital has recently published three studies on the identification of HGPs of CRC liver metastases by medical imaging^8, 9, 67^. It is important to note that these studies attempt to identify the predominant growth pattern. Cheng and colleagues^8^ analysed contrast-enhanced CT-images of 126 CRC liver metastases, of which 68 had a predominant (>50%) desmoplastic HGP and 58 had a predominant replacement HGP. Pre-contrast and post-contrast CT-images (from both the arterial and portal venous phases) were used. Of each of these 3 phases, 20 radiomics features were selected by an algorithm based on minimal redundancy and maximal relevance. A fused decision-tree based signature of the three phases resulted in a predictive model with an area under the curve of 0.94. Adding clinical information or qualitative information provided by the radiologist did not improve the predictive power.

In a similar study, MRI-derived regions, both covering the whole tumour volume as well as the tumour-liver interface specifically, were subjected to radiomic feature extraction in a cohort of 182 CRC liver metastases, of which 59 had a predominant (>50%) desmoplastic HGP and 123 had a predominant replacement HGP^9^. The predictive model that combined clinical characteristics, qualitative imaging data generated by the radiologist and radiomic feature data from the tumour-liver interface had and area under the curve of 0.91.

In their most recent study, the team at the Peking University People’s Hospital has used their CT-based radiomics HGP-signature to predict response and PFS in a cohort of 119 patients with liver metastatic CRC treated with a combination of chemotherapy and bevacizumab^67^. Among 346 metastases studied, 206 had a radiological predominant desmoplastic HGP and 140 had a radiological predominant replacement HGP. Patients with only metastases with a predominant desmoplastic HGP only as assessed by radiology had a significantly improved 1- year PFS (HR = 0.34; p<0.001).

Although the studies by Cheng J et al (2019)^8^, Han Y et al (2020)^9^ and Wei S et al (2021)^67^ are very promising, validation of the results in larger cohorts by independent research teams and with images acquired in different hospital is still necessary. In addition, at least for patients with CRLM, it will be necessary to select, by means of imaging, those patients who have metastases with a 100% desmoplastic growth pattern. So, even though considerable progresses have been made to better determine the HGP prior to resection of the liver metastases, there might still be a need to develop computational tools to integrate as many parameters as feasible to stratify patients more accurately.

## Biology

### New biological insights into growth patterns through immunohistochemical analyses

Why does a liver metastasis in one patient develop a desmoplastic rim, while a metastasis in a different patient has a replacement-type growth pattern, even when the primary tumour type is the same? The full answer to this question and the biological mechanisms that underlie the different growth patterns remain elusive. There are reasons to assume that cancer cell motility and differentiation^41^, angiocrine signals^68^ and interactions of cancer cells with hepatocytes^16^ and with stromal and inflammatory cells^32^ are important factors regulating the emergence of a distinct growth pattern. However, the precise mechanisms and the order of events leading to the specific growth patterns remain unclear. There are compelling observations to suggest that systemic treatment can alter the growth pattern^41, 61^. Also, given that some mouse PDX models can recapitulate the pattern observed in the donor patient, the growth pattern may be, at least in part, determined by cancer cell intrinsic properties^16^. However, this does not exclude epigenetic control and the influence of tumour microenvironment as important further mechanisms^52^.

Based on immunohistochemical stainings performed by the Karolinska team (Carlos Fernández Moro, Marco Gerling, Béla Bozóky) to map the spatial relationships and phenotypic states of epithelial and stromal cells, we propose two additional working hypotheses to explain the biology of the HGPs.

A first working hypothesis is that the replacement growth pattern is the default pattern of growth for cancer cells forming a tumour in the liver. This means that spontaneous or induced transition to the desmoplastic pattern regularly takes place as a second step. An intrinsic and important limitation of determining growth patterns by histological analysis of a resection specimen is that we only get information from a single time point. A non-invasive method to assess the HGPs, such as imaging, would allow longitudinal, repeated determination of HGPs. We may, however, be able to infer information about the history of a liver metastasis by comparing the centre of the tumour with its periphery. Surprisingly, after immunohistochemical analysis, we found remnants of portal triads (branches of the bile duct and of the hepatic artery) in the centre of both replacement and desmoplastic metastases. These portal elements are regularly found to be embedded in specialized portal-type stromal cells expressing Nerve Growth Factor Receptor (NGFR)- and alpha Smooth Muscle Actin (alpha-SMA, Figure 7A). This observation supports a model in which the metastatic tumour co-opts the sinusoidal blood vessels and the portal tract architecture of the liver, a mode of growth that likely is advantageous, both for blood supply and structural support. While portal triad co-option is readily identifiable at the tumour-liver interface of replacement-type liver metastases, it may be more subtle in the fibrous rim of the desmoplastic type, where pre-existing liver structures appear atrophic and attenuated. Here, immunohistochemistry can be used to identify atrophic remnants of the portal triad. Together, this leads us to propose the hypothesis that replacement growth, in most cases, precedes desmoplastic growth in metastases with the latter HGP. The time point at which the growth patterns may switch and the factors responsible for the proposed conversion remain unclear. There are other observations to support a model in which replacement growth is the default growth pattern of liver metastases. For example, when cancer cells spread within the bile ducts, the cancer cells rest on the basement membrane of the normal biliary epithelial lining and progress by replacing these normal cells and by co-opting the subepithelial stroma (Figure 7B). In addition, we occasionally observe bile ducts as part of the ductular reaction in the desmoplastic rim, in which cancer cells create hybrid cancer cell-cholangiocytes ductular structures (Figure 7C). Although these histological observations need further validation and quantification, they do support other observations consistent with growth pattern plasticity. Indeed, resistance to chemotherapy can coincide with a switch to the replacement HGP^41, 59^, while pre-operative chemotherapy converts metastases in some patients from replacement to desmoplastic HGP^61^. Also, during disease progression in patients with recurrent colorectal liver metastases, there is an evolution towards the more aggressive replacement HGP, as observed by analysing repeated resections^18^.

**Figure 7.**
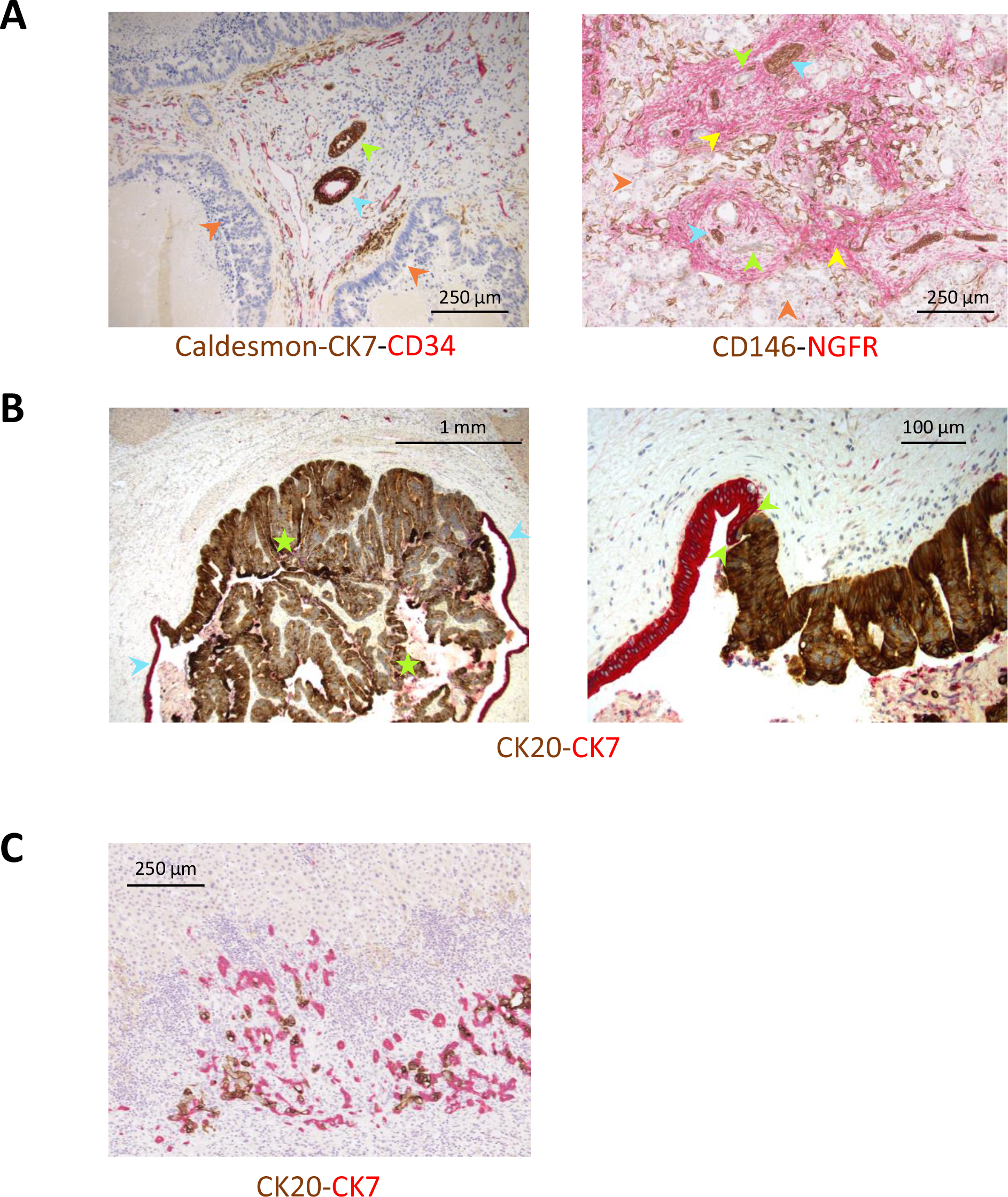
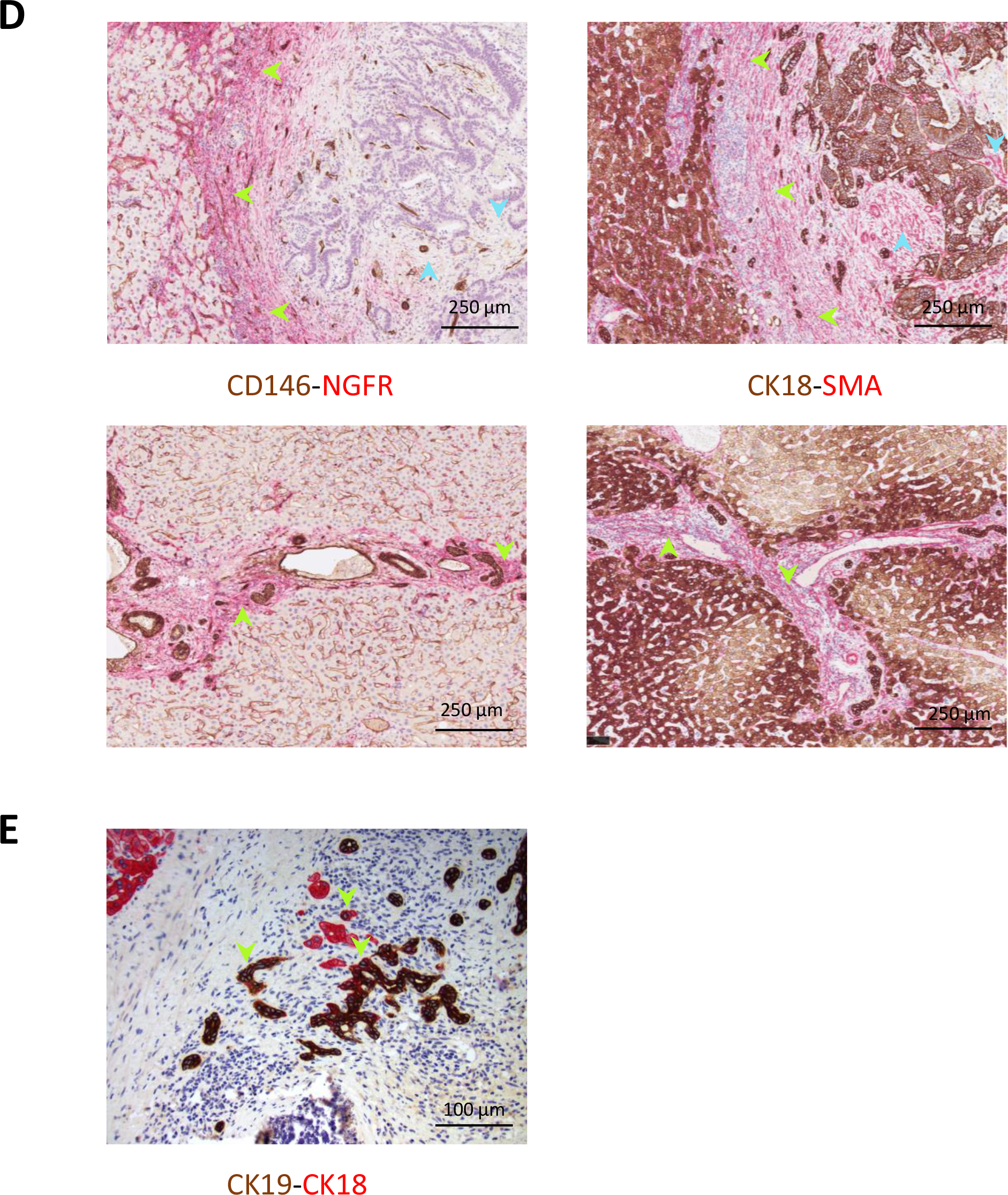
New biological insights into growth patterns through immunohistochemical analyses A. Remnants of portal zones in the centre of colorectal liver metastases. Left. Detail of a tumour centre in a metastasis with a predominant replacement HGP showing remnant of a portal zone with bile duct (green arrowhead) and hepatic artery branch (blue arrowhead). Note colonization by viable cancer cells of the periportal limiting plate region (orange arrowhead). Caldesmon (DAB, brown) stains smooth muscle cells, mainly in the media layer of the hepatic artery. CK7 (DAB, brown) stains bile duct epithelium. CD34 (AP, red) stains the endothelium of the hepatic artery and of the stromal capillary network. Right. Tumour centre in a metastasis with a desmoplastic HGP showing multiple remnants of portal zones between lobules that have undergone complete replacement by cancer cells (orange arrowheads). The bile ducts (green arrows) and branches of the hepatic artery (blue arrows) are embedded in NGFR+ portal stroma (yellow arrowheads). CD146 (DAB, brown) stains smooth muscle cells (mainly in the wall of hepatic arteries) and areas of ductular reaction. NGFR (AP, red) stains activated portal fibroblasts and stellate cells. B. Intrabiliary tumour growth in a CRC liver metastasis. Left. Densely packed cancer cells (green stars) show exophytic growth and fill the bile duct lumen. Portions of preserved biliary epithelium (blue arrows) are still identified. Right. Detail illustrating the replacement-like growth of cancer cells, which progress by establishing direct contact with and replacing the cholangiocytes while co-opting their basal membrane. CK20 (DAB, brown) stains colorectal cancer cells. CK7 (AP, red) stains cholangiocytes. C. Hybrid cancer cell-cholangiocyte ductular structures. Ductular reaction in the desmoplastic rim with cancer cells (CK20-positive, DAB, brown) forming hybrid structures with cholangiocytes (CK7-positive, AP, red). D. Stromal cell heterogeneity in a metastasis with a desmoplastic HGP. Top. The outer region of the desmoplastic rim stains strongly positively for NGFR (Left, green arrows) and α-smooth muscle actin (alpha-SMA) (Right, green arrowheads), consistent with activated portal/stellate cell stroma. In contrast, the stroma in the metastasis centre is positive for alpha-SMA but negative for NGFR, indicating a desmoplastic character (Left and right, blue arrows). Bottom. Reference illustrations of activated portal stroma in non-neoplastic liver, showing (Left) NGFR and (Right) alpha-SMA immunoreactivity (Left and right, green arrows). CD146 (DAB, brown) stains vascular and sinusoidal endothelium and smooth muscle in branches of the hepatic artery and portal vein. NGFR (AP, red) stains activated portal fibroblasts and stellate cells. CK18 (DAB, brown) stains hepatocytes and cholangiocytes. Alpha-SMA (AP, red) stains activated portal fibroblasts, stellate cells and desmoplasia-associated myofibroblasts. E. Ductular reaction in the desmoplastic rim with cells with a hepatocyte-like (CK18-positive, AP, red) and a cholangiocytes-like (CK18, DAB, brown) phenotype (green arrows).

A second working hypothesis is that the fibrous rim surrounding desmoplastic liver metastases and the portal tract are biologically related. This hypothesis is supported by two observations. *Firstly*, the stromal cells of the desmoplastic rim, and especially of the outer portion of the rim neighbouring the surrounding liver parenchyma, strongly co-express NGFR and alpha-SMA, indicative of a “myofibroblast” or “activated fibroblast” phenotype (Figure 7D). NGFR is expressed by progenitors of Ito/stellate cells and of portal fibroblasts in the foetal liver^69, 70^ and this receptor also plays a crucial role during pathological liver fibrosis by inducing fibrogenic gene expression, for example of the Transforming Growth Factor beta1-gene, in activated (myo)fibroblasts^71–73^. *Secondly*, by co-immunostaining for CK18, as a marker of hepatocytes, and CK19, as a marker of cholangiocytes, we often observe mosaic ductular structures in the desmoplastic rim composed of a mixture of cells with a hepatocyte-like and a cholangiocyte-like phenotype (Figure 7E). Activated fibroblasts are known to induce cholangiocyte differentiation in hepatic stem-like cells (for example, via Jagged-1 and Hedgehog ligands) and this process partly relies on NGFR expression in the activated liver fibroblasts^73, 74^. NGFR-expressing and activated, alpha-SMA-positive fibroblasts in the desmoplastic rim may therefore activate extracellular matrix production and induce a ductular reaction by engaging bipotent progenitors, resembling portal tract development as well as liver fibrosis in other pathological conditions involving liver injury^75^. In the metastasis context, destruction of liver cells by the invading tumour, inflammation, and damage of the peritumoral liver tissue are potential mechanisms of liver injury.

### Hypotheses to explain the biology of the distinct histopathological growth patterns

There is currently no satisfactory explanation for the specific biology of each of the histopathological growth patterns. Table 6 therefore summarizes some hypotheses to explain the distinct phenotypes of the desmoplastic and replacement growth patterns. These hypotheses are derived from histopathological insights, pre-clinical animal models and the comparison with organ development in the embryo. The hypotheses listed in Table 6 are not mutually exclusive and elements of each probably contribute to the specific growth patterns of liver metastases. In addition, some hypotheses outlined only address individual growth patterns.

**Table 6.** Hypotheses to explain the biology of the different histopathological growth patterns of liver metastases, including supporting evidence and/or supporting arguments.

| Hypotheses to explain the biology of the different HGPs of liver metastases | Supporting evidence and/or supporting argument |
| --- | --- |
| **Site of implantation**<br>The site of cancer cell implantation in the liver determines the HGP | In animal models of liver metastasis (where cancer cells were introduced either via an arterial route or a portal route) the arterial route gave rise to a significantly higher proportion of desmoplastic metastases, originating from within portal tracts, than when cancer cells entered the liver via the vena portae, which more often resulted in a replacement-type liver metastases (Paku S & Lapis K, 1993<sup>123</sup>; Vidal-Vanaclocha F, 2008<sup>124</sup>). |
| **Revertant *in situ* growth**<br>The replacement HGP is a reversion to *in situ* growth of cancer cells (growth within the boundaries of a basement membrane) | Cancer cells in replacement-type liver metastases take the place of hepatocytes and rest on the Space of Disse. The hybrid ductular structures (cancer cell-cholangiocyte) and growth within bile ducts are other examples of *in situ* growth of cancer cells in the liver. Revertant *in situ* growth has been described in lymph node metastases of cancer which adopt a similar growth pattern with cancer cells replacing lymphocytes and co-opting the vasculature (Barsky SH, 1997<sup>125</sup>). |
| **Coagulation and inflammation**<br>The presence (desmoplastic) or absence (replacement) of coagulation and inflammation determine the HGPs | Angiogenesis, coagulation, inflammation, and fibrosis are interrelated processes during wound healing and may also be the driving force behind the desmoplastic HGP of liver metastases. When cancer cells can avoid activating any of these processes, liver metastases can adopt the replacement HGP. Only minimal fibrin deposits and hypoxia, one of the factors inducing |
|  | angiogenesis, occur in liver metastases with a replacement HPG (Stessels F, 2004<sup>38</sup>) and replacement pattern liver metastases often show an ‘immune desert’ (Stremitzer S, 2020<sup>2</sup>). |
| **Response to liver injury**<br>The HGPs reflect the response patterns of the liver to injury, with the desmoplastic HGP resembling biliary liver fibrosis and the replacement pattern resembling liver regeneration. | There are two responses to liver injury – the fibrotic response and the liver regeneration response (Ding B, 2014<sup>82</sup>). The desmoplastic rim contains a portal-type of stroma (this manuscript) and proliferating bile ducts (ductular reaction) which resembles the fibrotic response to liver injury (Schuppan D, 2013<sup>126</sup>). In replacement-type liver metastases, the cancer cells are arranged in cell plates and replace the parenchymal hepatocytes, thereby preserving the vascular architecture of the liver parenchyma, which resembles morphologically progenitor cell-driven liver regeneration (Deszo K, 2012<sup>127</sup>). |
| **Transcriptional reprogramming**<br>The HGPs are the result of transcriptional reprogramming driven by an HGP-specific epigenetic landscape. | CRC cells have been shown to express liver-specific genes in liver metastases, thereby losing expression of colon-specific genes. This reprogramming is driven by a change in enhancer-regions in the genome (Teng S, 2020<sup>52</sup>). In the replacement HGP, cancer tissue mimics liver tissue histologically, supporting the hypothesis that cancer cells switch on a liver organogenesis program that may be driven by the sinusoidal endothelial cells as in vascularizing organogenesis (Matsumoto K, 2001<sup>80</sup>; Crivellato E, 2007<sup>81</sup>; Ding B, 2014<sup>82</sup>; Daniel E, 2019<sup>83</sup>). Desmoplastic liver metastases histologically resemble the primary colorectal tumour and may also have a similar transcriptional profile. |
| **Cancer cell motility** |  |
| The ability of cancer cells to move and migrate determines the HGPs because cancer cell motility is necessary for the replacement HGP. | Knocking down of *ARPC3*, a gene coding for a subunit of an actin nucleating complex necessary for cell motility, or *RUNX1*, coding for Runt Related Transcription Factor-1 (which is upstream of ARP2/3) changes the HGP from a replacement pattern to a desmoplastic pattern in an animal model of liver metastasis (Frentzas S, 2016<sup>41</sup>; Rada M, 2021<sup>111</sup>). |
| **Replacement HGP is the default**<br>The replacement HGP is the default growth pattern. | Remnants of co-opted portal triads are present in the centre of liver metastases, independent of the HGP. This suggests that desmoplastic liver metastases originate from replacement-type metastases, given that co-option of portal triads is not observed at the interface with the liver in desmoplastic liver metastases. Cancer cells also replace normal epithelial cells when they spread within a bile duct or form hybrid structures with cholangiocytes of a ductular reaction (this manuscript), supporting the idea the cancer cells have a natural tendency to interact with normal cells. What induces the transition from replacement to desmoplastic growth is still unknown. |
| **Angiotropic extravascular migration and pericyte mimicry**<br>The replacement HGP relies on these processes. | Both the growth along sinusoidal blood vessels via angiotropic migration and pericyte mimicry and the histological resemblance of replacement liver metastases to liver parenchyma suggest that programs of embryogenesis are active in this type of metastases (Lugassy C, 2020<sup>33</sup>). |

Taken together, cancer cells within a liver metastasis exhibiting a replacement growth pattern appear to adapt to the microenvironment of the liver parenchyma and may therefore be sensitive to a liver pro-metastatic reaction of the patient^76^, while cancer cells of a desmoplastic metastasis create their own microenvironment. Against this background, it could be argued that cancer cells in a replacement metastasis behave like hepatocytes or hepatocyte progenitor cells communicating with the liver niche (for example with the co-opted sinusoidal endothelial cells), whereas cancer cells in a desmoplastic metastasis more autonomously form a tumour that resembles the primary tumour. The plasticity of the growth patterns suggested by clinical observations appears to indicate that this divergent behaviour of cancer cells in the liver is not, or at least not entirely, the result of a different mutational gene profile, but rather of epigenetic events and the ability to respond to stimuli from the microenvironment, such as soluble factors elicited by the liver pro-metastatic reaction^76^ and liver immune responses^77^.

## Discussion

Since the publication of the first consensus guidelines^1^, numerous studies have been conducted describing the impact of HGPs on the outcome of patients with liver metastases (Table 1). These studies are not limited to liver metastases from colorectal carcinoma, but also include patients with liver metastases from breast carcinoma, melanoma, and pancreatic adenocarcinoma^5, 42, 44, 45^. The association between replacement HGP and poorer patient outcome, independent of the primary tumour type, has been confirmed by these new studies. A new cut-off to categorize patients with CRLM according to the HGPs is presented in the current guidelines. This cut-off is derived from the observation in a large multi-centre cohort of patients that any proportion of non-desmoplastic HGP, however small, is associated with a worse prognosis. The extent of non-desmoplastic features within the metastases in itself does not seem to modulate outcome any further. We have therefore updated the guidelines for scoring the HGPs of CRLM for the purposes of prognostication of patients, and we propose herein some immunohistochemical assays that may help to identify the growth patterns in more challenging situations, such as in the presence of dense inflammation or systemic treatment effects.

The tumour-type independent prognostic value of the HGPs fuels the idea that the biology of the replacement HGP is fundamentally different from that of the desmoplastic growth pattern. Some of these differences have been well described. In the desmoplastic growth pattern, a dense immune-inflammatory cell infiltrate surrounds the fibrous rim, while the replacement growth pattern has the characteristics of an immune desert, especially when no chemotherapy is involved^2–4, 32, 78^. The desmoplastic pattern has angiogenic vascular hot spots in between cancer cell nests and hypoxic areas while the replacement growth pattern shows a uniformly high vessel density and minimal hypoxia, probably because of efficient vessel co- option^30, 38, 41^. Consequently, replacement-type liver metastases are also metabolically more active than desmoplastic liver metastases, as demonstrated by FDG-PET analyses^79^.

A striking morphological difference between the growth patterns lies in the organisation of the cancer cells and the interaction with the host liver tissue. In replacement-type liver metastases, cancer cells mimic hepatocytes by an arrangement in solid cell plates in between the co-opted sinusoidal blood vessels. This type of growth clearly resembles the ‘vascularizing organogenesis’ that takes place when the liver develops in the embryo and may also be guided by instructive signals originating in the liver sinusoidal endothelial cells^80–83^. Accordingly, cancer cells belonging to replacement-type liver metastases seem to hijack the embryological program of liver development with the resulting tumour adopting the histological architecture of liver tissue. The work of Teng and team^52^ supports this hypothesis. They have shown that CRLM, when compared with primary colorectal cancer, simultaneously gain liver-specific and lose colon-specific transcription programs. They also showed that this is the result of a reprogrammed enhancer landscape. Enhancers are regulatory elements in the genome that are influenced by the environment and, as such, play an important role in tissue-specific gene expression patterns and cell identity. However, whether differences in the enhancer landscape can also explain the morphological differences between the replacement and the desmoplastic growth pattern of liver metastases still needs to be investigated. During desmoplastic growth of liver metastases, the cancer cells arrange in more differentiated structures, not as cancer cell plates, and resemble the glandular structures of primary colorectal and breast cancer. In other words, desmoplastic liver metastases morphologically mimic the primary tumour they originate from, where cancer cells typically induce a continuous wound-healing response with inflammation, fibrosis, coagulation, and angiogenesis. This probably involves tumour-host interactions that are active in the primary tumour and epithelial-stromal interactions of the normal tissue counterpart (e.g., colon, breast, etc.). These hypothetical and morphology-driven views on the divergent biological mechanisms of the liver metastasis growth patterns are now being investigated by bulk RNA-sequencing, single cell RNA-sequencing, in situ RNA-sequencing and multiplex immunohistochemistry. PDX-models and co-organoids derived from patient liver metastases are used for functional validation. Alternative hypotheses to explain the distinct histopathological growth patterns have been listed in Table 6.

At a single time-point, patients often have liver metastases consisting of both desmoplastic and replacement HGP regions. This is, for example, true for about 60% of all patients with resected CRLM^18^, independent of whether chemotherapy was administered before surgery. Co-occurrence of distinct HGPs thus seems to be part of the growth process of liver metastases and this may be the consequence of transitioning from one HGP to another. We now propose the working hypothesis, based on immunohistochemical analyses, that the replacement HGP is the default growth pattern of liver metastases. Although there are data to support the view that pre-surgery chemotherapy can induce desmoplastic growth in some metastases^18, 61^ and that a switch to replacement growth can occur upon resistance to systemic treatment^41^, the cellular and molecular mechanisms responsible for these transitions from one growth pattern to another remain to be elucidated. What these and other findings do seem to suggest is that epigenetic processes drive the growth patterns rather than mutational hardwiring. Recently, the concept of ‘histostasis’, driven by cancer cell-autonomous properties, has been put forward to explain the morphological resemblance between metastatic tissue and the corresponding primary tumour^84^. As a complement, we propose here to introduce the concept of ‘histokinesis’, a process driven by cancer cell-responsiveness to instructive host tissue-derived signals, such as the pro-metastatic liver reaction^76^, to explain the clear morphological differences between the primary tumour and, for example, replacement-type liver metastases. This is probably a more general biological concept, given the observations of similar growth patterns in, for example, lung metastases^56, 85–87^.

The plasticity of the growth patterns might be exploited in future therapeutic strategies. A prerequisite to feasibility would be a continuous evaluation of the growth pattern in a pre-surgical setting of systemic treatment. This implies a reliable non-invasive method for repeatedly identifying the growth patterns during the patient treatment. Table 1 highlights the initiatives of several teams worldwide to develop algorithms to assess the growth patterns of liver metastases by medical imaging^8–10, 66^. In addition, several studies are still ongoing with results to be expected in the coming years. As an alternative, circulating markers in the blood of patients may be useful to identify the prevailing growth pattern at a certain moment in time. A study by Tabariès^16^ proposes exosome-derived claudin-profiling as a tool to predict the growth pattern of CRLM.

The role of systemic treatment, either neo-adjuvant or adjuvant, for patients with a priori resectable metachronous CRLM remains unclear. In many countries, patients will receive standard post-operative chemotherapy, following metastasectomy performed with curative intent. Although the benefit of adjuvant treatment is still to be fully appreciated, surgery alone is often not considered. To face the problem of potentially low accrual in a study that compares surgery alone with surgery combined with adjuvant chemotherapy, we suggest limiting the study population to those patients with liver metastases that exclusively have the desmoplastic growth pattern upon careful pathological evaluation of the resected metastases, as a first approach. Alternatively, and only when a non-invasive pre-operative marker of the HGPs becomes available (as liver biopsies to evaluate the HGP are not suitable), a window of opportunity study could be envisioned to examine the role of specific treatments for replacement and desmoplastic liver metastases in patients with (borderline) resectable liver metastases. For example, given the distinct immune contexture of each of the growth patterns, the choice of immunotherapy may need to be adapted to the growth pattern of the liver metastases. It is also unclear whether the clinically relevant systemic immunosuppressive effects of the presence of liver metastases, leading to reduced benefit from immune checkpoint inhibitors, may be growth pattern-dependent^88^. A better insight into the interaction of liver-metastatic cancer cells with the complex immune environment of the liver will contribute to understanding the biology of the HGPs^77^.

In conclusion, we provide updated guidelines for scoring the histopathological growth patterns of liver metastases. These are of importance not only to implement the HGPs in the clinical care of patients with liver metastatic cancer, but also to properly conduct studies that seek to identify the biological basis for these growth patterns. The latter is important to better understand the heterogeneity of liver metastases, and thus perhaps also of tumour expansion in other organs where similar growth patterns have been described, such as in the lungs^56, 87^.

## Data Availability

All data produced in the present study are available upon reasonable request to the authors.

## Acknowledgement

The current guidelines are the result of numerous discussions at the annual meetings of the international Liver Metastasis Research Network.

## References

1. van Dam, P.J., van der Stok, E.P., Teuwen, L.A., Van den Eynden, G.G., Illemann, M., Frentzas, S., et al. International consensus guidelines for scoring the histopathological growth patterns of liver metastasis. Br J Cancer 117, 1427–1441 (2017).

2. Stremitzer, S., Vermeulen, P., Graver, S., Kockx, M., Dirix, L., Yang, D. et al. Immune phenotype and histopathological growth pattern in patients with colorectal liver metastases. Br J Cancer 122, 1518–1524 (2020).

3. Liang, J.Y., Xi, S.Y., Shao, Q., Yuan, Y.F., Li, B.K., Zheng, Y. et al. Histopathological growth patterns correlate with the immunoscore in colorectal cancer liver metastasis patients after hepatectomy. Cancer Immunol Immunother 69, 2623–2634 (2020).

4. Hoppener, D.J., Nierop, P.M.H., Hof, J., Sideras, K., Zhou, G., Visser, L. et al. Enrichment of the tumour immune microenvironment in patients with desmoplastic colorectal liver metastasis. Br J Cancer 123, 196–206 (2020).

5. Watanabe, K., Mitsunaga, S., Kojima, M., Suzuki, H., Irisawa, A., Takahashi, H. et al. The “histological replacement growth pattern” represents aggressive invasive behavior in liver metastasis from pancreatic cancer. Cancer Med 9, 3130–3141 (2020).

6. Messaoudi, N., Henault, D., Stephen, D., Cousineau, I., Simoneau, E., Rong, Z. et al. Prognostic implications of adaptive immune features in MMR-proficient colorectal liver metastases classified by histopathological growth patterns. Br J Cancer (2022).

7. Gulia, S., Khurana, S., Shet, T. & Gupta, S. Radiographically occult intrasinusoidal liver metastases leading to hepatic failure in a case of breast cancer. BMJ Case Rep 2016, (2016).

8. Cheng, J., Wei, J., Tong, T., Sheng, W., Zhang, Y., Han, Y. et al. Prediction of Histopathologic Growth Patterns of Colorectal Liver Metastases with a Noninvasive Imaging Method. Ann Surg Oncol 26, 4587–4598 (2019).

9. Han, Y., Chai, F., Wei, J., Yue, Y., Cheng, J., Gu, D. et al. Identification of Predominant Histopathological Growth Patterns of Colorectal Liver Metastasis by Multi-Habitat and Multi-Sequence Based Radiomics Analysis. Front Oncol 10, 1363 (2020).

10. Starmans, M.P.A., Buisman, F.E., Renckens, M., Willemssen, F., van der Voort, S.R., Groot Koerkamp, B. et al. Distinguishing pure histopathological growth patterns of colorectal liver metastases on CT using deep learning and radiomics: a pilot study. Clin Exp Metastasis 38, 483–494 (2021).

11. Alzubi, M.A., Sohal, S.S., Sriram, M., Turner, T.H., Zot, P., Idowu, M. et al. Quantitative assessment of breast cancer liver metastasis expansion with patient-derived xenografts. Clin Exp Metastasis 36, 257–269 (2019).

12. Piquet, L., Dewit, L., Schoonjans, N., Millet, M., Berube, J., Gerges, P.R.A. et al. Synergic Interactions Between Hepatic Stellate Cells and Uveal Melanoma in Metastatic Growth. Cancers (Basel) 11, (2019).

13. Vlachogiannis, G., Hedayat, S., Vatsiou, A., Jamin, Y., Fernandez-Mateos, J., Khan, K. et al. Patient-derived organoids model treatment response of metastatic gastrointestinal cancers. Science 359, 920–926 (2018).

14. Ibrahim, N.S., Lazaris, A., Rada, M., Petrillo, S.K., Huck, L., Hussain, S. et al. Angiopoietin1 Deficiency in Hepatocytes Affects the Growth of Colorectal Cancer Liver Metastases (CRCLM). Cancers (Basel) 12, (2019).

15. Masaki, S., Hashimoto, Y., Kunisho, S., Kimoto, A. & Kitadai, Y. Fatty change of the liver microenvironment influences the metastatic potential of colorectal cancer. Int J Exp Pathol 101, 162–170 (2020).

16. Tabaries, S., Annis, M.G., Lazaris, A., Petrillo, S.K., Huxham, J., Abdellatif, A. et al. Claudin-2 promotes colorectal cancer liver metastasis and is a biomarker of the replacement type growth pattern. Commun Biol 4, 657 (2021).

17. Bartlett, A.Q., Pennock, N.D., Klug, A. & Schedin, P. Immune Milieu Established by Postpartum Liver Involution Promotes Breast Cancer Liver Metastasis. Cancers (Basel) 13, (2021).

18. Galjart, B., Nierop, P.M.H., van der Stok, E.P., van den Braak, R., Hoppener, D.J., Daelemans, S., et al. Angiogenic desmoplastic histopathological growth pattern as a prognostic marker of good outcome in patients with colorectal liver metastases. Angiogenesis 22, 355–368 (2019).

19. Hoppener, D.J., Galjart, B., Nierop, P.M.H., Buisman, F.E., van der Stok, E.P., Coebergh van den Braak, R.R.J., et al. Histopathological Growth Patterns and Survival After Resection of Colorectal Liver Metastasis: An External Validation Study. JNCI Cancer Spectr 5, pkab026 (2021).

20. Zhaoyang, X., Carlos Fernádez, M., Danyil, K., Béla, B., Le, D. & Qianni, Z., *Tissue Region Growing for Hispathology Image Segmentation*, in *Proceedings of the 2018 3rd International Conference on Biomedical Imaging*, Signal Processing. 2018, Association for Computing Machinery: Bari, Italy.

21. Tellez, D., Höppener, D., Verhoef, C., Grünhagen, D., Nierop, P., Drozdzal, M., et al. Extending unsupervised neural image compression with supervised multitask learning. in Medical Imaging with Deep Learning. 2020. PMLR.

22. Tellez, D., Litjens, G., van der Laak, J. & Ciompi, F. Neural Image Compression for Gigapixel Histopathology Image Analysis. IEEE Trans Pattern Anal Mach Intell 43, 567–578 (2021).

23. Nierop, P.M.H., Galjart, B., Hoppener, D.J., van der Stok, E.P., Coebergh van den Braak, R.R.J., Vermeulen, P.B., et al. Salvage treatment for recurrences after first resection of colorectal liver metastases: the impact of histopathological growth patterns. Clin Exp Metastasis 36, 109–118 (2019).

24. Nierop, P.M.H., Hoppener, D.J., van der Stok, E.P., Galjart, B., Buisman, F.E., Balachandran, V.P. et al. Histopathological growth patterns and positive margins after resection of colorectal liver metastases. HPB (Oxford) 22, 911–919 (2020).

25. Buisman, F.E., Van der Stok, E.P., Galjart, B., Vermeulen, P.B., Balachandran, V.P., Coebergh van den Braak, R.R.J., et al. Histopathological growth patterns as biomarker for adjuvant systemic chemotherapy in patients with resected colorectal liver metastases. Clin Exp Metastasis (2020).

26. Masson, P. Les tumeurs. Traité De Pathologie Et Thérapie Appliquées 27, 572–574 (1923).

27. Hamperl, H. Die morphologie der tumoren. Lehrbuch Der Allgemeinen Pathologie Und Der Pathologischen Anatomie 6, 243–244 (1956).

28. Elias, H. & Bouldin, R.F. Reaction of the normal liver parenchyma to metastatic carcinoma. Acta Hepatosplenol 9, 357–386 (1962).

29. Elias, H., Bierring, F. & Grunnet, I. Cellular Changes in the Vicinity of Metastatic Carcinoma, Observed by Light and Electron Microscopy. Oncologia 18, 210–24 (1964).

30. Vermeulen, P.B., Colpaert, C., Salgado, R., Royers, R., Hellemans, H., Van Den Heuvel, E. et al. Liver metastases from colorectal adenocarcinomas grow in three patterns with different angiogenesis and desmoplasia. J Pathol 195, 336–42 (2001).

31. Latacz, E., Caspani, E., Barnhill, R., Lugassy, C., Verhoef, C., Grunhagen, D. et al. Pathological features of vessel co-option versus sprouting angiogenesis. Angiogenesis 23, 43–54 (2020).

32. van Dam, P.J., Daelemans, S., Ross, E., Waumans, Y., Van Laere, S., Latacz, E. et al. Histopathological growth patterns as a candidate biomarker for immunomodulatory therapy. Semin Cancer Biol 52, 86–93 (2018).

33. Lugassy, C., Kleinman, H.K., Vermeulen, P.B. & Barnhill, R.L. Angiotropism, pericytic mimicry and extravascular migratory metastasis: an embryogenesis-derived program of tumor spread. Angiogenesis 23, 27–41 (2020).

34. Allison, K.H., Fligner, C.L. & Parks, W.T. Radiographically occult, diffuse intrasinusoidal hepatic metastases from primary breast carcinomas: a clinicopathologic study of 3 autopsy cases. Arch Pathol Lab Med 128, 1418–23 (2004).

35. Simone, C., Murphy, M., Shifrin, R., Zuluaga Toro, T. & Reisman, D. Rapid liver enlargement and hepatic failure secondary to radiographic occult tumor invasion: two case reports and review of the literature. J Med Case Rep 6, 402 (2012).

36. Watson, A.J. Diffuse intra-sinusoidal metastatic carcinoma of the liver. J Pathol Bacteriol 69, 207–17 (1955).

37. Vaideeswar, P., Munot, S., Rojekar, A. & Deodhar, K. Hepatic diffuse intra-sinusoidal metastases of pulmonary small-cell carcinoma. J Postgrad Med 58, 230–1 (2012).

38. Stessels, F., Van den Eynden, G., Van der Auwera, I., Salgado, R., Van den Heuvel, E., Harris, A.L., et al. Breast adenocarcinoma liver metastases, in contrast to colorectal cancer liver metastases, display a non-angiogenic growth pattern that preserves the stroma and lacks hypoxia. Br J Cancer 90, 1429–36 (2004).

39. Hoppener, D.J., Nierop, P.M.H., Herpel, E., Rahbari, N.N., Doukas, M., Vermeulen, P.B. et al. Histopathological growth patterns of colorectal liver metastasis exhibit little heterogeneity and can be determined with a high diagnostic accuracy. Clin Exp Metastasis 36, 311–319 (2019).

40. Terayama, N., Terada, T. & Nakanuma, Y. Histologic growth patterns of metastatic carcinomas of the liver. Jpn J Clin Oncol 26, 24–9 (1996).

41. Frentzas, S., Simoneau, E., Bridgeman, V.L., Vermeulen, P.B., Foo, S., Kostaras, E. et al. Vessel co-option mediates resistance to anti-angiogenic therapy in liver metastases. Nat Med 22, 1294–1302 (2016).

42. Bohlok, A., Vermeulen, P., Leduc, S., Latacz, E., Botzenhart, L., Richard, F. et al. Association between the histopathological growth patterns of liver metastases and survival after hepatic surgery in breast cancer patients. NPJ Breast Cancer 6, 64 (2020).

43. Horn, S.R., Stoltzfus, K.C., Lehrer, E.J., Dawson, L.A., Tchelebi, L., Gusani, N.J. et al. Epidemiology of liver metastases. Cancer Epidemiol 67, 101760 (2020).

44. Barnhill, R., Vermeulen, P., Daelemans, S., van Dam, P.J., Roman-Roman, S., Servois, V. et al. Replacement and desmoplastic histopathological growth patterns: A pilot study of prediction of outcome in patients with uveal melanoma liver metastases. J Pathol Clin Res 4, 227–240 (2018).

45. Barnhill, R., van Dam, P.J., Vermeulen, P., Champenois, G., Nicolas, A., Rawson, R.V. et al. Replacement and desmoplastic histopathological growth patterns in cutaneous melanoma liver metastases: frequency, characteristics, and robust prognostic value. J Pathol Clin Res 6, 195–206 (2020).

46. Grossniklaus, H.E., Zhang, Q., You, S., McCarthy, C., Heegaard, S. & Coupland, S.E. Metastatic ocular melanoma to the liver exhibits infiltrative and nodular growth patterns. Hum Pathol 57, 165–175 (2016).

47. Grossniklaus, H.E. Progression of ocular melanoma metastasis to the liver: the 2012 Zimmerman lecture. JAMA Ophthalmol 131, 462–9 (2013).

48. Meyer, Y., Bohlok, A., Hoppener, D., Galjart, B., Doukas, M., Grunhagen, D.J. et al. Histopathological growth patterns of resected non-colorectal, non-neuroendocrine liver metastases: a retrospective multicenter studyss. Clin Exp Metastasis (2022).

49. Fernandez Moro, C., Bozoky, B. & Gerling, M. Growth patterns of colorectal cancer liver metastases and their impact on prognosis: a systematic review. BMJ Open Gastroenterol 5, e000217 (2018).

50. Kwapisz, D. Oligometastatic breast cancer. Breast Cancer 26, 138–146 (2019).

51. Tran, C.G., Sherman, S.K., Chandrasekharan, C. & Howe, J.R. Surgical Management of Neuroendocrine Tumor Liver Metastases. Surg Oncol Clin N Am 30, 39–55 (2021).

52. Teng, S., Li, Y.E., Yang, M., Qi, R., Huang, Y., Wang, Q. et al. Tissue-specific transcription reprogramming promotes liver metastasis of colorectal cancer. Cell Res 30, 34–49 (2020).

53. Nielsen, K., Rolff, H.C., Eefsen, R.L. & Vainer, B. The morphological growth patterns of colorectal liver metastases are prognostic for overall survival. Mod Pathol 27, 1641–8 (2014).

54. Eefsen, R.L., Vermeulen, P.B., Christensen, I.J., Laerum, O.D., Mogensen, M.B., Rolff, H.C., et al. Growth pattern of colorectal liver metastasis as a marker of recurrence risk. Clin Exp Metastasis 32, 369–81 (2015).

55. Kuczynski, E.A., Yin, M., Bar-Zion, A., Lee, C.R., Butz, H., Man, S. et al. Co-option of Liver Vessels and Not Sprouting Angiogenesis Drives Acquired Sorafenib Resistance in Hepatocellular Carcinoma. J Natl Cancer Inst 108, (2016).

56. Bridgeman, V.L., Vermeulen, P.B., Foo, S., Bilecz, A., Daley, F., Kostaras, E. et al. Vessel co-option is common in human lung metastases and mediates resistance to anti- angiogenic therapy in preclinical lung metastasis models. J Pathol 241, 362–374 (2017).

57. Leenders, W.P., Kusters, B., Verrijp, K., Maass, C., Wesseling, P., Heerschap, A. et al. Antiangiogenic therapy of cerebral melanoma metastases results in sustained tumor progression via vessel co-option. Clin Cancer Res 10, 6222–30 (2004).

58. Martens, T., Laabs, Y., Gunther, H.S., Kemming, D., Zhu, Z., Witte, L. et al. Inhibition of glioblastoma growth in a highly invasive nude mouse model can be achieved by targeting epidermal growth factor receptor but not vascular endothelial growth factor receptor-2. Clin Cancer Res 14, 5447–58 (2008).

59. Mentha, G., Terraz, S., Morel, P., Andres, A., Giostra, E., Roth, A. et al. Dangerous halo after neoadjuvant chemotherapy and two-step hepatectomy for colorectal liver metastases. Br J Surg 96, 95–103 (2009).

60. Rubbia-Brandt, L., Giostra, E., Brezault, C., Roth, A.D., Andres, A., Audard, V. et al. Importance of histological tumor response assessment in predicting the outcome in patients with colorectal liver metastases treated with neo-adjuvant chemotherapy followed by liver surgery. Ann Oncol 18, 299–304 (2007).

61. Nierop, P.M., Hoppener, D.J., Buisman, F.E., van der Stok, E.P., Galjart, B., Balachandran, V.P. et al. Preoperative systemic chemotherapy alters the histopathological growth patterns of colorectal liver metastases. J Pathol Clin Res 8, 48–64 (2022).

62. Gomez Dorronsoro, M.L., Vera, R., Ortega, L., Plaza, C., Miquel, R., Garcia, M., et al. Recommendations of a group of experts for the pathological assessment of tumour regression of liver metastases of colorectal cancer and damage of non-tumour liver tissue after neoadjuvant therapy. Clin Transl Oncol 16, 234–42 (2014).

63. Powley, I.R., Patel, M., Miles, G., Pringle, H., Howells, L., Thomas, A. et al. Patient- derived explants (PDEs) as a powerful preclinical platform for anti-cancer drug and biomarker discovery. Br J Cancer 122, 735–744 (2020).

64. Voabil, P., de Bruijn, M., Roelofsen, L.M., Hendriks, S.H., Brokamp, S., van den Braber, M., et al. An ex vivo tumor fragment platform to dissect response to PD-1 blockade in cancer. Nat Med 27, 1250–1261 (2021).

65. Fornabaio, G., Barnhill, R.L., Lugassy, C., Bentolila, L.A., Cassoux, N., Roman-Roman, S. et al. Angiotropism and extravascular migratory metastasis in cutaneous and uveal melanoma progression in a zebrafish model. Sci Rep 8, 10448 (2018).

66. Latacz, E., van Dam, P.J., Vanhove, C., Llado, L., Descamps, B., Ruiz, N. et al. Can medical imaging identify the histopathological growth patterns of liver metastases? Semin Cancer Biol 71, 33–41 (2021).

67. Wei, S., Han, Y., Zeng, H., Ye, S., Cheng, J., Chai, F. et al. Radiomics diagnosed histopathological growth pattern in prediction of response and 1-year progression free survival for colorectal liver metastases patients treated with bevacizumab containing chemotherapy. Eur J Radiol 142, 109863 (2021).

68. Er, E.E., Valiente, M., Ganesh, K., Zou, Y., Agrawal, S., Hu, J. et al. Pericyte-like spreading by disseminated cancer cells activates YAP and MRTF for metastatic colonization. Nat Cell Biol 20, 966–978 (2018).

69. Suzuki, K., Tanaka, M., Watanabe, N., Saito, S., Nonaka, H. & Miyajima, A. p75 Neurotrophin receptor is a marker for precursors of stellate cells and portal fibroblasts in mouse fetal liver. Gastroenterology 135, 270–281 e3 (2008).

70. Wells, R.G. The portal fibroblast: not just a poor man’s stellate cell. Gastroenterology 147, 41–7 (2014).

71. Trim, N., Morgan, S., Evans, M., Issa, R., Fine, D., Afford, S. et al. Hepatic stellate cells express the low affinity nerve growth factor receptor p75 and undergo apoptosis in response to nerve growth factor stimulation. Am J Pathol 156, 1235–43 (2000).

72. Passino, M.A., Adams, R.A., Sikorski, S.L. & Akassoglou, K. Regulation of hepatic stellate cell differentiation by the neurotrophin receptor p75NTR. Science 315, 1853–6 (2007).

73. Kendall, T.J., Hennedige, S., Aucott, R.L., Hartland, S.N., Vernon, M.A., Benyon, R.C. et al. p75 Neurotrophin receptor signaling regulates hepatic myofibroblast proliferation and apoptosis in recovery from rodent liver fibrosis. Hepatology 49, 901–10 (2009).

74. Aimaiti, Y., Jin, X., Shao, Y., Wang, W. & Li, D. Hepatic stellate cells regulate hepatic progenitor cells differentiation via the TGF-beta1/Jagged1 signaling axis. J Cell Physiol 234, 9283–9296 (2019).

75. Cassiman, D., Denef, C., Desmet, V.J. & Roskams, T. Human and rat hepatic stellate cells express neurotrophins and neurotrophin receptors. Hepatology 33, 148–58 (2001).

76. Vidal-Vanaclocha, F., Crende, O., Garcia de Durango, C., Herreros-Pomares, A., Lopez- Domenech, S., Gonzalez, A., et al. Liver prometastatic reaction: Stimulating factors and responsive cancer phenotypes. Semin Cancer Biol 71, 122–133 (2021).

77. Ciner, A.T., Jones, K., Muschel, R.J. & Brodt, P. The unique immune microenvironment of liver metastases: Challenges and opportunities. Semin Cancer Biol 71, 143–156 (2021).

78. Kurebayashi, Y., Kubota, N. & Sakamoto, M. Immune microenvironment of hepatocellular carcinoma, intrahepatic cholangiocarcinoma and liver metastasis of colorectal adenocarcinoma: Relationship with histopathological and molecular classifications. Hepatol Res 51, 5–18 (2021).

79. Bohlok, A., Duran Derijckere, I., Azema, H., Lucidi, V., Vankerckhove, S., Hendlisz, A. et al. Clinico-metabolic characterization improves the prognostic value of histological growth patterns in patients undergoing surgery for colorectal liver metastases. J Surg Oncol 123, 1773–1783 (2021).

80. Matsumoto, K., Yoshitomi, H., Rossant, J. & Zaret, K.S. Liver organogenesis promoted by endothelial cells prior to vascular function. Science 294, 559–63 (2001).

81. Crivellato, E., Nico, B. & Ribatti, D. Contribution of endothelial cells to organogenesis: a modern reappraisal of an old Aristotelian concept. J Anat 211, 415–27 (2007).

82. Ding, B.S., Cao, Z., Lis, R., Nolan, D.J., Guo, P., Simons, M. et al. Divergent angiocrine signals from vascular niche balance liver regeneration and fibrosis. Nature 505, 97–102 (2014).

83. Daniel, E. & Cleaver, O. Vascularizing organogenesis: Lessons from developmental biology and implications for regenerative medicine. Curr Top Dev Biol 132, 177–220 (2019).

84. Muthuswamy, S.K. Self-organization in cancer: Implications for histopathology, cancer cell biology, and metastasis. Cancer Cell 39, 443–446 (2021).

85. Pezzella, F., Di Bacco, A., Andreola, S., Nicholson, A.G., Pastorino, U. & Harris, A.L. Angiogenesis in primary lung cancer and lung secondaries. Eur J Cancer **32A**, 2494–500 (1996).

86. Evidence for novel non-angiogenic pathway in breast-cancer metastasis. Breast Cancer Progression Working Party. Lancet 355, 1787-8 (2000).

87. Teuwen, L.A., De Rooij, L., Cuypers, A., Rohlenova, K., Dumas, S.J., Garcia-Caballero, M. et al. Tumor vessel co-option probed by single-cell analysis. Cell Rep 35, 109253 (2021).

88. Lee, J.C., Green, M.D., Huppert, L.A., Chow, C., Pierce, R.H. & Daud, A.I. The Liver- Immunity Nexus and Cancer Immunotherapy. Clin Cancer Res 28, 5–12 (2022).

89. Garcia-Vicién, G., Mezheyeuski, A., Micke, P., Ruiz, N., Ruffinelli, J.C., Mils, K. et al. Spatial Immunology in Liver Metastases from Colorectal Carcinoma according to the Histologic Growth Pattern. Cancers 14, 689 (2022).

90. Szczepanski, J.M., Mendiratta-Lala, M., Fang, J.M., Choi, W.T., Karamchandani, D.M. & Westerhoff, M. Sinusoidal Growth Pattern of Hepatic Melanoma Metastasis: Implications for Histopathologic Diagnosis. Am J Surg Pathol (2021).

91. Li, W.H., Wang, S., Liu, Y., Wang, X.F., Wang, Y.F. & Chai, R.M. Differentiation of histopathological growth patterns of colorectal liver metastases by MRI features. Quant Imaging Med Surg 12, 608–617 (2022).

92. de Ridder, J.A., Knijn, N., Wiering, B., de Wilt, J.H. & Nagtegaal, I.D. Lymphatic Invasion is an Independent Adverse Prognostic Factor in Patients with Colorectal Liver Metastasis. Ann Surg Oncol 22 **Suppl 3**, S638–45 (2015).

93. Serrablo, A., Paliogiannis, P., Pulighe, F., Moro, S.S., Borrego-Estella, V., Attene, F. et al. Impact of novel histopathological factors on the outcomes of liver surgery for colorectal cancer metastases. Eur J Surg Oncol 42, 1268–77 (2016).

94. Fonseca, G.M., de Mello, E.S., Faraj, S.F., Kruger, J.A.P., Coelho, F.F., Jeismann, V.B. et al. Prognostic significance of poorly differentiated clusters and tumor budding in colorectal liver metastases. J Surg Oncol 117, 1364–1375 (2018).

95. Cremolini, C., Milione, M., Marmorino, F., Morano, F., Zucchelli, G., Mennitto, A. et al. Differential histopathologic parameters in colorectal cancer liver metastases resected after triplets plus bevacizumab or cetuximab: a pooled analysis of five prospective trials. Br J Cancer 118, 955–965 (2018).

96. Falcao, D., Alexandrino, H., Caetano Oliveira, R., Martins, J., Ferreira, L., Martins, R. et al. Histopathologic patterns as markers of prognosis in patients undergoing hepatectomy for colorectal cancer liver metastases - Pushing growth as an independent risk factor for decreased survival. Eur J Surg Oncol 44, 1212–1219 (2018).

97. Ao, T., Kajiwara, Y., Yonemura, K., Shinto, E., Mochizuki, S., Okamoto, K. et al. Prognostic significance of histological categorization of desmoplastic reaction in colorectal liver metastases. Virchows Arch 475, 341–348 (2019).

98. Zhang, Y., Luo, X., Lin, J., Fu, S., Feng, P., Su, H. et al. Gelsolin Promotes Cancer Progression by Regulating Epithelial-Mesenchymal Transition in Hepatocellular Carcinoma and Correlates with a Poor Prognosis. J Oncol 2020, 1980368 (2020).

99. Baldin, P., Van den Eynde, M., Mlecnik, B., Bindea, G., Beniuga, G., Carrasco, J., et al. Prognostic assessment of resected colorectal liver metastases integrating pathological features, RAS mutation and Immunoscore. J Pathol Clin Res 7, 27–41 (2021).

100. Temido, M.J., Caetano Oliveira, R., Martins, R., Serodio, M., Costa, B., Carvalho, C. et al. Prognostic Factors After Hepatectomy for Gastric Adenocarcinoma Liver Metastases: Desmoplastic Growth Pattern as the Key to Improved Overall Survival. Cancer Manag Res 12, 11689–11699 (2020).

101. Jayme, V.R., Fonseca, G.M., Amaral, I.M.A., Coelho, F.F., Kruger, J.A.P., Jeismann, V.B. et al. Infiltrative Tumor Borders in Colorectal Liver Metastasis: Should We Enlarge Margin Size? Ann Surg Oncol 28, 7636–7646 (2021).

102. Zhang, Y.L., He, H.J., Cheng, J. & Shen, D.H. [Value of histopathological growth pattern in predicting 3-year progression free survival after operation in patients with liver metastasis of colorectal cancer]. Zhonghua Bing Li Xue Za Zhi 50, 26–31 (2021).

103. Meyer, Y.M., Beumer, B.R., Hoppener, D.J., Nierop, P.M.H., Doukas, M., de Wilde, R.F. et al. Histopathological growth patterns modify the prognostic impact of microvascular invasion in non-cirrhotic hepatocellular carcinoma. HPB (Oxford) (2021).

104. Vles, M.D., Hoppener, D.J., Galjart, B., Moelker, A., Vermeulen, P.B., Grunhagen, D.J. et al. Local tumour control after radiofrequency or microwave ablation for colorectal liver metastases in relation to histopathological growth patterns. HPB (Oxford) (2022).

105. Ceausu, A.R., Ciolofan, A., Cimpean, A.M., Magheti, A., Mederle, O. & Raica, M. The Mesenchymal-Epithelial and Epithelial-Mesenchymal Cellular Plasticity of Liver Metastases with Digestive Origin. Anticancer Res 38, 811–816 (2018).

106. Lazaris, A., Amri, A., Petrillo, S.K., Zoroquiain, P., Ibrahim, N., Salman, A. et al. Vascularization of colorectal carcinoma liver metastasis: insight into stratification of patients for anti-angiogenic therapies. J Pathol Clin Res 4, 184–192 (2018).

107. Wu, J.B., Sarmiento, A.L., Fiset, P.O., Lazaris, A., Metrakos, P., Petrillo, S. et al. Histologic features and genomic alterations of primary colorectal adenocarcinoma predict growth patterns of liver metastasis. World J Gastroenterol 25, 3408-3425 (2019).

108. Blank, A., Schenker, C., Dawson, H., Beldi, G., Zlobec, I. & Lugli, A. Evaluation of Tumor Budding in Primary Colorectal Cancer and Corresponding Liver Metastases Based on H&E and Pancytokeratin Staining. Front Med (Lausanne) 6, 247 (2019).

109. Palmieri, V., Lazaris, A., Mayer, T.Z., Petrillo, S.K., Alamri, H., Rada, M. et al. Neutrophils expressing lysyl oxidase-like 4 protein are present in colorectal cancer liver metastases resistant to anti-angiogenic therapy. J Pathol 251, 213–223 (2020).

110. Ao, T., Kajiwara, Y., Yonemura, K., Shinto, E., Mochizuki, S., Okamoto, K. et al. Morphological consistency of desmoplastic reactions between the primary colorectal cancer lesion and associated metastatic lesions. Virchows Arch 477, 47–55 (2020).

111. Rada, M., Kapelanski-Lamoureux, A., Petrillo, S., Tabaries, S., Siegel, P., Reynolds, A.R. et al. Runt related transcription factor-1 plays a central role in vessel co-option of colorectal cancer liver metastases. Commun Biol 4, 950 (2021).

112. Burren, S., Reche, K., Blank, A., Galvan, J.A., Dawson, H., Berger, M.D. et al. RHAMM in liver metastases of stage IV colorectal cancer with mismatch-repair proficient status correlates with tumor budding, cytotoxic T-cells and PD-1/PD-L1. Pathol Res Pract 223, 153486 (2021).

113. Donnem, T., Reynolds, A.R., Kuczynski, E.A., Gatter, K., Vermeulen, P.B., Kerbel, R.S. et al. Non-angiogenic tumours and their influence on cancer biology. Nat Rev Cancer 18, 323–336 (2018).

114. Baldin, P., Van den Eynde, M., Hubert, C., Jouret-Mourin, A. & Komuta, M. The role of the pathologist and clinical implications in colorectal liver metastasis. Acta Gastroenterol Belg 81, 419–426 (2018).

115. Kuczynski, E.A., Vermeulen, P.B., Pezzella, F., Kerbel, R.S. & Reynolds, A.R. Vessel co- option in cancer. Nat Rev Clin Oncol 16, 469–493 (2019).

116. Oliveira, R.C., Alexandrino, H., Cipriano, M.A. & Tralhao, J.G. Liver Metastases and Histological Growth Patterns: Biological Behavior and Potential Clinical Implications- Another Path to Individualized Medicine? J Oncol 2019, 6280347 (2019).

117. Kuczynski, E.A. & Reynolds, A.R. Vessel co-option and resistance to anti-angiogenic therapy. Angiogenesis 23, 55–74 (2020).

118. Oliveira, R.C., Alexandrino, H., Cipriano, M.A., Alves, F.C. & Tralhao, J.G. Predicting liver metastases growth patterns: Current status and future possibilities. Semin Cancer Biol 71, 42–51 (2021).

119. Rigamonti, A., Feuerhake, F., Donadon, M., Locati, M. & Marchesi, F. Histopathological and Immune Prognostic Factors in Colo-Rectal Liver Metastases. Cancers (Basel) 13, (2021).

120. Garcia-Vicien, G., Mezheyeuski, A., Banuls, M., Ruiz-Roig, N. & Mollevi, D.G. The Tumor Microenvironment in Liver Metastases from Colorectal Carcinoma in the Context of the Histologic Growth Patterns. Int J Mol Sci 22, (2021).

121. Haas, G., Fan, S., Ghadimi, M., De Oliveira, T. & Conradi, L.C. Different Forms of Tumor Vascularization and Their Clinical Implications Focusing on Vessel Co-option in Colorectal Cancer Liver Metastases. Front Cell Dev Biol 9, 612774 (2021).

122. Rompianesi, G., Pegoraro, F., Ceresa, C.D., Montalti, R. & Troisi, R.I. Artificial intelligence in the diagnosis and management of colorectal cancer liver metastases. World J Gastroenterol 28, 108–122 (2022).

123. Paku, S. & Lapis, K. Morphological aspects of angiogenesis in experimental liver metastases. Am J Pathol 143, 926–36 (1993).

124. Vidal-Vanaclocha, F. The prometastatic microenvironment of the liver. Cancer Microenviron 1, 113–29 (2008).

125. Barsky, S.H., Doberneck, S.A., Sternlicht, M.D., Grossman, D.A. & Love, S.M. ’Revertant’ DCIS in human axillary breast carcinoma metastases. J Pathol 183, 188–94 (1997).

126. Schuppan, D. & Kim, Y.O. Evolving therapies for liver fibrosis. J Clin Invest 123, 1887–901 (2013).

127. Dezso, K., Papp, V., Bugyik, E., Hegyesi, H., Safrany, G., Bodor, C. et al. Structural analysis of oval-cell-mediated liver regeneration in rats. Hepatology 56, 1457–67 (2012).

